# Antibody levels following vaccination against SARS-CoV-2: associations with post-vaccination infection and risk factors

**DOI:** 10.1101/2022.05.19.22275214

**Authors:** Nathan J. Cheetham, Milla Kibble, Andrew Wong, Richard J. Silverwood, Anika Knuppel, Dylan M. Williams, Olivia K. L. Hamilton, Paul H. Lee, Charis Bridger Staatz, Giorgio Di Gessa, Jingmin Zhu, Srinivasa Vittal Katikireddi, George B. Ploubidis, Ellen J. Thompson, Ruth C. E. Bowyer, Xinyuan Zhang, Golboo Abbasian, Maria Paz Garcia, Deborah Hart, Jeffrey Seow, Carl Graham, Neophytos Kouphou, Sam Acors, Michael H. Malim, Ruth E. Mitchell, Kate Northstone, Daniel Major-Smith, Sarah Matthews, Thomas Breeze, Michael Crawford, Lynn Molloy, Alex S. F. Kwong, Katie J. Doores, Nishi Chaturvedi, Emma L. Duncan, Nicholas J. Timpson, Claire J. Steves

**Author notes:** Correspondence to: Claire J. Steves, Nicholas J. Timpson.

## Abstract

SARS-CoV-2 antibody levels can be used to assess humoral immune responses following SARS-CoV-2 infection or vaccination, and may predict risk of future infection. From cross-sectional antibody testing of 9,361 individuals from TwinsUK and ALSPAC UK population-based longitudinal studies (jointly in April-May 2021, and TwinsUK only in November 2021-January 2022), we tested associations between antibody levels following vaccination and: (1) SARS-CoV-2 infection following vaccination(s); (2) health, socio-demographic, SARS-CoV-2 infection and SARS-CoV-2 vaccination variables.

Within TwinsUK, single-vaccinated individuals with the lowest 20% of anti-Spike antibody levels at initial testing had 3-fold greater odds of SARS-CoV-2 infection over the next six to nine months, compared to the top 20%. In TwinsUK and ALSPAC, individuals identified as at increased risk of COVID-19 complication through the UK “Shielded Patient List” had consistently greater odds (2 to 4-fold) of having antibody levels in the lowest 10%. Third vaccination increased absolute antibody levels for almost all individuals, and reduced relative disparities compared with earlier vaccinations.

These findings quantify the association between antibody level and risk of subsequent infection, and support a policy of triple vaccination for the generation of protective antibodies.

**Lay summary:** In this study, we analysed blood samples from 9,361 participants from two studies in the UK: an adult twin registry, TwinsUK (4,739 individuals); and the Avon Longitudinal Study of Parents and Children, ALSPAC (4,622 individuals). We did this work as part of the UK Government National Core Studies initiative researching COVID-19. We measured blood antibodies which are specific to SARS-CoV-2 (which causes COVID-19). Having a third COVID-19 vaccination boosted antibody levels. More than 90% of people from TwinsUK had levels after third vaccination that were greater than the average level after second vaccination. Importantly, this was the case even in individuals on the UK “Shielded Patient List”. We found that people with lower antibody levels after first vaccination were more likely to report having COVID-19 later on, compared to people with higher antibody levels. People on the UK “Shielded Patient List”, and individuals who reported that they had poorer general health, were more likely to have lower antibody levels after vaccination. In contrast, people who had had a previous COVID-19 infection were more likely to have higher antibody levels following vaccination compared to people without infection. People receiving the Oxford/AstraZeneca rather than the Pfizer BioNTech vaccine had lower antibody levels after one or two vaccinations. However, after a third vaccination, there was no difference in antibody levels between those who had Oxford/AstraZeneca and Pfizer BioNTech vaccines for their first two doses. These findings support having a third COVID-19 vaccination to boost antibodies.

## Introduction

Immunological responses to severe acute respiratory syndrome coronavirus 2 (SARS-CoV-2) infection and SARS-CoV-2 vaccination vary between individuals and over time [1–3]. Within two to four weeks of infection, most individuals generate detectable levels of several antibody subtypes (Immunoglobulin A, M, G) directed against different domains of the virus (Nucleocapsid protein, Spike protein, receptor-binding domain of Spike), which gradually decline over time [4–7]. Levels of anti-Spike antibodies correlate with SARS-CoV-2-neutralising anti-receptor-binding domain antibody titre [8]. A similar profile of antibody induction with subsequent waning is observed after vaccination against SARS-CoV-2 [1,2,9,10]. Anti-Nucleocapsid antibodies are generated by infection but not by any current SARS-CoV-2 vaccines. Thus, presence of Anti-Nucleocapsid antibodies distinguishes between antibody responses due to infection versus vaccination.

The assumption that higher antibody levels, whether from vaccination or prior infection, decrease risk of subsequent SARS-CoV-2 infection was supported by early reports that showed that higher anti-Spike and anti-receptor-binding domain levels after two-dose vaccination correlated with increased protection against subsequent infection [11–13]. Thus, the extent of initial antibody induction and subsequent decrease over time may correlate with individual variability in vaccine effectiveness [14–16]. In addition to temporal changes in antibody levels, the neutralisation ability of generated antibodies and overall vaccine effectiveness varies according to the virulence and divergence from the ancestral Spike of emerging variants. Observations suggest reduced neutralisation and lower vaccine effectiveness for newer variants versus unmutated ‘wild-type’ and older variants of SARS-CoV-2, in both double- [15,17,18], and triple-[15,18,19], vaccinated individuals. In addition, early studies report that triple vaccination increases both antibody levels and protection against symptomatic infection, disease severity, and death, compared with double vaccination [19–26].

Several clinical variables contribute to variation in antibody response following vaccination. Lower antibody levels following both first and second vaccinations have been observed in individuals with particular comorbidities (including cancer [27,28], renal disease, hepatic disease, and anti-neutrophil cytoplasmic autoantibody-associated vasculitis [29]), individuals using immunosuppressant medications [1,2,27,28], and individuals identified from electronic health records data as of moderate or high risk of COVID-19 complications (according to UK government prior “Shielded Patient List” criteria of conditions, ongoing treatments and medications) [1,2,30]. Studies testing for associations between antibody response and non-clinical factors such as socio-demographics have been more limited. Here, the use of longitudinal studies, with broader catalogues of bio-social data, are well-suited to such investigations.

To date, studies directly testing antibody levels following SARS-CoV-2 vaccination as a correlate of subsequent risk of infection have been limited to risk after two vaccinations only, examining relative antibody levels [11], or absolute antibody levels in vaccine trial populations before the emergence of Delta and Omicron variants [12,13]. While COVID-19 severity [31], and some symptoms [32], have been shown to be heritable, no studies have yet examined heritability of the immune response (humoral or cell-mediated) following vaccination.

Here, we aimed to examine both the consequences and origins of variation in post-vaccination antibody levels. We measured the antibody levels of participants from two population-based longitudinal cohorts during the time of the UK vaccination roll-out: TwinsUK (in April-May 2021 and November 2021-January 2022) [33] and Avon Longitudinal Study of Parents and Children (ALSPAC) [34,35] (April-May 2021 only). We aimed firstly to assess the relationship between anti-Spike antibody levels (used as a proxy for the humoral immune response), measured after first or second vaccination in April-May 2021, and an outcome of subsequent post-vaccination infection over the following six to nine months (identified through further serological evidence and/or self-reported COVID-19 from data collected in TwinsUK between November 2021 and January 2022). Secondly, we used peri-pandemic and historical data to investigate associations with an outcome of having relatively low antibody levels following first, second (ALSPAC and TwinsUK) or third (TwinsUK only) vaccination, for multiple socio-demographic, physical and mental health characteristics, prior SARS-CoV-2 infection, and vaccination history. Finally, we used twin-pair analysis within TwinsUK to probe genetic and environmental contributions to antibody level variation.

## Results

### Cohort characteristics

Antibody levels were measured in 9,361 individuals at two time points – 4,256 individuals from TwinsUK and 4,622 individuals from ALSPAC during April and May 2021 (referred to throughout as Q2 [calendar year quarter 2] testing), and 3,575 individuals from TwinsUK in follow-up testing from November 2021 to January 2022 (referred to throughout as Q4 [quarter 4] testing). Response rates, as the percentage who returned sample after consenting and being sent a sample collection kit, were as follows: TwinsUK Q2: 87%, TwinsUK Q4: 80%, ALSPAC Q2: 79%. Flow charts showing identification of analysis samples are given in Figure S 1 to Figure S 3. Results of antibody testing and selected characteristics are summarised in Table 2 (with extended characteristics given in Table S 2). Consistent with the tiered UK vaccination campaign, individuals who had received more vaccinations at either timepoint were older, more likely to be on the UK “Shielded Patient List” (criteria provided in supplementary information), and had lower self-rated health, compared with those with fewer vaccinations. Participants were predominantly female and the vast majority were of white ethnicity in both cohorts, consistent with the broader composition of both cohorts.

**Table 1.**
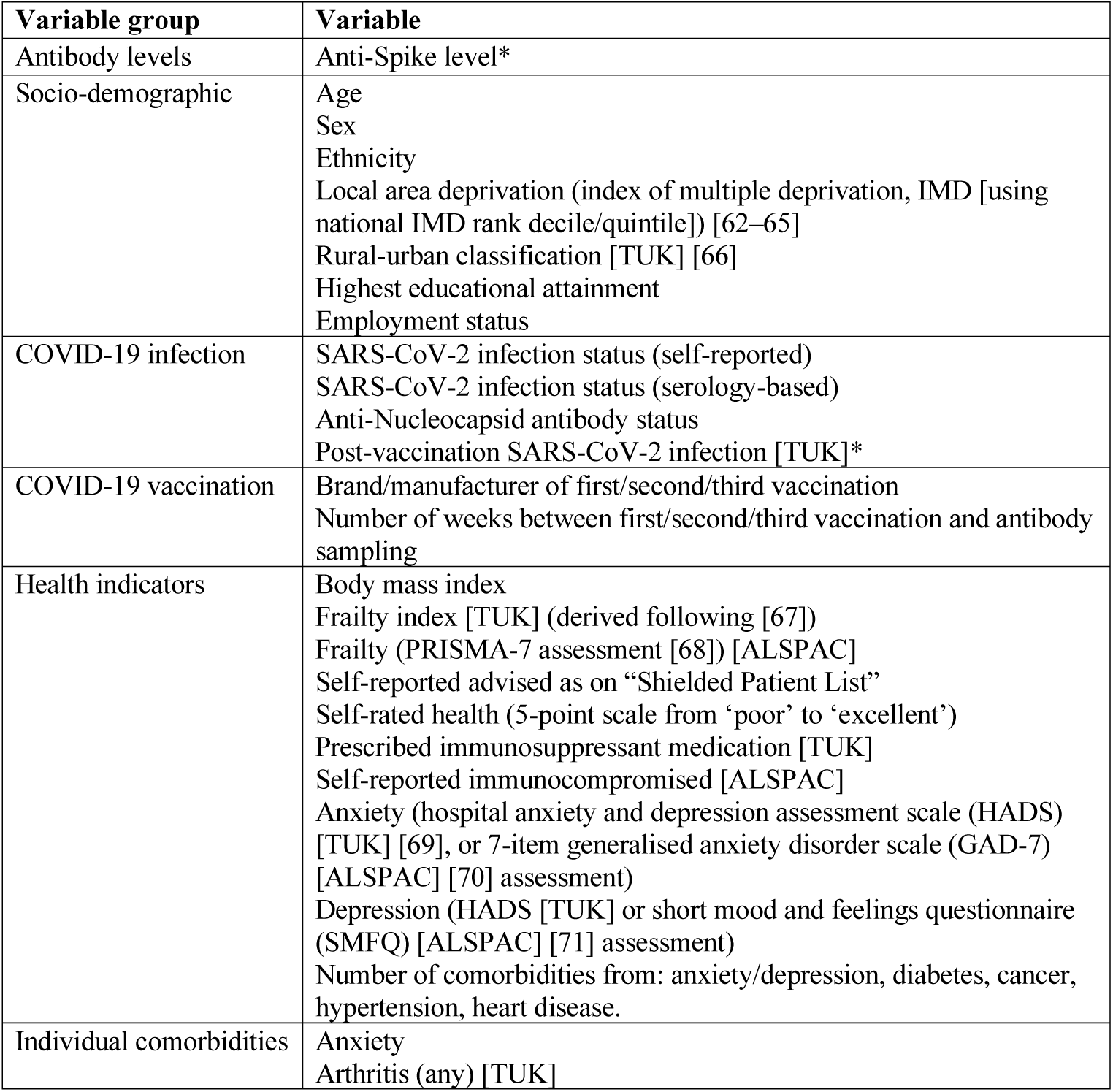
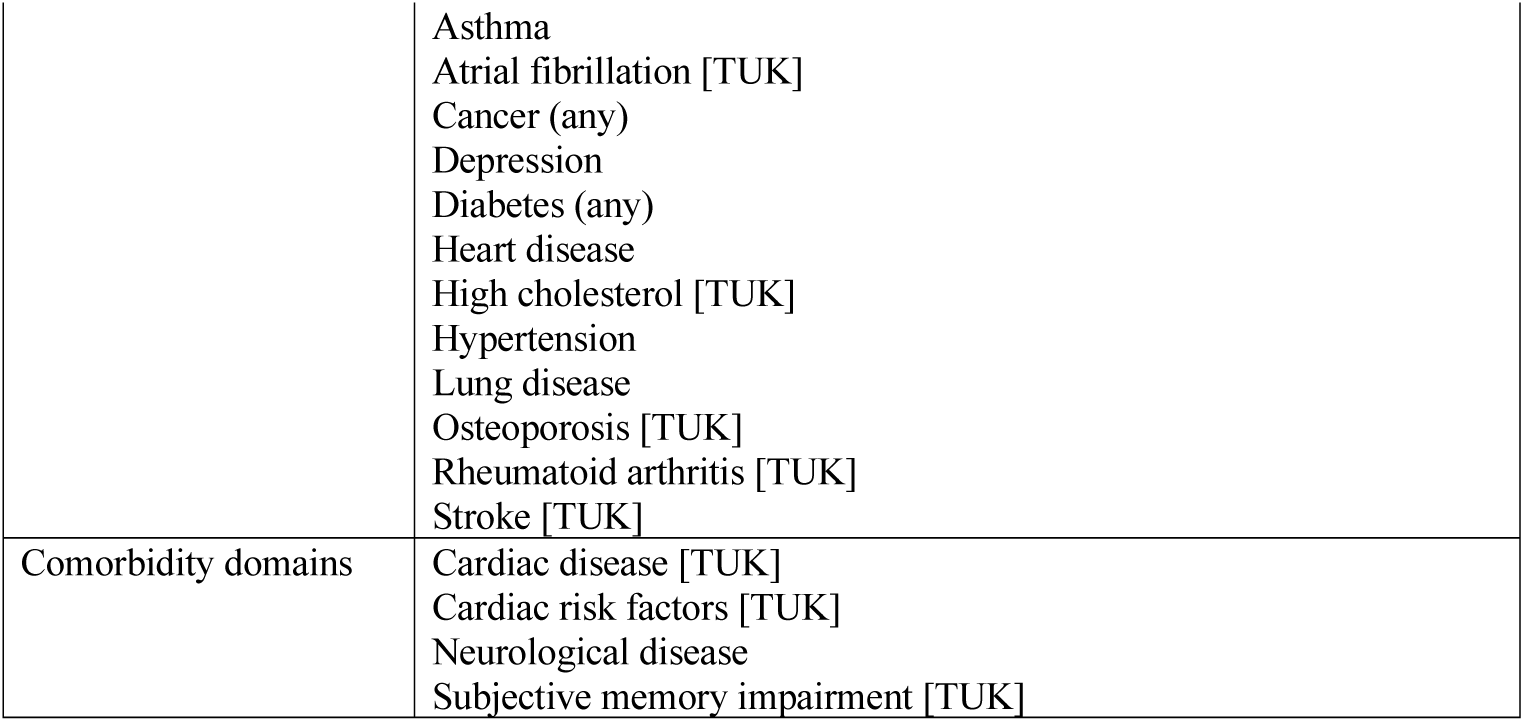
Phenotypic variables used in analyses. Variables marked with an asterisk were outcome variables in logistic regression analyses; all other variables were adjustment or exposure variables. Variables only available in TwinsUK are notated as [TUK], and those only in ALSPAC as [ALSPAC].

| Variable group | Variable |
| --- | --- |
| Antibody levels | Anti-Spike level* |
| Socio-demographic | Age<br>Sex<br>Ethnicity<br>Local area deprivation (index of multiple deprivation, IMD [using national IMD rank decile/quintile]) [62–65]<br>Rural-urban classification [TUK] [66]<br>Highest educational attainment<br>Employment status |
| COVID-19 infection | SARS-CoV-2 infection status (self-reported)<br>SARS-CoV-2 infection status (serology-based)<br>Anti-Nucleocapsid antibody status<br>Post-vaccination SARS-CoV-2 infection [TUK]* |
| COVID-19 vaccination | Brand/manufacture of first/second/third vaccination<br>Number of weeks between first/second/third vaccination and antibody sampling |
| Health indicators | Body mass index<br>Frailty index [TUK] (derived following [67])<br>Frailty (PRISMA-7 assessment [68]) [ALSPAC]<br>Self-reported advised as on “Shielded Patient List”<br>Self-rated health (5-point scale from ‘poor’ to ‘excellent’)<br>Prescribed immunosuppressant medication [TUK]<br>Self-reported immunocompromised [ALSPAC]<br>Anxiety (hospital anxiety and depression assessment scale (HADS) [TUK] [69], or 7-item generalised anxiety disorder scale (GAD-7) [ALSPAC] [70] assessment)<br>Depression (HADS [TUK] or short mood and feelings questionnaire (SMFQ) [ALSPAC] [71] assessment)<br>Number of comorbidities from: anxiety/depression, diabetes, cancer, hypertension, heart disease. |
| Individual comorbidities | Anxiety<br>Arthritis (any) [TUK] |
|  | Asthma<br>Atrial fibrillation [TUK]<br>Cancer (any)<br>Depression<br>Diabetes (any)<br>Heart disease<br>High cholesterol [TUK]<br>Hypertension<br>Lung disease<br>Osteoporosis [TUK]<br>Rheumatoid arthritis [TUK]<br>Stroke [TUK] |
| Comorbidity domains | Cardiac disease [TUK]<br>Cardiac risk factors [TUK]<br>Neurological disease<br>Subjective memory impairment [TUK] |

**Table 2.**
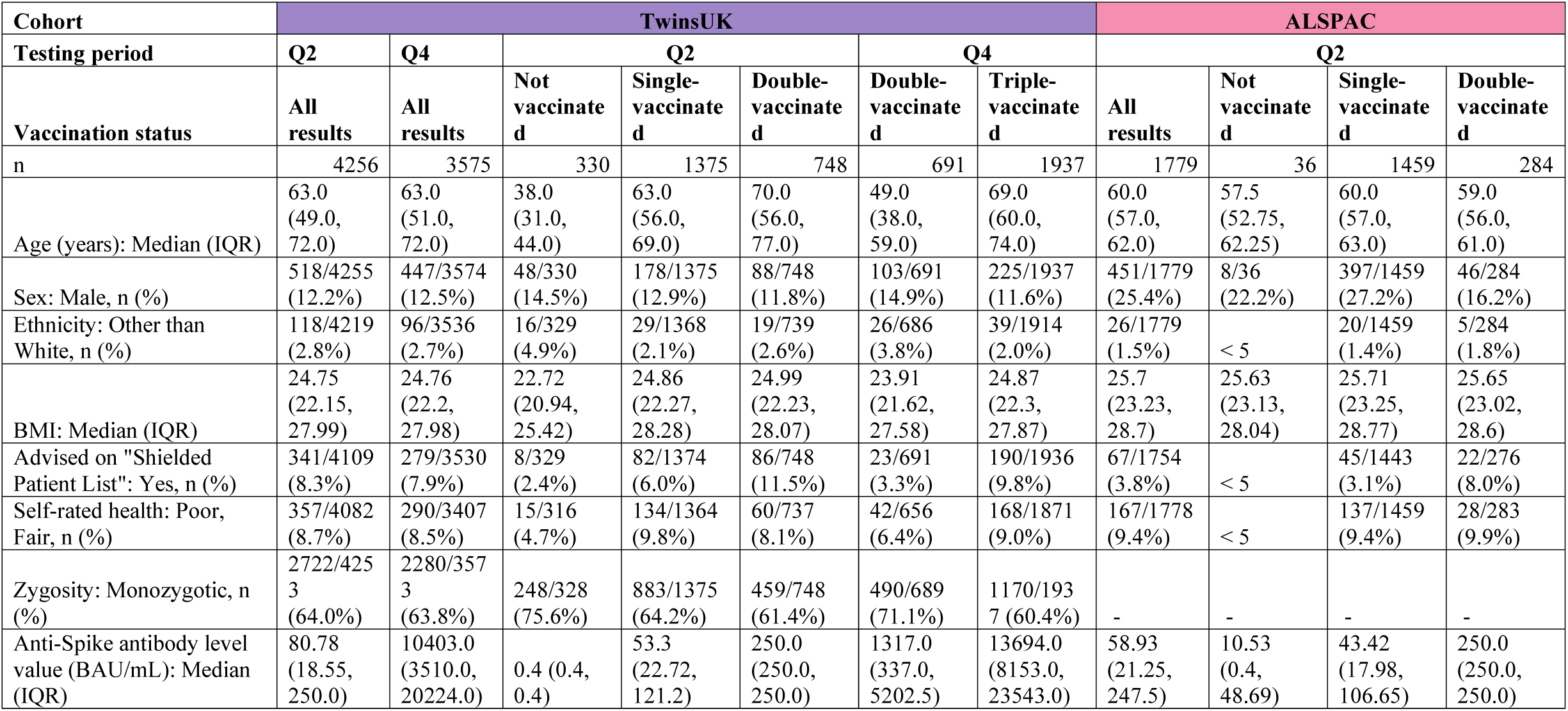
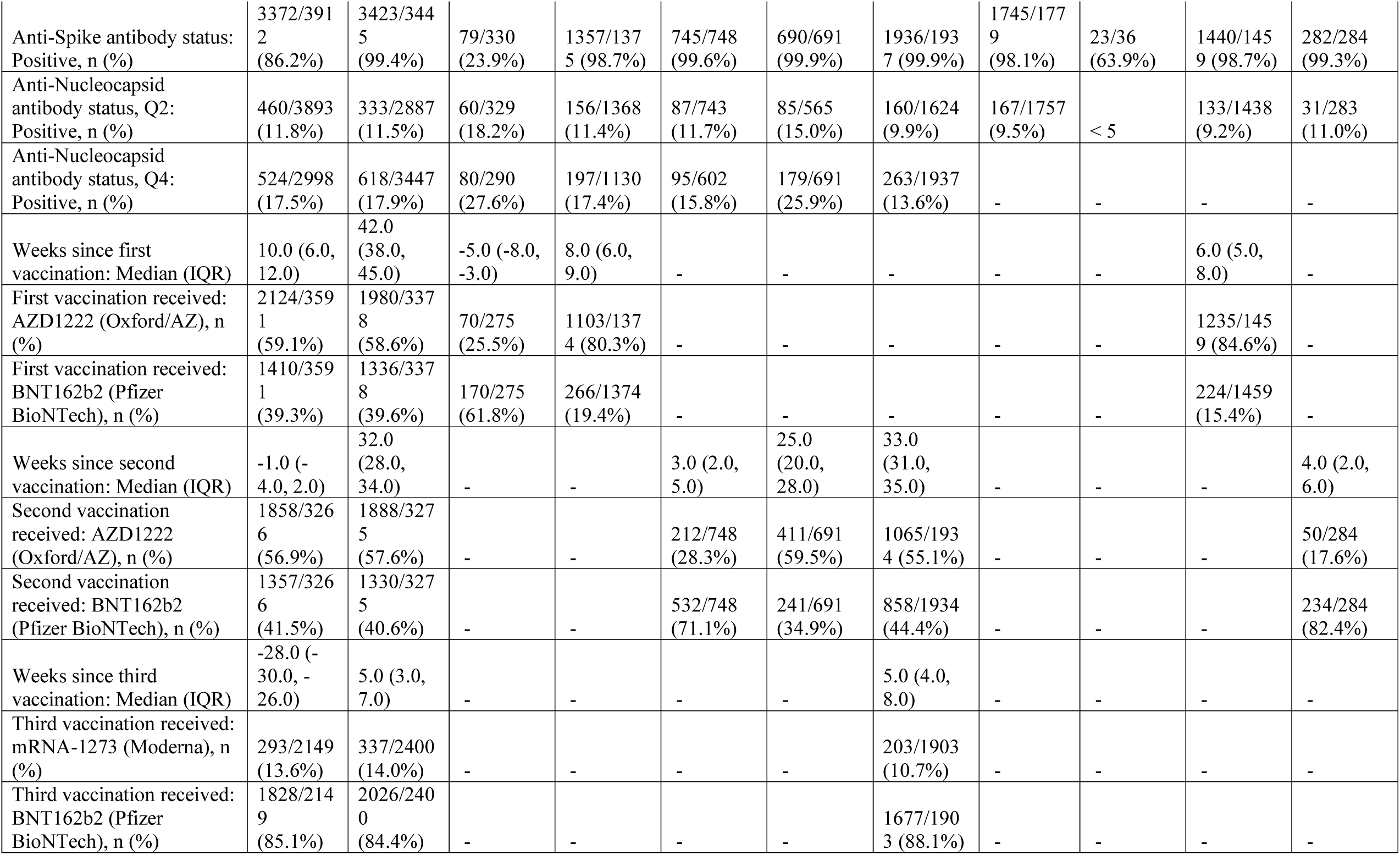
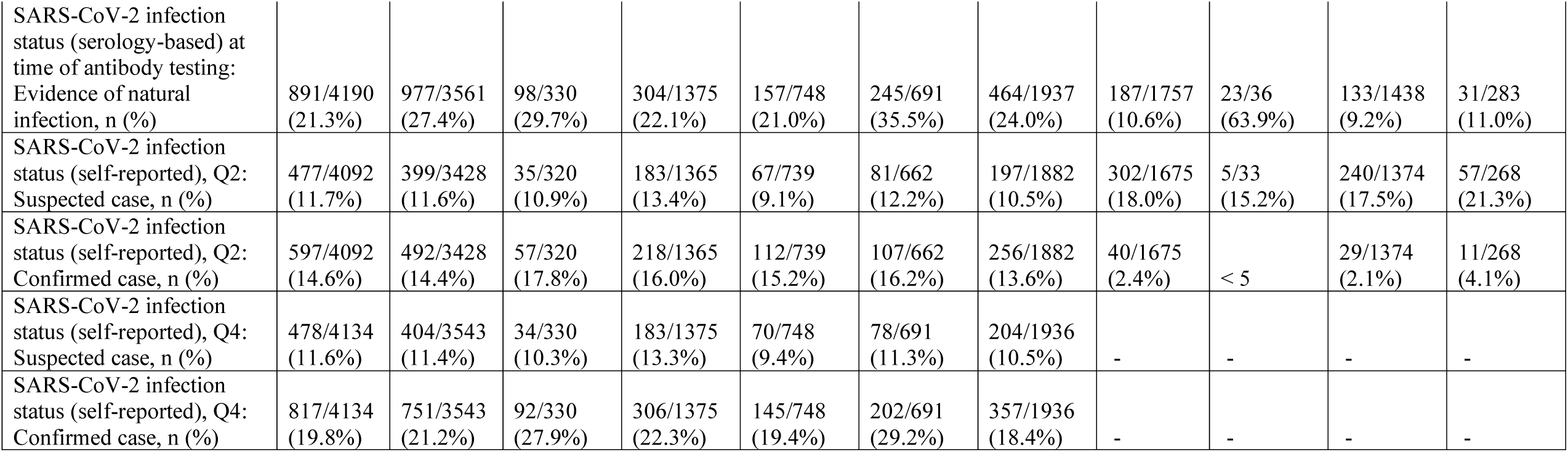
Sample characteristics. Antibody level values and characteristics for TwinsUK and ALSPAC individuals sampled in Q2 and Q4 antibody collections. Individuals are stratified by vaccination status at time of sampling. Data shown for individuals sampled at least 4 weeks after first vaccination, and at least 2 weeks after second or third vaccination. The anti-Spike antibody level assay range is 0.4 to 250 BAU/mL for Q2 results and 0.4 to 25,000 BAU/mL for Q4 results, with a positive threshold of 0.8 BAU/mL. Categories with fewer than 5 individuals are suppressed.

Prevalence of SARS-CoV-2 infection from self-reported suspected and confirmed cases was 26% in TwinsUK and 20% in ALSPAC at Q2 testing (Table 2), while Q2 testing anti-Nucleocapsid antibody positivity was lower at 12% in TwinsUK and 10% in ALSPAC. Within TwinsUK, incorporating additional serology testing undertaken prior to Q2 (between April 2020 and May 2021) gave a higher (21%) serology-based infection prevalence compared with Q2 testing alone, consistent with previously reported waning in anti-Nucleocapsid levels [4]. As expected, higher prevalence of SARS-CoV-2 infection was observed at Q4 testing within TwinsUK, with 27% of individuals having evidence of infection based on all serology up to and including Q4 testing, 17% with a positive anti-Nucleocapsid result at time of Q4 testing alone, and 33% with self-reported suspected or confirmed cases.

Variation in infection prevalence across socio-demographic variables was observed within TwinsUK at most recent Q4 testing (Table S 3 and Figure S 4). Male sex, other than white ethnicity, those living in urban areas, those living in areas of higher deprivation, and younger age were associated with a higher prevalence in self-reported and/or serology-based infection measures.

Seroprevalence estimates from 24,107 individuals within nine further UK-based longitudinal studies at Q2 testing are presented in Table S 4. The trends with age observed in TwinsUK and ALSPAC are replicated in these cohorts (i.e., higher anti-Spike seropositivity and lower anti-Nucleocapsid seropositivity associated with increasing average age (Figure S 5)).

### Antibody levels after first, second, and third vaccination in TwinsUK

Considering firstly data from Q4 testing undertaken within TwinsUK only, cross-sectional antibody levels following third vaccination were much greater and more sustained, with less inter-individual variability, compared to levels for those with fewer vaccinations. The median anti-Spike antibody levels in individuals who had received a third vaccination (unadjusted for time since vaccination) were over 10-fold higher than for individuals after second vaccination: 13,700 BAU/mL after third vaccination, 1,300 BAU/mL after second vaccination; 50 BAU/mL after first vaccination (Figure 1, detailed univariable splits of anti-Spike levels given in Table S 5). The antibody level distribution after third vaccination was narrower compared with earlier vaccination, with smaller scale-factor differences between those with median and lowest levels, quantified at the 5^th^ & 10^th^ percentiles (among individuals with the same vaccination status). There were large increases in absolute levels for individuals at the bottom of the antibody level distribution after third vaccination: individuals sampled on average four to eight weeks since third vaccination had 10-fold higher median levels and 50-fold higher levels at both 5^th^ and 10^th^ percentiles, compared with double-vaccinated individuals sampled at 28-34 weeks since second vaccination (and approximately two-to 15-fold higher median levels when comparing previously reported levels at three to six weeks since second vaccination [2], with levels at three to six weeks since third vaccination reported here).

**Figure 1.**
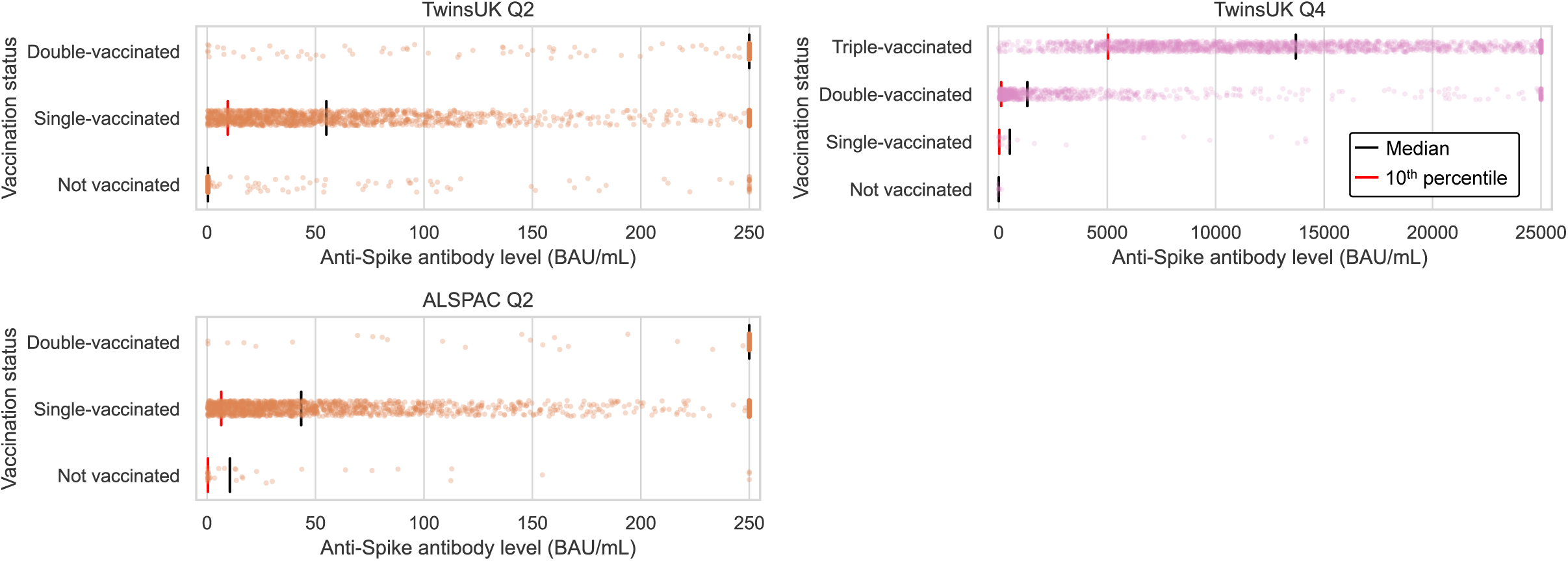
Anti-Spike antibody levels stratified by cohort and vaccination status at Q2 and Q4 antibody testing. Dot plots showing distribution of anti-Spike antibody levels within TwinsUK and ALSPAC, for those not vaccinated or single-, double- or triple-vaccinated individuals at time of sampling. Black lines show median levels and red lines show 10^th^percentile levels. The assay range is 0.4 to 250 BAU/mL for Q2 results and 0.4 to 25,000 BAU/mL for Q4 results, with a positive threshold of 0.8 BAU/mL.

Considering antibody levels versus time since vaccination: within TwinsUK Q4 results, median antibody levels up to 16 weeks since third vaccination were highest in individuals sampled two to three weeks after vaccination (median: 24,600 BAU/mL, n = 203) (Figure 2). Although median antibody levels decreased between two and eight weeks after third vaccination, there was no evidence of further decline between eight and 16 weeks (Mann-Kendall trend test in median levels at 8+ weeks, p = 0.60), and high absolute levels of antibodies were sustained (8+ weeks median = 9,200 BAU/mL [IQR: 5,800-16,000 BAU/mL], n = 519). These cross-sectional trends in median antibody levels versus time since third vaccination persisted when stratifying by age and other variables. Similarly, for individuals sampled 13 to 33 weeks after second vaccination, longer time since vaccination was also associated with lower antibody levels.

**Figure 2.**
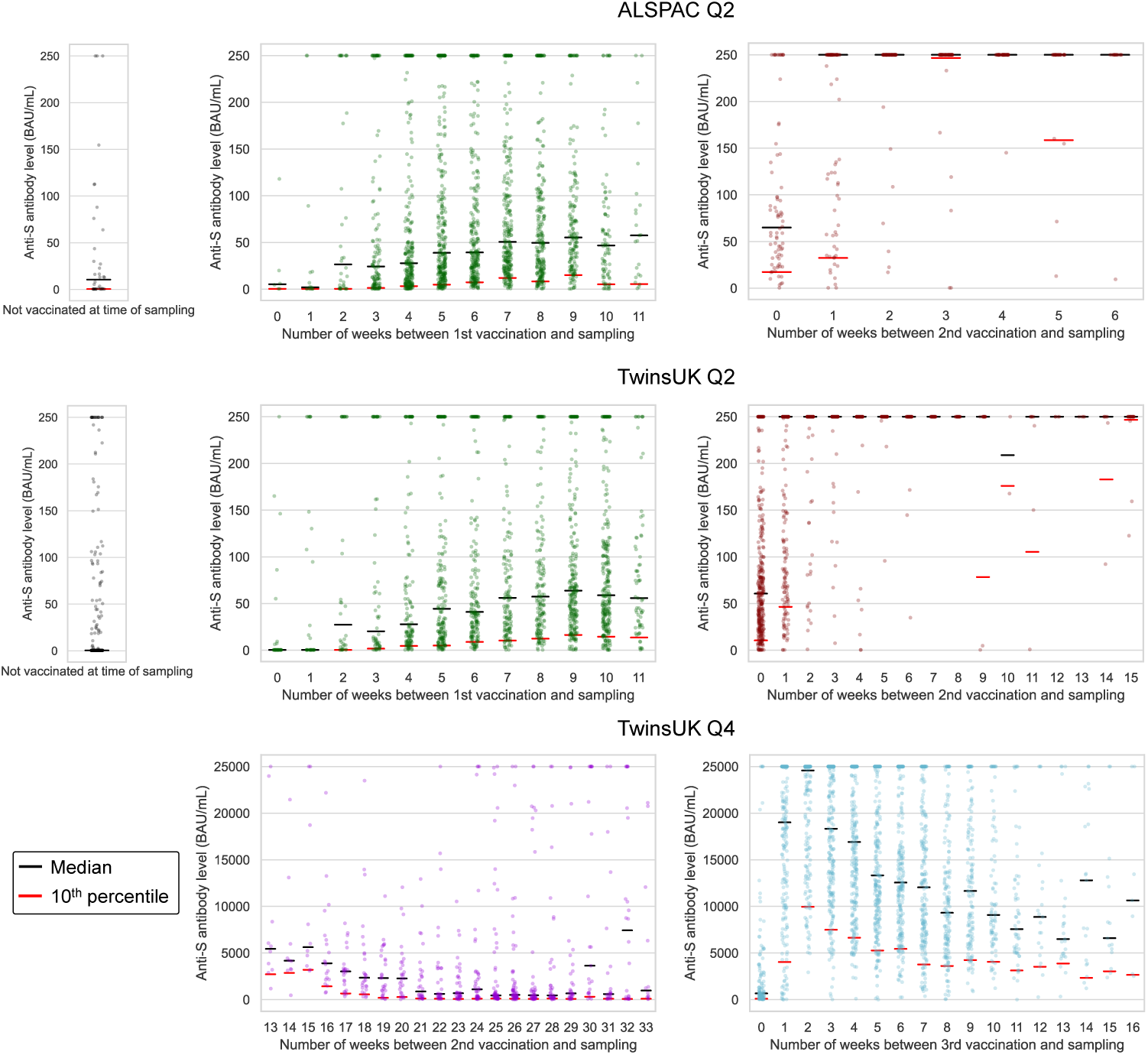
Anti-Spike antibody levels versus time since most recent vaccination, stratified by cohort and vaccination status at Q2 and Q4 antibody testing. Dot plots showing distribution of anti-Spike (anti-S) antibody levels within unvaccinated, single-, double- and triple-vaccinated individuals within ALSPAC (Q2 testing) and TwinsUK (Q2 and Q4 testing), plotted against the number of weeks since most recent vaccination at time of sampling. Black lines show median levels and red lines show 10^th^ percentile levels. The assay range is 0.4 to 250 BAU/mL for Q2 results and 0.4 to 25,000 BAU/mL for Q4 results, with a positive threshold of 0.8 BAU/mL. X-axes are limited to weeks with results for 5 or more individuals, noting TwinsUK Q4 second vaccination sub-plot begins at 13 weeks since vaccination.

From Q2 results, antibody levels peaked at nine weeks after first vaccination in both TwinsUK and ALSPAC. After second vaccination, median levels breached the 250 BAU/mL assay limit from two weeks onwards, precluding further time assessment.

### Factors associated with recorded post-vaccination infection in TwinsUK

Given the large variability in antibody response after first vaccination (Figure 1), we investigated whether a lower antibody response after first vaccination associated with post-vaccination (‘breakthrough’) infection, as evidenced by self-report (suspected or confirmed case) and/or serological testing (positive anti-Nucleocapsid test after vaccination). Within TwinsUK, post-vaccination SARS CoV-2 infection (between first vaccination and Q4 testing) was recorded in 276 of 2,993 (9.2%) individuals (further details related to post-vaccination infection given in Table S 6). Of those single-vaccinated at Q2 testing, individuals who subsequently recorded a post-vaccination infection had lower median Q2 anti-Spike antibody levels versus those who did not (median = 40 BAU/mL [IQR: 14-82 BAU/mL], n = 105, versus median = 57 BAU/mL [IQR: 24-129 BAU/mL], n = 1,057, p = 0.001), with a clear trend in post-vaccination infection incidence with Q2 antibody level quintile (Figure 3).

**Figure 3.**
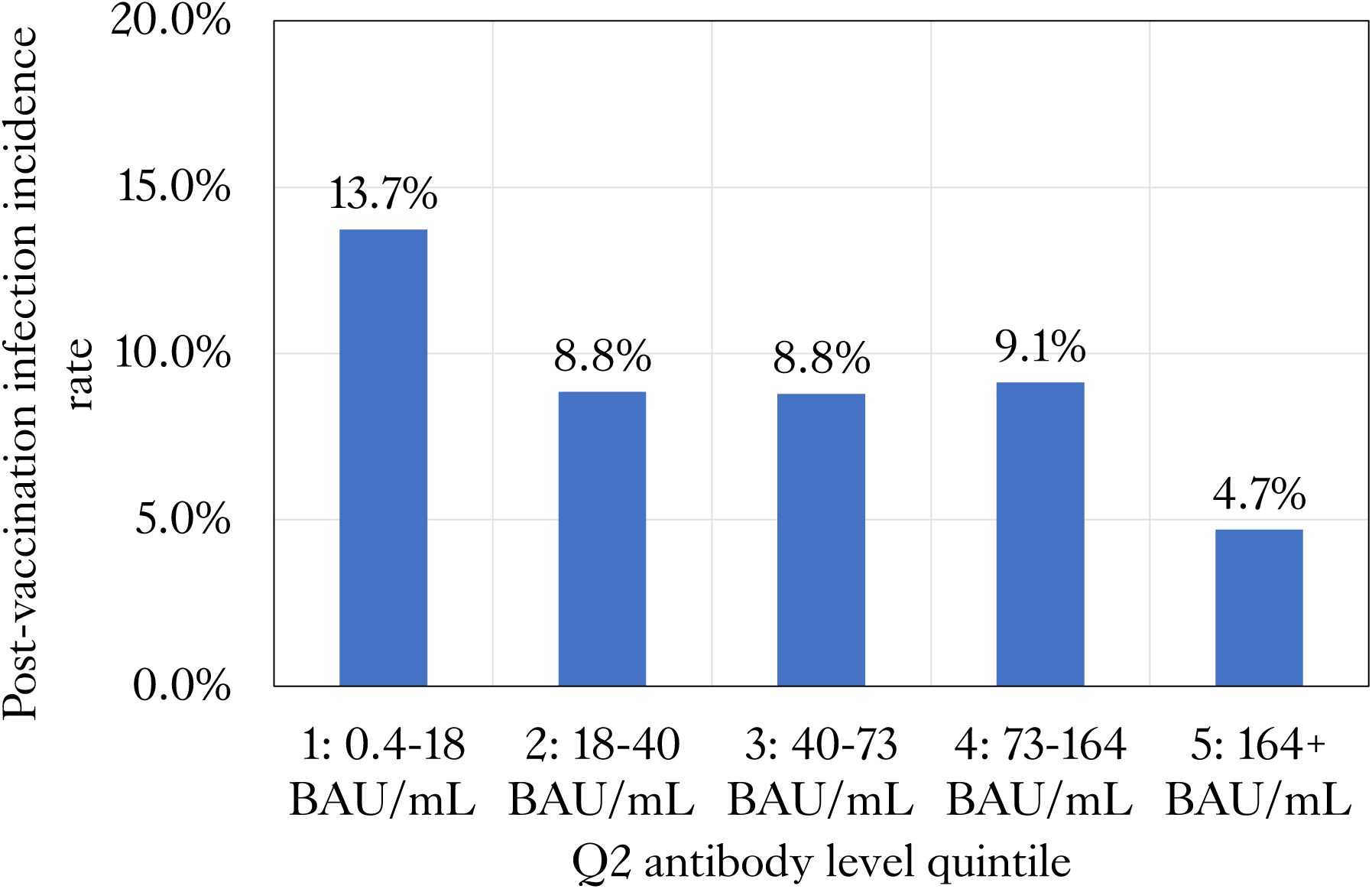
Post-vaccination infection incidence versus anti-Spike antibody level in TwinsUK. Incidence of post-vaccination infection between Q2 and Q4 antibody testing within TwinsUK individuals single-vaccinated at Q2, split by anti-Spike antibody level quintile among all single-vaccinated individuals tested at Q2.

From univariable logistic regression, vaccinated individuals with antibody levels in the lowest quintile at Q2 (range: 0.4-18.1 BAU/mL) had greater odds of post-vaccination infection (OR = 3.2 [95% CI: 1.6-6.6], p = 0.009), compared to those in the top quintile, (range: > 165 BAU/mL) (Figure 4 and Table 3, extended models presented in Table S 7). This association persisted after inclusion of weeks since first vaccination (OR = 2.9 [95% CI: 1.4-5.9], p = 0.03); and weeks since first vaccination, age, and sex (OR = 2.9 [95% CI: 1.4-6.0], p = 0.02) in multivariable models.

**Figure 4.**
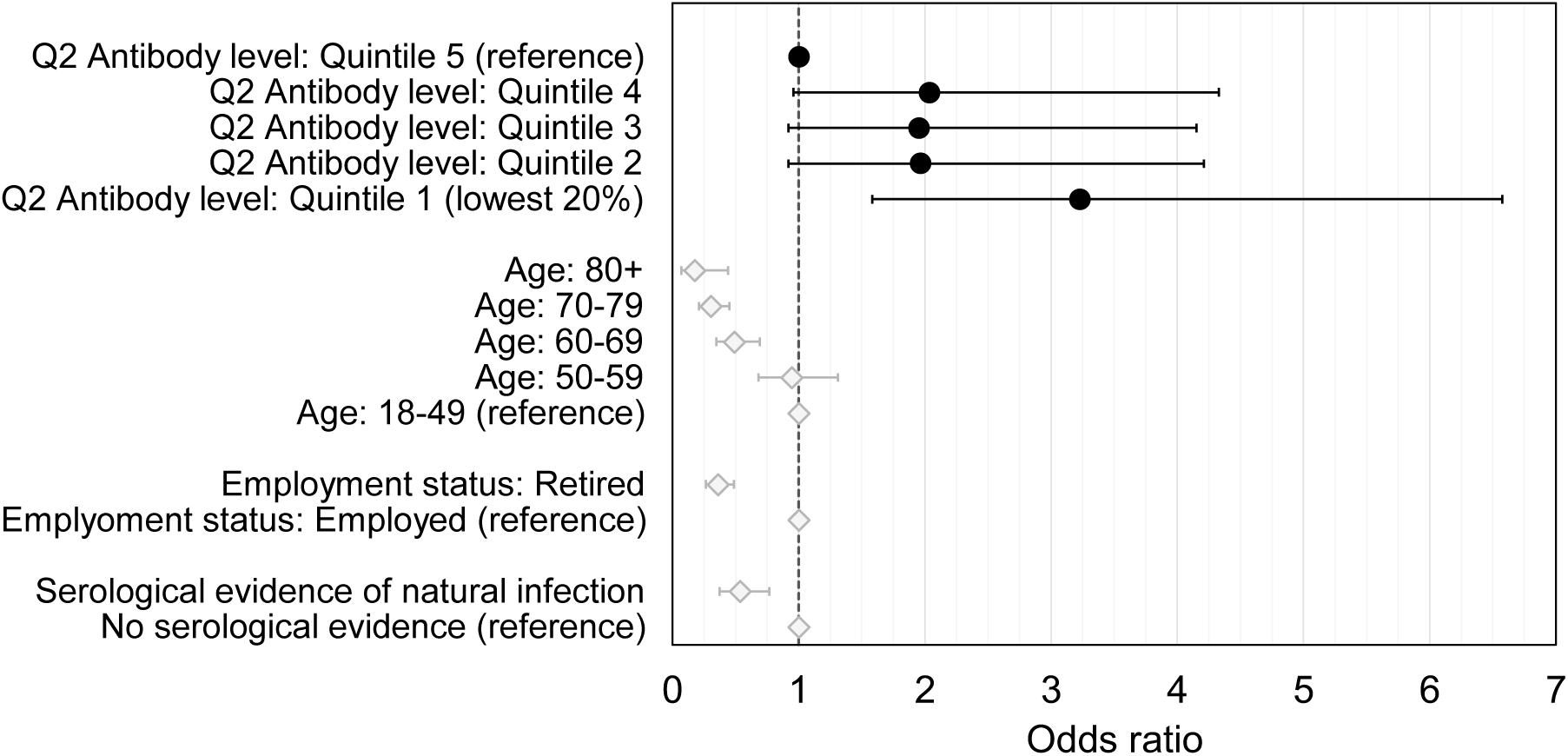
Associations with post-vaccination infection within TwinsUK. Odds ratios with unadjusted 95% confidence intervals for unadjusted, univariate logistic regression models, testing associations between post-vaccination infection and Q2 antibody levels (black circles), and selected socio-demographic and COVID-19 infection variables with evidence of association (grey diamonds) within TwinsUK. Odds ratio = 1 is indicated with a dashed black line.

**Table 3.**
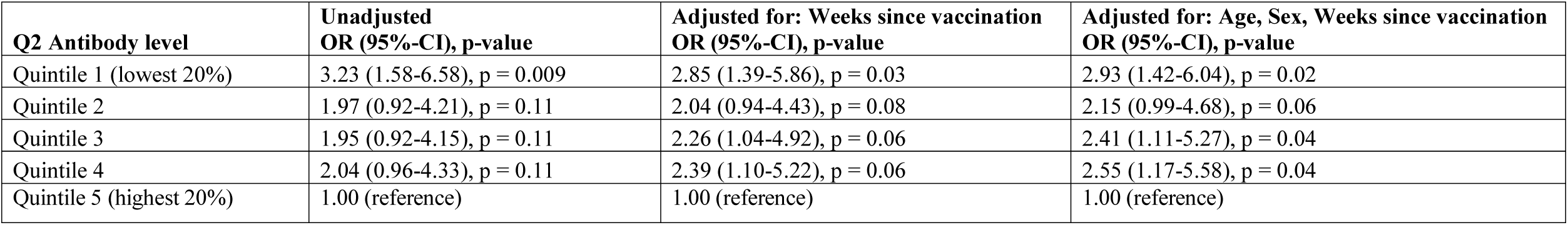
Association between post-vaccination infection and anti-Spike antibody levels within TwinsUK. Logistic regression model results, testing association between post-vaccination infection and Q2 anti-Spike antibody levels in single-vaccinated individuals within TwinsUK. Reference category was a Q2 Antibody level in quintile 5 (highest 20%). Results present odds ratios, unadjusted 95% confidence intervals, and p-values adjusted for multiple testing.

| **Q2 Antibody level** | **Unadjusted**<br>**OR (95%-CI), p-value** | **Adjusted for: Weeks since vaccination**<br>**OR (95%-CI), p-value** | **Adjusted for: Age, Sex, Weeks since vaccination**<br>**OR (95%-CI), p-value** |
| --- | --- | --- | --- |
| Quintile 1 (lowest 20%) | 3.23 (1.58-6.58), p = 0.009 | 2.85 (1.39-5.86), p = 0.03 | 2.93 (1.42-6.04), p = 0.02 |
| Quintile 2 | 1.97 (0.92-4.21), p = 0.11 | 2.04 (0.94-4.43), p = 0.08 | 2.15 (0.99-4.68), p = 0.06 |
| Quintile 3 | 1.95 (0.92-4.15), p = 0.11 | 2.26 (1.04-4.92), p = 0.06 | 2.41 (1.11-5.27), p = 0.04 |
| Quintile 4 | 2.04 (0.96-4.33), p = 0.11 | 2.39 (1.10-5.22), p = 0.06 | 2.55 (1.17-5.58), p = 0.04 |
| Quintile 5 (highest 20%) | 1.00 (reference) | 1.00 (reference) | 1.00 (reference) |

In addition to the association between post-vaccination infection and antibody levels, associations with post-vaccination infection were also found for socio-demographic and COVID-19 infection variables in univariable and multivariable logistic regression models (Figure 4 and Table S 8). Odds of post-vaccination infection decreased sharply with increasing age (e.g., 80+ versus 18-49, OR = 0.18 [95% CI: 0.07-0.44], p = 0.002), while infection was less likely for those with serological evidence of SARS-CoV-2 infection prior to Q2 testing versus those without (OR = 0.53 [95% CI: 0.37-0.77], p = 0.006), and for those who were retired versus employed (OR = 0.36 [95% CI: 0.27-0.49], p < 0.0001).

We also tested association between antibody response after second vaccination and subsequent post-vaccination infection, from which odds ratios > 1 were observed for double-vaccinated individuals at Q2 whose antibody levels did not exceed upper limit of assay (i.e., < 250 BAU/mL) compared with those whose did (250+ BAU/mL) (Table S 7). But as mentioned, analysis was constrained by the assay threshold.

### Factors associated with lower antibody levels within TwinsUK and ALSPAC

After observing associations between post-vaccination infection and lower antibody levels after first vaccination, we tested for associations with having lower antibody levels after each round of vaccination. Increased odds of lower antibody levels (defined as lowest 5%, 8% or 10% within each group stratified by vaccination status) were consistently observed across multiple vaccination rounds in both TwinsUK and ALSPAC (Figure 5) for the follow health-related variables:

a. those advised as being on the UK “Shielded Patient List” [36,37]. For example, for lowest 10% after first vaccination, TwinsUK: (OR = 4.0, [95% CI: 2.2-7.4], p = 0.0001), ALSPAC: (OR = 4.1, [95% CI: 1.8-9.5], p = 0.02);
b. those with poorer self-rated health. For example, for lowest 10% after first vaccination in TwinsUK: (OR = 1.4, [95% CI: 1.1-1.6], p = 0.02), for lowest 5% after first vaccination in ALSPAC: (OR = 1.4, [95% CI: 1.1-1.8], p = 0.08), for a -1 step on an ordinal 1-5 (poor-excellent) scale;
c. those with indicators of immunosuppression. For example, for lowest 10% after second vaccination in TwinsUK: (OR = 4.2, [95% CI: 1.9-9.5], p = 0.006), for lowest 10% after first vaccination in ALSPAC: (OR = 6.2, [95% CI: 2.7-14.5], p = 0.001).

**Figure 5.**
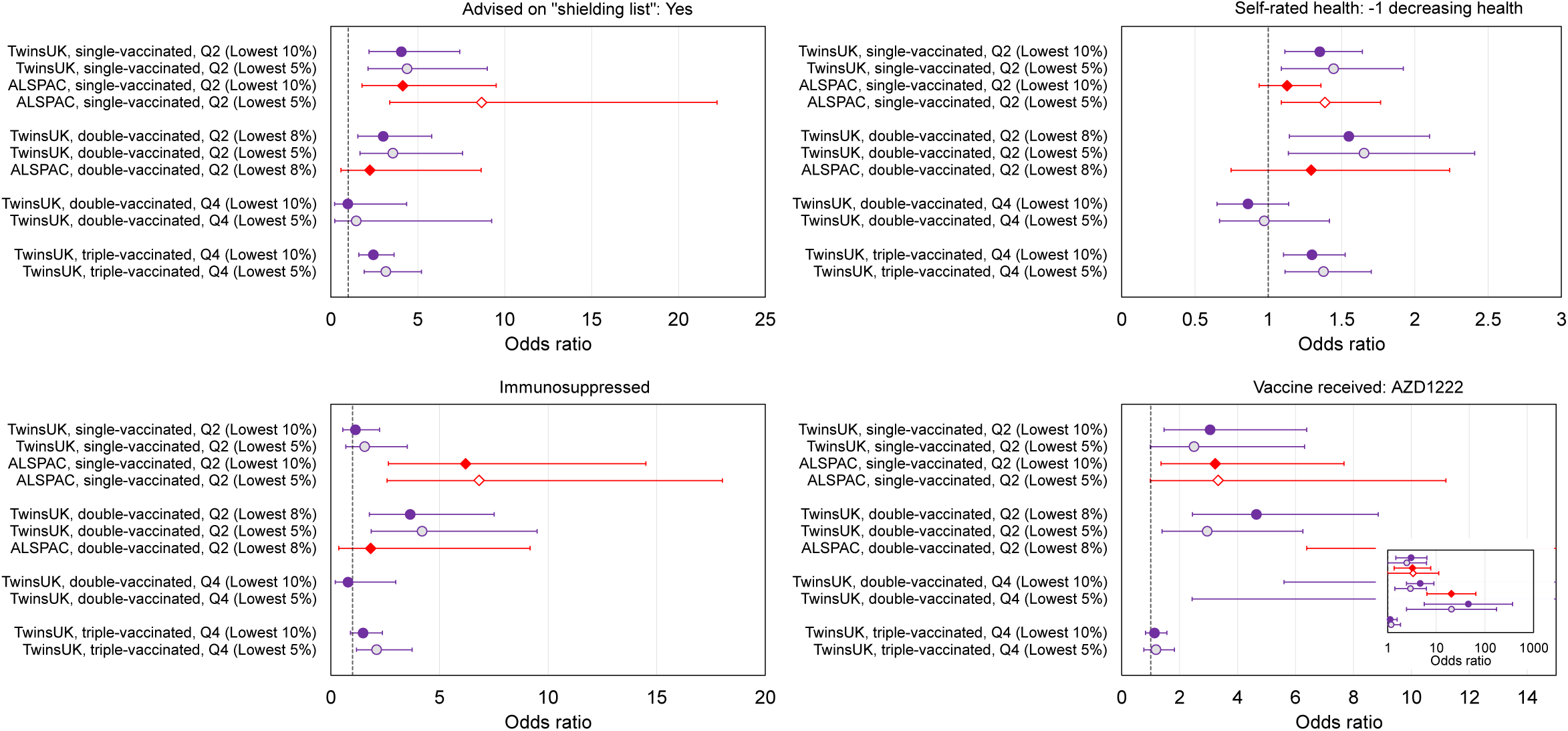
Associations with low relative anti-Spike antibody levels within TwinsUK and ALSPAC. Odds ratios with unadjusted 95% confidence intervals for selected variables, testing associations with low anti-Spike antibody levels (lowest 5%, 8% or 10%), for TwinsUK (purple circles) and ALSPAC (red diamonds) individuals tested in Q2 or Q4, after first, second and third vaccination. Each point estimate originates from a distinct multivariate logistic regression model, including the variable of interest and adjustment variables of age, sex, name of most recent vaccine received and weeks since most recent vaccination. Note x-axis ranges on subplots vary. Odds ratio = 1 is indicated with a dashed black line. Vaccine received: AZD1222 panel inset presents results on a logarithmic x-axis.

Results for all exposure variables are presented Table S 9 and further visualisations in Figure S 7 to Figure S 13.

Individuals in both cohorts who received the AZD1222 (Oxford/AstraZeneca) vaccine versus BNT162b2 (Pfizer BioNTech) were more likely to have lower antibody levels after first vaccination (for lowest 10% in TwinsUK: (OR = 3.1, [95% CI: 1.5-6.4], p = 0.02), and ALSPAC: (OR = 3.2, [95% CI: 1.4-7.7], p = 0.09)), and second vaccination (for lowest 8% in TwinsUK Q2: (OR = 3.0, [95% CI: 1.4-6.2], p = 0.03), TwinsUK Q4: (OR = 45.7, [95% CI: 5.6-372], p = 0.001), and ALSPAC: (OR = 20.3, [95% CI: 6.4-64.7], p = 0.0001)). However, receiving AZD1222 at second vaccination was not associated with lower antibody levels after third vaccination in TwinsUK (for lowest 10%, (OR = 1.1, [95% CI: 0.8-1.6], p = 0.8)). Those with longer time since vaccination at time of sampling had increased odds of lower antibody levels after second and third vaccination, while individuals sampled later after first vaccination had decreased odds of lower antibody levels. Lower likelihood of lower antibody levels was seen across multiple rounds of vaccination within TwinsUK for those with evidence of SARS-CoV-2 infection prior to antibody testing, either through serological testing (e.g., outcome: lowest 10% after third vaccination (OR = 0.45, [95% CI: 0.28-0.71], p = 0.004]) or self-reported confirmed cases (e.g., outcome: lowest 10% after third vaccination (OR = 0.25, [95% CI: 0.13-0.45], p = 0.0001)), but not for self-reported suspected cases. Several logistic regression results testing association with positive anti-Nucleocapsid antibodies were either suppressed or not feasible to run, due to disproportionately low numbers of seropositive individuals having lower antibody levels in both TwinsUK and ALSPAC – further suggesting association between evidence of infection and increased antibody levels.

Less consistent associations (i.e., not observed across more than one round of vaccination) with increased likelihood of lower antibody levels were seen in TwinsUK for several other variables: very frail, high multimorbidity (3 or more of 5 selected comorbidities), rheumatoid arthritis, employment status of permanently or long-term sick or disabled, and lower educational attainment (Table S 9). No clear associations with lower antibody levels were seen with age, sex, or BMI in either TwinsUK or ALSPAC (Figure S 14).

### Twin-pair analysis in TwinsUK after third vaccination

Within TwinsUK, pairs of identical monozygotic (MZ) twins showed smaller average intra-pair anti-Spike antibody level differences after third vaccination versus non-identical dizygotic (DZ) twin-pairs (median twin-pair difference = 5,000 BAU/mL versus 6,800 BAU/mL, p = 0.0002 for MZ versus DZ), while differences between pairs of non-related individuals were largest (median difference = 7,900 BAU/mL, p < 0.0001 for MZ versus non-related) (Table S 10 and Figure S 15).

Generalised linear mixed effects regression models of MZ and DZ twin pairs were performed with anti-Spike antibody levels after third vaccination as the dependent variable, to further test the persistence of, and genetic and environmental contributions to, associations with antibody levels. Within MZ twin-pairs discordant for “Shielded Patient List” status or rheumatoid arthritis, twins on the “Shielded Patient List” (within-pair regression coefficient: -3,700 BAU/mL, [95% CI: -6,500, - 880 BAU/mL], p = 0.01) and twins with rheumatoid arthritis (within-pair regression coefficient: - 5,800 BAU/mL, [95% CI: -11,200, -420 BAU/mL], p = 0.03) had lower antibody levels after third vaccination than their co-twin (Table S 11). Evidence of between-pair associations was also observed for self-rated health, frailty index, and highest educational attainment.

## Discussion

In this study we used SARS-CoV-2 anti-Nucleocapsid and anti-Spike antibody testing, and questionnaire data collected at multiple time points during and before the COVID-19 pandemic, to investigate associations with antibody response to vaccination in TwinsUK and ALSPAC longitudinal population-based cohorts.

Firstly, we observed large variability in the magnitude of antibody response to first vaccination, with decreasing variability after second and third vaccinations. Secondly, individuals with lower levels of anti-Spike antibodies following first vaccination were at higher risk of future SARS-CoV-2 infection at any subsequent time, including after further vaccinations. Acknowledging that anti-Spike antibodies are only one component of the complex immune response to vaccination [38], our data supports the contention that anti-Spike antibody level measurement is a useful proxy indicator of overall immune response. Thirdly, individuals who: were on the UK “Shielded Patient List”; had lower self-rated health; received AZD1222 (Oxford/AstraZeneca) vaccine for first and second vaccination; were sampled at longer time since second vaccination and third vaccination; were prescribed immunosuppressant medication (in TwinsUK) or with self-reported immunosuppression (in ALSPAC), all had higher odds of lower antibody levels following vaccination. These findings were consistent across multiple rounds of vaccination and in both cohorts. Individuals with evidence of SARS-CoV-2 infection prior to sampling were less likely to have lower antibody levels, consistent with previous studies that postulating that the quantity and quality of antibody response were linked to the total number of exposures to SARS-CoV-2 [39,40]. Fourthly and finally, in analyses exploiting the twin-pair design of one cohort, we found that genetic factors influenced antibody level variation (considered only after third vaccination), with smaller differences in antibody levels within genetically identical MZ pairs compared with DZ pairs. Twin-pair regression models showed that genetic factors may contribute to the associations between self-rated health, frailty and highest educational attainment and antibody response following vaccination. In contrast, models showed that association between antibody levels and “Shielded Patient List” status was independent of such genetic and other shared factors, after explicit adjustment for key vaccination and infection variables. Altogether, our data support the usefulness of the UK “Shielded Patient List” criteria as a measure of COVID-19-related risk, even after UK government guidance regarding shielding was ended in April 2021 [41].

In our regression analyses, there were some patterns of note. Health-related variables associated with lower antibody levels were more often general (self-rated poor health, immunosuppression indicators) and/or collective measures with wide-ranging criteria (e.g., “Shielded Patient List”, very frail, multimorbidity), rather than specific factors such as individual comorbidities (e.g., rheumatoid arthritis). Associations were also found with some socio-demographic variables, and with COVID-19 infection and vaccination variables, suggesting that variation in post-vaccination antibody levels between individuals may originate from a wide range of variables *in combination*. This exposes a limitation to our analytical approach, as we have limited scale and power to examine the intersection of likely risk factors in combination. Further, of the several variables associated with antibody levels, only serology-based evidence of prior SARS-CoV-2 infection was directly associated (here, negatively associated) with subsequent post-vaccination infection between April-May 2021 and November 2021-January 2022 (with the majority sampled before the peak of the current Omicron wave). We found no consistent associations of lower antibody levels with age or employment status, but a very strong age gradient (lower incidence with older age) and lower likelihood among retired (vs. employed) individuals of post-vaccination infection. These results again are consistent with risk of infection being a complex combination of SARS-CoV-2 case prevalence, individual immune response to vaccination, and individual level of exposure (i.e., behaviour). Such aspects often oppose each other by design (for example, higher shielding behaviours by more elderly individuals). Nevertheless, given the ongoing relaxation of measures across many countries, groups previously less exposed may become more at risk. Consequently, our findings regarding factors associated with post-vaccination infection may change over time, to align more closely with variables identified as associated with lower antibody levels.

Longitudinal antibody testing within TwinsUK at Q4 highlighted the effectiveness of third vaccination at both generating high absolute levels of antibodies and reducing variability in post-vaccination antibody levels evident after earlier doses. Even among sub-groups associated with having the lowest antibody levels and/or higher risk of severe COVID-19 (such as shielding, frail, and/or immunosuppressed individuals), 90-95% recorded absolute anti-Spike levels of 1,000 BAU/mL or higher (Table S 5) – a level similar to the median level (1,300 BAU/mL) of all individuals tested at 28-34 weeks after second vaccination, and levels (900 BAU/mL) previously associated with a vaccine efficacy of 90% after second vaccination by the Oxford COVID Vaccine Trial Group (prior to the emergence of Delta and Omicron variants) [12]. These results are consistent with other studies – for example, kidney transplant patients with poor responses to first and second vaccination had improved serological responses after third vaccination [24]. Moreover, although individuals receiving AZD1222 vaccine (versus BNT162b2 [Pfizer BioNTech]) were more likely to have lower antibody levels after first and second vaccination, these differences were no longer evident after third vaccination. Our finding is also consistent with lower vaccine effectiveness estimates and increased post-vaccination infection incidence after first or second vaccination following AZD1222 versus BNT162b2 [15,42,43], but only minor differences after third vaccination [15,44].

The cross-sectional nature of antibody testing and varied age profile of TwinsUK meant that antibody waning at longer time since second vaccination (sampled at 28-34 weeks on average) within a population-based study could be assessed, in larger numbers than comparable studies that tested health and social care workers [9,45]. Our study highlights the extent of the antibody boost received after third vaccination, while our characterisation of SARS-CoV-2 infection and vaccination within our cohorts illustrates the immunological resilience of the UK population, potentially relevant for future waves of infection. The following findings are valuable for epidemiological modelling of infection rates and hospitalisations: age gradients observed in anti-Nucleocapsid and anti-Spike seropositivity across 11 UK longitudinal studies at Q2 (Figure S 5), and within TwinsUK at Q4 the observed variation in infection prevalence estimates across key socio-demographic variables and comparison of self-reported and serology-based infection measures (Figure S 4).

We also acknowledge limitations of this work. Firstly, it is unclear to what extent higher anti-Spike antibody levels after third vaccination translate into increased protection against Omicron and other emerging variants, noting vaccination-induced antibodies appear less able to neutralise newer variants compared with wild-type [17,18]. Concurrently, waning antibody levels are not necessarily indicative of declining neutralisation ability of antibodies or of cell-mediated immunity [46].

Therefore, further work will be needed to update this study and previous other studies that have correlated anti-Spike antibody levels with risk of SARS-CoV-2 infection [12], as well as further work to develop understanding of the relationship between humoral and cellular immunity [47]. Secondly, both TwinsUK and ALSPAC (Generation 0) participants are disproportionately older, female, and more likely of white ethnicity, in comparison to the UK population. Geographically, TwinsUK (based in London) is skewed towards lower deprivation areas in south east England and ALSPAC (based in Bristol) towards south west England. Consequently, our ability to detect associations with smaller effect sizes, and within groups that here are of small sample size (such as individuals of other than white ethnicities, and individuals living in higher deprivation areas) is limited. Timely availability of SARS-CoV-2 vaccination data also limited our ability to perform regression analyses in the nine other UK longitudinal studies for which antibody testing was undertaken concurrently with TwinsUK and ALSPAC at Q2. As well as general replication of analyses herein, the varied cohort characteristics captured within this total dataset would have particularly benefitted testing of associations with socio-demographic groups underrepresented in TwinsUK and ALSPAC. Thirdly, our analyses are subject to selection biases due to use of multiple and varying data collections that rely on voluntary participation. As a result, the participants with available data for all variables herein may not be representative of the wider cohorts, which may cause collider bias and affect findings as outlined elsewhere [48,49]. For example, indicators of poorer health have been associated with lower response to COVID-19 questionnaires in ALSPAC [50], which may bias the observed results. Acknowledging the potential effects of biases, the replication of multiple associations with lower antibody levels across compositionally-varied TwinsUK and ALSPAC cohorts and across multiple rounds of vaccination support the robustness of our findings. It is these replicated findings that we chose to discuss primarily.

In conclusion, our results suggest measurement of anti-Spike antibodies generated in response to first SARS-CoV-2 vaccination may have potential use as an early indicator to identify individuals at higher risk of a future SARS-CoV-2 infection, particularly in the many countries where vaccination roll-out is at an earlier stage. Individuals who previously met UK “Shielded Patient List” criteria had consistently lower antibody responses to vaccination than other participants, highlighting the importance of continuing to inform such individuals of their personal risk of SARS-CoV-2 infection, despite the UK government decision to end shielding guidance in April 2021 [41]. This result should inform prioritisation of vaccination towards these individuals in any future immunisation campaigns.

## Methods

### Study participants

TwinsUK is a UK-based national registry of monozygotic and dizygotic twins, with over 15,000 twins registered since 1992 [33].

ALSPAC is a prospective population-based cohort of pregnant women with expected delivery dates between April 1991 and December 1992 who lived in Bristol, UK and the nearby surrounding area; with follow-up of these women and their partners (collectively known as Generation 0, G0), and their children (Generation 1, G1), ever since [34,35]. The initial cohort consisted of 14,541 pregnancies, with 13,988 children alive at one year, and was later expanded when children were approximately age 7, to give a total of 15,454 pregnancies, with 14,901 children alive at one year. Analyses herein were carried out solely with G0 participants due to low rates of vaccination among the G1 children generation at the time of initial serology.

During the COVID-19 pandemic, participants from both cohorts were invited to complete cohort-specific questionnaires and to submit blood samples via post for SARS-CoV-2 antibody testing. In the first round of coordinated testing in TwinsUK and ALSPAC, participants submitted samples in April and May 2021. This first testing round is referred to throughout as Q2 testing (from calendar year quarter 2 start date). Participants of TwinsUK were later invited for a second round of antibody testing with the same assay, with samples collected from November 2021 to January 2022. This round of antibody testing is referred to throughout as Q4 testing (from quarter 4 start date). Further details of COVID-19 questionnaires and antibody testing are given in following sections.

Flow diagrams of participants identified for analyses based on available data are shown in Figure S 1 and Figure S 2 for TwinsUK and Figure S 3 for ALSPAC. In brief, individuals with unknown vaccination status at time of antibody testing were excluded from all analyses. For descriptive analysis of antibody levels versus time since vaccination, all individuals with known vaccination status were included. For analysis of variables associated with low antibody levels, individuals sampled fewer than 28 days since first vaccination, or fewer than 14 days since second or third vaccination, were excluded (these thresholds were chosen to allow sufficient time for an immunological response after each vaccine dose, based on previous studies [1,2]), while individuals with 77 days or more since first vaccination were excluded in case of misclassification due to unreported further vaccination (based on 11-12 week spacing between doses for majority of adults in the UK). In addition to the above criteria, for analysis of variables associated with post-vaccination infection within TwinsUK, individuals must have participated in Q2 antibody testing followed by either Q4 antibody testing and/or concurrent COVID-19 questionnaire.

### Questionnaires administered during the COVID-19 pandemic

TwinsUK COVID-19 questionnaires were administered in April-May 2020 [51], July-August 2020, October-December 2020, April-July 2021 (approximating first round of antibody testing, Q2), and November 2021-February 2022 (approximating second round of antibody testing, Q4). ALSPAC COVID-19 questionnaires were administered in April-May 2020 [52], May-July 2020 [53], October 2020 [54], November 2020-March 2021 (approximating first round of antibody testing, Q2) [55], and July-December 2021.

Details of variables collected through cohort-specific pandemic questionnaires are provided in Table S 1. Questions included self-reported SARS-CoV-2 infection and symptoms, results of SARS-CoV-2 testing, and vaccination status (date, dose number, manufacturer/brand) once the UK SARS-CoV-2 vaccination programme had commenced (8 December 2020). Questions made no distinction between pre-planned third vaccination for high-risk individuals and third vaccination given as part of the wider community ‘booster’ campaign – as such we refer to third vaccination or triple-vaccinated individuals throughout. By virtue of the national vaccination roll-out policy (tiered by age and at-risk status), at Q2 participants may have received nought, one, or two vaccination doses; by Q4 some individuals had received a third dose.

As questionnaires were cohort-specific, assessed variables were not completely uniform (both question wording and collected data). Details for comparison are shown in Table S 1.

### SARS-CoV-2 antibody testing

Q2 testing in TwinsUK and ALSPAC occurred along with an additional nine UK-based longitudinal studies who collected samples in unison as part of the UK National Core Studies Longitudinal Health & Wellbeing (NCS-LH&W) programme [56]. Additional cohort-specific details and results for ALSPAC and Extended Cohort for E-health, Environment and DNA (EXCEED) are provided elsewhere [57,58]. Data availability in cohorts other than TwinsUK and ALSPAC limited analysis to presentation of overall seropositivity and variation with cohort mean age.

For TwinsUK antibody testing in Q2 and Q4, invitation criteria were based on availability of email addresses and/or completion of previous COVID-19 questionnaires. ALSPAC invitation criteria are given in detail elsewhere [58]. For both cohorts, participants received fingerprick blood sample collection kits via post. Blood sample collection was self-administered. Samples were sent via post to either Pura Diagnostics or Eurofins County Pathology (partner laboratories of Thriva Ltd), who assayed samples and shared results with TwinsUK and ALSPAC. Quantitative IgG anti-Spike SARS-CoV-2 antibody levels and qualitative IgG anti-Nucleocapsid antibody status were assayed using CE-marked capillary blood Roche Elecsys Anti-SARS-CoV-2 immunoassays [59]. Quantitative anti-Spike results were given in units per millilitre (U/mL), with a quantitative range of 0.4-250 U/mL for Q2 testing. For Q4 testing, samples were diluted to give an expanded quantitative range of 0.4-25,000 U/mL, allowing quantitative discernment for higher levels at this timepoint. Tests had a positive threshold of 0.8 U/mL. 1 U/ml is equivalent to 1 unit of WHO standardised unit, binding antibody units per millilitre (BAU/mL) (WHO international standard: 20/136 [60]). Thus, we have quoted results in BAU/mL to aid comparison across studies. Anti-Nucleocapsid results were qualitative, with a positive result for a value greater than a cut-off unit = 1.

Additional antibody testing was also undertaken in-house for TwinsUK samples between April 2020 and April 2021. Quantitative enzyme-linked immunosorbent (ELISA) assays testing anti-Nucleocapsid and anti-Spike antibody levels were performed using previously published methods [4]. These data were used to determine serology-based infection status prior to Q2 antibody testing.

### Identification of SARS-CoV-2 infection

#### Assessment of prior SARS-CoV-2 infection, at time of antibody testing

Prior SARS-CoV-2 infection was classified with three distinct variables derived from self-reported questionnaire data or serological testing.

1. “SARS-CoV-2 infection status (self-reported)”: derived solely from self-reported COVID-19 infection and testing questionnaire data. The classification was primarily derived from responses to “Do you think that you have or have had COVID-19?” in prior questionnaires. Classification options are given below:
  a. Confirmed case: “Yes, confirmed by a positive test”.
  b. Suspected case: “Yes, suspected by a doctor but not tested”.
  c. Suspected case: “Yes, my own suspicions”.
  d. Unsure (TwinsUK only): “Unsure”.
  e. No: “No”.
  In TwinsUK questionnaires only, individuals were also asked to self-report any positive COVID-19 tests. Infection status of individuals who self-reported a positive test was classified as a confirmed case, irrespective of their answer to the question above.
2. “SARS-CoV-2 infection status (serology-based)”: derived from laboratory serological testing (Q2 [TwinsUK and ALSPAC], Q4 [TwinsUK only] and/or other within-cohort testing [TwinsUK only]), informed by self-reported vaccination status. We followed Centers for Disease Control and Prevention guidance on interpretation of anti-Nucleocapsid and anti-Spike results while accounting for vaccination status [61] as follows:
  a. Evidence of SARS-CoV-2 infection: A positive anti-Nucleocapsid result at any time or a positive anti-Spike result prior to vaccination.
  b. No evidence of SARS-CoV-2 infection: Negative anti-Nucleocapsid and anti-Spike result prior to vaccination, or negative anti-Nucleocapsid and positive anti-Spike result following vaccination (anti-Spike antibody assumed to be generated by vaccination).
3. “Anti-Nucleocapsid antibody status”: derived solely from laboratory serological testing (from Q2 or Q4 testing only). The classification was as follows:
  a. Positive: Positive anti-Nucleocapsid test result at Q2 or Q4 testing.
  b. Negative: Negative anti-Nucleocapsid test result at Q2 or Q4 testing.

From these variables, distinct measures of the proportion of individuals with evidence of prior SARS-CoV-2 infection, or “natural infection”, at time of Q2 and Q4 testing were quantified within both cohorts.

Thus, “SARS-CoV-2 infection status (self-reported)” and “SARS-CoV-2 infection status (serology-based)” variables identify individuals with any history of SARS-CoV-2 infection (who are not necessarily seropositive for anti-Nucleocapsid antibodies at time of testing), while “Anti-Nucleocapsid antibody status” assesses the contemporaneous level of infection-induced antibody response.

#### Assessment of post-vaccination SARS-CoV-2 infection

For analysis of variables associated with post-vaccination SARS-CoV-2 infection (performed within TwinsUK only), individuals with post-vaccination SARS-CoV-2 infections were identified using the following criteria:

1. A ‘suspected case’ or ‘confirmed case’ from “SARS-CoV-2 infection status (self-reported)” variable at Q4 testing, with symptoms commencing after first vaccination. Infection and vaccination dates obtained from COVID-19 questionnaires.
2. A ‘confirmed case’ from “SARS-CoV-2 infection status (self-reported)” variable at Q4 testing, with a self-reported positive antigen test dated after first vaccination. Infection and vaccination dates obtained from COVID-19 questionnaires.
3. A positive SARS-CoV-2 anti-Nucleocapsid result at Q4 testing after previous negative anti-Nucleocapsid results up to and including Q2, for individuals vaccinated at least once at Q2. The approximate date of infection is unknown for individuals who meet this criterion only.

Individuals meeting one or more of these criteria were considered as having post-vaccination infection. Individuals who did not meet any of these criteria were considered as controls (i.e., no post-vaccination infection). Individuals must have participated in TwinsUK Q4 antibody testing and/or concurrent COVID-19 questionnaire for post-vaccination infection to be determinable and for inclusion as controls or cases.

### Phenotypic data list

Variables from antibody testing and pandemic questionnaire data were supplemented with pre-pandemic socio-demographic and health variables for TwinsUK and ALSPAC analyses (details in Table S 1). A full list of variables considered in analyses is given in Table 1.

### Statistical analyses

#### Descriptive analysis of antibody levels after first, second, and third vaccination

Median, 10^th^ and 5^th^ percentile antibody levels were produced for univariate splits of adjustment and exposure variables listed in Table 1. Differences in median antibody levels (per Results) were tested using a two-sided Mann-Whitney U-test [72]. Trend in median antibody level versus number of weeks post-vaccination was tested using the Mann-Kendall trend test [73,74].

#### Association between SARS-CoV-2 infection and socio-demographic variables

Associations between SARS-CoV-2 infection, quantified from SARS-CoV-2 infection status (self-reported), SARS-CoV-2 infection status (serology-based), and Anti-Nucleocapsid antibody status, and age, sex, ethnicity, local area deprivation and rural-urban classification were tested using the chi-square test of independence.

#### Logistic regression analyses

Within TwinsUK only, univariable and multivariable logistic regression were used to test associations between an outcome of post-vaccination SARS-CoV-2 infection and exposure variables related to: Q2 anti-Spike antibody levels; socio-demographics; COVID-19 infection; COVID-19 vaccination. In TwinsUK and ALSPAC, multivariable logistic regression was also performed to test associations between the outcome of low anti-Spike antibody levels (as defined below) after each round of vaccination (after first and second vaccinations for both TwinsUK and ALSPAC, and after third vaccination for TwinsUK only) and all exposure variables previously listed.

Each model included the outcome variable, a single exposure variable of interest, and a set of adjustment variables. Individual exposure variables of interest were tested in sequence, fitting a separate logistic regression model for each combination of outcome, adjustment, and exposure variables. Only individuals with complete data for the given model were included. For each categorical variable within logistic regression models, reference categories were chosen based on the normative, modal, maximum or minimum value/category, as appropriate (reference categories given in Table S 1). Within TwinsUK models only, the HC3 estimator of logistic regression coefficient standard errors was used to account for heteroskedasticity (which biases conventional standard errors in analysis of related twin pairs [75–77]). (Two-sided) p-values were corrected for multiple testing using the Benjamini/Hochberg p-value adjustment [78].

An outcome of post-vaccination SARS-CoV-2 infection was identified using the criteria previously described. An a priori outcome of ‘low anti-Spike antibody levels’ was defined relatively within each group stratified by vaccination status (single-, double-, triple-vaccinated within TwinsUK, and single-, double-vaccinated within ALSPAC) and assigned to individuals in either the lowest 5% or lowest 10% (with a separate model for each threshold) of anti-Spike antibody levels. As such the anti-Spike threshold value used to define low levels varied between models. Most double-vaccinated individuals at Q2 testing had antibody levels above the upper assay limit of 250 BAU/mL (TwinsUK: 92%, ALSPAC: 92%). Thus, a threshold of < 250 BAU/mL was used instead of 10% to identify low antibody levels after second vaccination, corresponding to the lowest 8% in both TwinsUK and ALSPAC. Due to small sample size, models testing association in individuals with the lowest 5% of antibody levels after second vaccination were not performed in ALSPAC. In total, for each exposure variable, there were six TwinsUK models and three ALSPAC models.

Multivariable models testing association between post-vaccination SARS-CoV-2 infection and anti-Spike antibody levels used the following sets of adjustment variables: 1) number of weeks since most recent vaccination; 2) age, sex, number of weeks since most recent vaccination. Multivariable models testing association between post-vaccination SARS-CoV-2 infection and socio-demographic variables used the following sets of adjustment variables: 1) age; 2) age, SARS-CoV-2 infection status (serology-based); 3) age, sex, SARS-CoV-2 infection status (serology-based). Multivariable models testing associations with low anti-Spike antibody levels used the following set of adjustment variables: age, sex, most recent vaccine received and number of weeks since most recent vaccination. Adjustment variables were chosen based on relatively large effects observed in preliminary descriptive analysis.

#### Twin-pair analyses

To assess the relationship between zygosity and relatedness on variation in antibody levels between pairs of individuals after third vaccination within TwinsUK, antibody level differences were calculated for all pairs of monozygotic and dizygotic twins, and within all combinations of non-related pairs. Difference between the resulting median pair-differences within monozygotic, dizygotic, and non-related pairs were tested using the two-sided Mann-Whitney U-test.

For variables associated with low antibody levels (from logistic regression analyses), within-twin-pair associations with unadjusted anti-Spike antibody levels after third vaccination were tested using “within-between” generalised linear mixed effects models. Such models implicitly control for pair-specific shared genetic and environmental factors by design and are commonly used in twin-pair studies as described elsewhere [79]. The pseudonymised family identifier variable was fitted as a random effect, allowing intercept to vary for each twin-pair. For the exposure variable of interest, twin-pair mean values and difference-to-twin-pair-mean values were calculated and both included as “between-pair” and “within-pair” variables in models, respectively. Age, sex, number of weeks since third vaccination, brand of vaccine received for third vaccination, and SARS-CoV-2 infection status (serology-based) were also included in models as adjustment variables. For each exposure variable, separate models were fitted for monozygotic and dizygotic twin pairs. Differences between “between-pair” and “within-pair” coefficients were tested using a Wald test. Unpaired single twins and individuals without data for all variables were excluded from the given model.

### Software

TwinsUK analyses were performed using python v3.8.8 [80] and packages: numpy v1.20.1, pandas v1.2.4, statsmodels v0.12.2, scipy v1.6.2, scikit-learn v0.24.1, matplotlib v3.3.4, pymannkendall v1.4.2, seaborn v0.11.1. ALSPAC analyses were performed using python v3.9.7 and packages: numpy v1.20.3, pandas v1.3.4, matplotlib v3.4.3, and seaborn v0.11.2, and R v4.1.2 [81] and packages: plyr v1.8.6, dplyr v1.0.7 and broom v0.7.11.

## Data Availability

Antibody test data are available within the UK Longitudinal Linkage Collaboration upon application (see https://ukllc.ac.uk/apply/). UK LLC houses COVID-19 related datasets from over 20 UK longitudinal population studies (see https://ukllc.ac.uk/datasets/). TwinsUK data are available to researchers on application. Data access is managed by the TwinsUK Resource Executive Committee (TREC) to ensure privacy and protect against misuse (see https://twinsuk.ac.uk/resources-for-researchers/access-our-data/). ALSPAC data are available via an online proposal system (see http://www.bristol.ac.uk/media-library/sites/alspac/documents/researchers/data-access/ALSPAC_Access_Policy.pdf).

https://twinsuk.ac.uk/resources-for-researchers/access-our-data/

http://www.bristol.ac.uk/media-library/sites/alspac/documents/researchers/data-access/ALSPAC_Access_Policy.pdf

## Acknowledgements

We are grateful to participants from all studies for participating in testing and completing questionnaires during the COVID-19 pandemic and thank colleagues (TwinsUK: Gulsah Akdag, Andy Anastasiou, Julia Brown, Rachel Horsfall, Genevieve Lachance, Ayrun Nessa, Timothy Spector, Dovile Vaitkute, Sam Wadge, Sivasubramaniam Rajan Wignarajah, Darioush Yarand. ALSPAC: Amy-Louise Gordon, Alix Groom, Nicholas Wells, Hannah Woodward) for coordinating and undertaking data collections and visits during the COVID-19 pandemic.

NJT is a Wellcome Trust Investigator (202802/Z/16/Z), is the PI of the Avon Longitudinal Study of Parents and Children (MRC & WT 217065/Z/19/Z), is supported by the University of Bristol NIHR Biomedical Research Centre (BRC-1215-2001), the MRC Integrative Epidemiology Unit (MC_UU_00011/1) and works within the CRUK Integrative Cancer Epidemiology Programme (C18281/A29019). MK is supported by the Medical Research Council (MR/W021315/1). AK is supported by Characterisation, determinants, mechanisms and consequences of the long-term effects of COVID-19: providing the evidence base for health care services (CONVALESCENCE) funded by NIHR (COV-LT-0009). SVK acknowledges funding from a NRS Senior Clinical Fellowship (SCAF/15/02), the Medical Research Council (MC_UU_00022/2) and the Scottish Government Chief Scientist Office (SPHSU17).

Antibody testing was funded by UK Health Security Agency. The National Core Studies program is funded by COVID-19 Longitudinal Health and Wellbeing – National Core Study (LHW-NCS) HMT/UKRI/MRC (MC_PC_20030 & MC_PC_20059). Related funding was also provided by the NIHR 606 (CONVALESCENCE grant COV-LT-0009).

TwinsUK is funded by the Wellcome Trust, Medical Research Council, Versus Arthritis, European Union Horizon 2020, Chronic Disease Research Foundation (CDRF), Zoe Ltd and the National Institute for Health Research (NIHR) Clinical Research Network (CRN) and Biomedical Research Centre based at Guy’s and St Thomas’ NHS Foundation Trust in partnership with King’s College London.

The UK Medical Research Council and Wellcome (Grant ref: 217065/Z/19/Z) and the University of Bristol provide core support for ALSPAC. We are extremely grateful to all the families who took part in this study, the midwives for their help in recruiting them, and the whole ALSPAC team, which includes interviewers, computer and laboratory technicians, clerical workers, research scientists, volunteers, managers, receptionists and nurses. This publication is the work of the authors and NJT and CJS serve as guarantors for the contents of this paper.

## Author contributions

Conceptualisation: NJC, MK, CJS, AW, NJT

Funding acquisition: CJS, ED, NJC, NC, NJT, MPG, GBP

Project administration: NJC, MK, AW, NJT, CJS, MPG, DH

Methodology: NJC, MK, AW, NJT, CJS, RJS, AK, DMW, OKLH, PHL, CHS, GDG, JZ, SVK, GBP

Formal analysis: NJC, MK, RCEB, EJT, XZ, GA

Investigation: NJC, MK

Visualisation: NJC, MK

Data curation: NJC, MK

In-house antibody testing: JS, SA, CG, NK, KJD, MHM

Writing – original draft: NJC, MK, CJS, AW, NJT, ED

Writing – review and editing: All authors

## Code availability

Copies of analysis codes are available upon request.

## Conflict of interest statement

All authors have completed the ICMJE uniform disclosure form at www.icmje.org/coi_disclosure.pdf. Financial support from organisations are detailed in the acknowledgements section. CJS is a consultant for Zoe Ltd. All remaining authors declare: no financial relationships with any commercial entities that might have an interest in the submitted work in the previous three years; no other relationships or activities that could appear to have influenced the submitted work.

## Additional information

**Supplementary information** is provided as additional files.

**Correspondence** and requests for materials should be addressed to Claire Steves and Nicholas Timpson.

## Notes

### Author Declarations

TwinsUK: All waves of TwinsUK have received ethical approval associated with TwinsUK Biobank (19/NW/0187), TwinsUK (EC04/015) or Healthy Ageing Twin Study (H.A.T.S) (07/H0802/84) studies from HRA/NHS Research Ethics Committees. The TwinsUK Resource Executive Committee (TREC) oversees management, data sharing and collaborations involving the TwinsUK registry (for further details see https://twinsuk.ac.uk/resources-for-researchers/access-our-data/), in consultation with the TwinsUK Volunteer Advisory Panel (VAP) where needed. ALSPAC: Ethical approval for the study was obtained from the ALSPAC Ethics and Law Committee and the Local Research Ethics Committees. Informed consent for the use of data collected via questionnaires and clinics was obtained from participants following the recommendations of the ALSPAC Ethics and Law Committee at the time. Consent for biological samples has been collected in accordance with the Human Tissue Act (2004). USoc: The University of Essex Ethics Committee has approved all data collection for the Understanding Society main study and COVID-19 web and telephone surveys (ETH1920-1271). The March 2021 web survey was reviewed and ethics approval granted by the NHS Health Research Authority, London - City & East Research Ethics Committee (reference 21/HRA/0644). No additional ethical approval was necessary for this secondary data analysis. 1958 NCDS, 1970 BCS70, Next Steps, MCS: The most recent sweeps of 1958 NCDS, 1970 BCS, Next Steps and MCS have all been granted ethical approval by the National Health Service (NHS) Research Ethics Committee and all participants have given informed consent. ELSA: Waves 1-9 of ELSA were approved by the London Multicentre Research Ethics Committee (approval number MREC/01/2/91), and the COVID-19 sub-study was approved by the University College London Research Ethics Committee (0017/003). All participants provided informed consent. 1946 NSHD: Ethical approval for the study was obtained from the NHS Research Ethics Committee (19/LO/1774). All participants provided informed consent. SABRE: Ethical approval for the study was obtained from the NHS Research Ethics Committee (19/LO/1774). All participants provided informed consent. EXCEED: The original EXCEED study was approved by the Leicester Central Research Ethics Committee (Ref: 13/EM/0226). Substantial amendments have been approved by the same Research Ethics Committee for the collection of new data relating to the COVID-19 pandemic, including the COVID-19 questionnaires and antibody testing.

