## Supplemental File 1 for "Antibody levels following vaccination against SARS-CoV-2: associations with post-vaccination infection and risk factors"

#### Figure and table locations

Note: All main text and supplementary tables are provided in a separate spreadsheet file. Tables available in the separate spreadsheet file only are noted below.

| Item | Page |
|---|---|
| Table S 1 | Separate spreadsheet file |
| Table S 2 | 10 |
| Table S 3 | 15 |
| Table S 4 | 17 |
| Table S 5 | 19 |
| Table S 6 | 28 |
| Table S 7 | 30 |
| Table S 8 | 31 |
| Table S 9 | 36 |
| Table S 10 | 52 |
| Table S 11 | 53 |
| Figure S 1 | 6 |
| Figure S 2 | 7 |
| Figure S 3 | 8 |
| Figure S 4 | 16 |
| Figure S 5 | 18 |
| Figure S 6 | 35 |
| Figure S 7 | 44 |
| Figure S 8 | 45 |
| Figure S 9 | 46 |
| Figure S 10 | 47 |
| Figure S 11 | 48 |
| Figure S 12 | 49 |
| Figure S 13 | 50 |
| Figure S 14 | 51 |
| Figure S 15 | 52 |

#### Additional ethics statements

TwinsUK: All waves of TwinsUK have received ethical approval associated with TwinsUK Biobank (19/NW/0187), TwinsUK (EC04/015) or Healthy Ageing Twin Study (H.A.T.S) (07/H0802/84) studies from HRA/NHS Research Ethics Committees. The TwinsUK Resource Executive Committee (TREC) oversees management, data sharing and collaborations involving the TwinsUK registry (for further details see https://twinsuk.ac.uk/resources-for-researchers/access-our-data/), in consultation with the TwinsUK Volunteer Advisory Panel (VAP) where needed.

ALSPAC: Ethical approval for the study was obtained from the ALSPAC Ethics and Law Committee and the Local Research Ethics Committees. Informed consent for the use of data collected via questionnaires and clinics was obtained from participants following the recommendations of the ALSPAC Ethics and Law Committee at the time. Consent for biological samples has been collected in accordance with the Human Tissue Act (2004).

USoc: The University of Essex Ethics Committee has approved all data collection for the Understanding Society main study and COVID-19 web and telephone surveys (ETH1920-1271). The March 2021 web survey was reviewed and ethics approval granted by the NHS Health Research Authority, London – City & East Research Ethics Committee (reference 21/HRA/0644). No additional ethical approval was necessary for this secondary data analysis.

1958 NCDS, 1970 BCS70, Next Steps, MCS: The most recent sweeps of 1958 NCDS, 1970 BCS, Next Steps and MCS have all been granted ethical approval by the National Health Service (NHS) Research Ethics Committee and all participants have given informed consent.

ELSA: Waves 1-9 of ELSA were approved by the London Multicentre Research Ethics Committee (approval number MREC/01/2/91), and the COVID-19 sub-study was approved by the University College London Research Ethics Committee (0017/003). All participants provided informed consent.

1946 NSHD: Ethical approval for the study was obtained from the NHS Research Ethics Committee (19/LO/1774). All participants provided informed consent.

SABRE: Ethical approval for the study was obtained from the NHS Research Ethics Committee (19/LO/1774). All participants provided informed consent.

EXCEED: The original EXCEED study was approved by the Leicester Central Research Ethics Committee (Ref: 13/EM/0226). Substantial amendments have been approved by the same Research Ethics Committee for the collection of new data relating to the COVID-19 pandemic, including the COVID-19 questionnaires and antibody testing.

#### Additional study funding statements

USoc: Understanding Society is an initiative funded by the Economic and Social Research Council and various Government Departments, with scientific leadership by the Institute for Social and Economic Research, University of Essex, and survey delivery by NatCen Social Research and Kantar Public. The Understanding Society COVID-19 study is funded by the Economic and Social Research Council (ES/K005146/1) and the Health Foundation (2076161). Serology testing was funded by the COVID-19 Longitudinal Health and Wellbeing – National Core Study (MC_PC_20030). The research data are distributed by the UK Data Service (SN 6614 and SN 8644).

1958 NCDS, 1970 BCS70, Next Steps, MCS: National Child Development Study 1958, British Cohort Study 1970, Next Steps, and The Millennium Cohort Study are supported by the Centre for Longitudinal Studies, Resource Centre 2015-20 grant (ES/M001660/1) and a host of other co-founders.

ELSA: The English Longitudinal Study of Ageing was developed by a team of researchers based at University College London, NatCen Social Research, the Institute for Fiscal Studies, the University of Manchester and the University of East Anglia. The data were collected by NatCen Social Research. The funding is currently provided by the National Institute on Aging in the US, and a consortium of UK government departments coordinated by the National Institute for Health Research. Funding has also been received by the Economic and Social Research Council. The English Longitudinal Study of Ageing Covid-19 Substudy was supported by the UK Economic and Social Research Grant (ESRC) ES/V003941/1.

1946 NSHD: The NSHD is funded by the MRC (MC_UU_00019/1). Serology testing was funded by the COVID-19 Longitudinal Health and Wellbeing – National Core Study (MC_PC_20030).

SABRE: The SABRE study is funded at baseline by the British Heart Foundation and the Wellcome Trust. Serology testing was funded by the COVID-19 Longitudinal Health and Wellbeing – National Core Study (MC_PC_20030).

EXCEED: The study is supported by the University of Leicester, the NIHR Leicester Respiratory Biomedical Research Centre, by Wellcome [202849, https://doi.org/10.35802/202849] and by Cohort Access fees from studies funded by the Medical Research Council (MRC), BBRSC, NIHR, the UK Space Agency, and GSK. It was previously supported by MRC grant G0902313.

#### Additional data statements

ALSPAC study data were collected and managed using REDCap electronic data capture tools hosted at the University of Bristol. REDCap (Research Electronic Data Capture) is a secure, web-based software platform designed to support data capture for research studies [1]. The study website contains details of all the data that is available through a fully searchable data dictionary and variable search tool on the study website [2].

#### Analysis sample flow charts

Notes: Unknown vaccination status included a small number of individuals with contradictory vaccination dates (e.g., first vaccination dated after second vaccination), in addition to those who did not complete vaccination status questions.

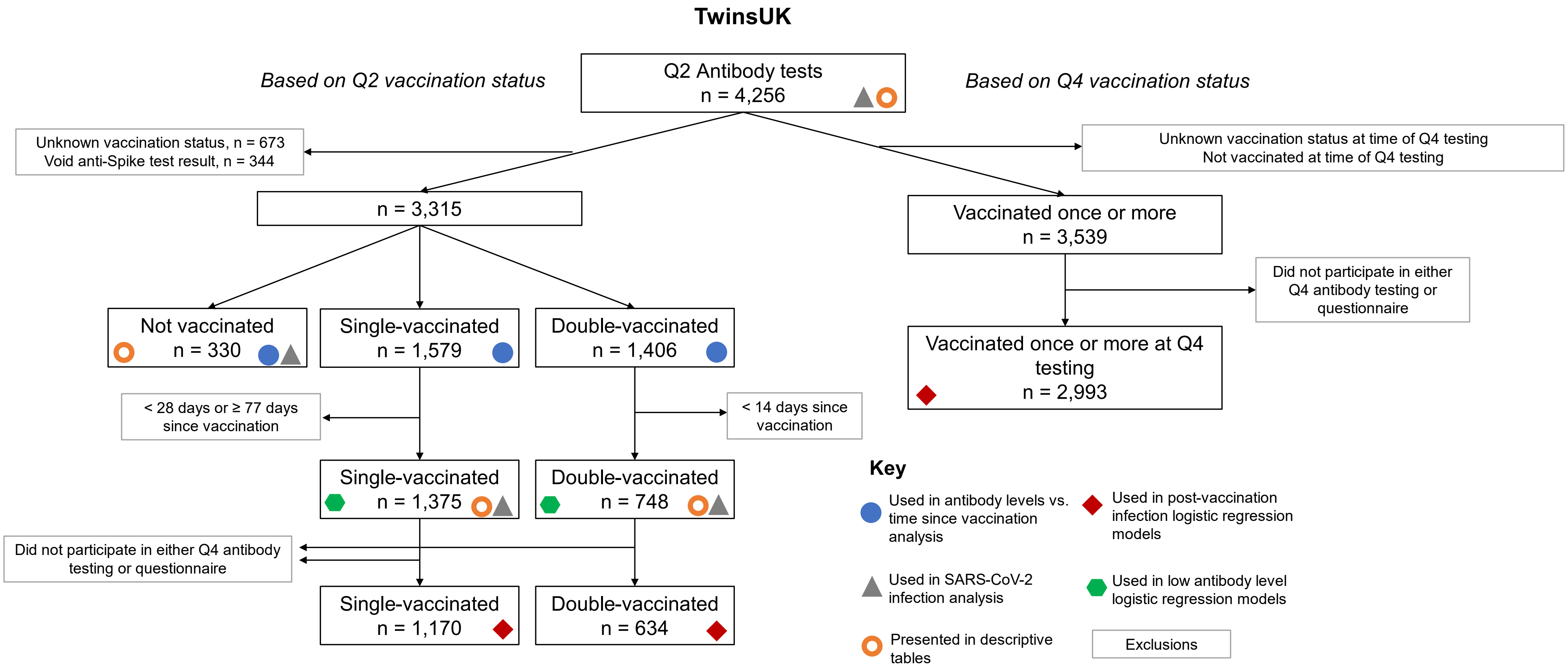

Figure S 1. Flow chart showing identification of analysis samples from Q2 antibody testing within TwinsUK. The use of groups of individuals in various analyses is highlighted with symbols.

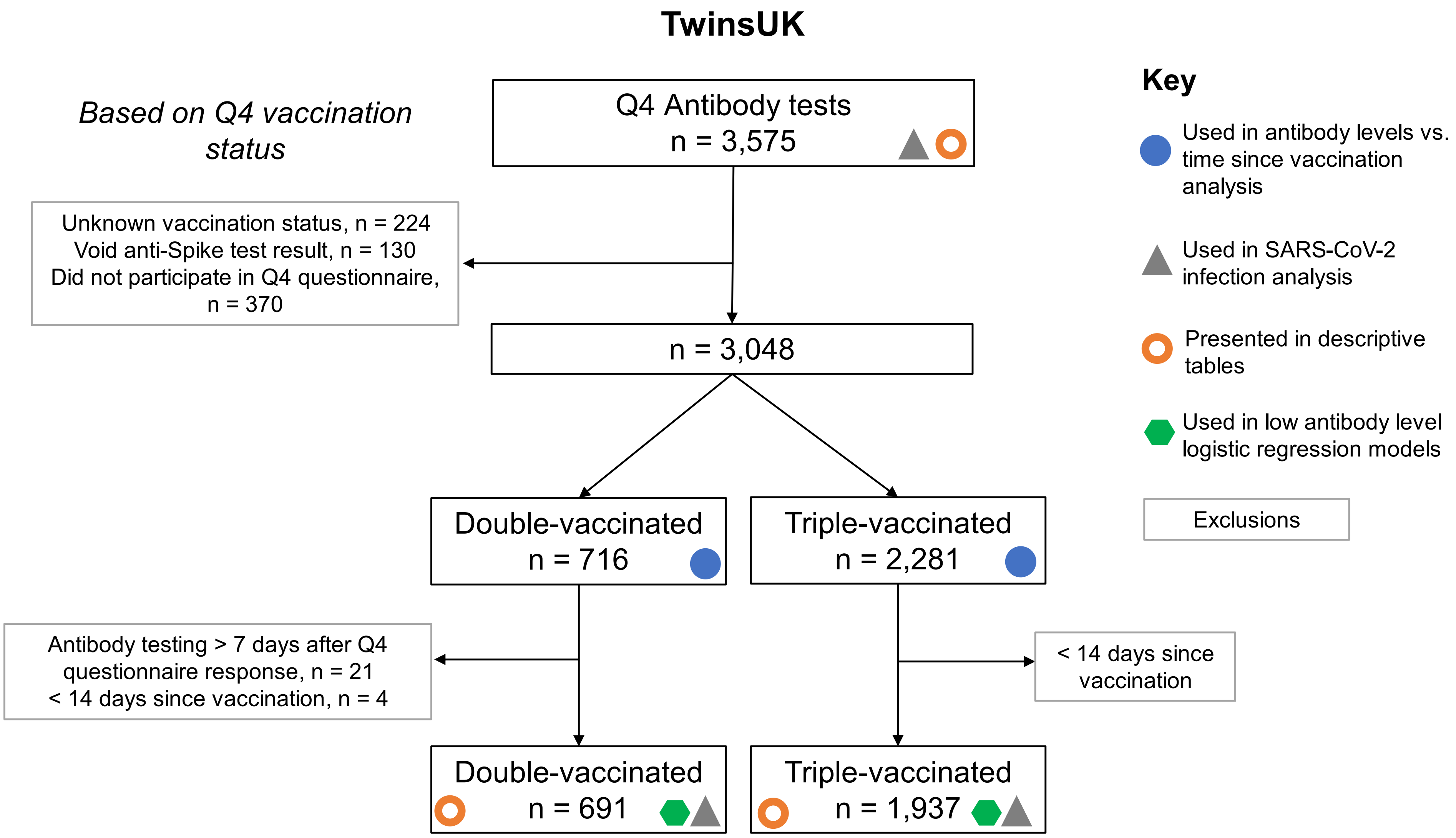

Figure S 2. Flow chart showing identification of analysis samples from Q4 antibody testing within TwinsUK. The use of groups of individuals in various analyses is highlighted with symbols.

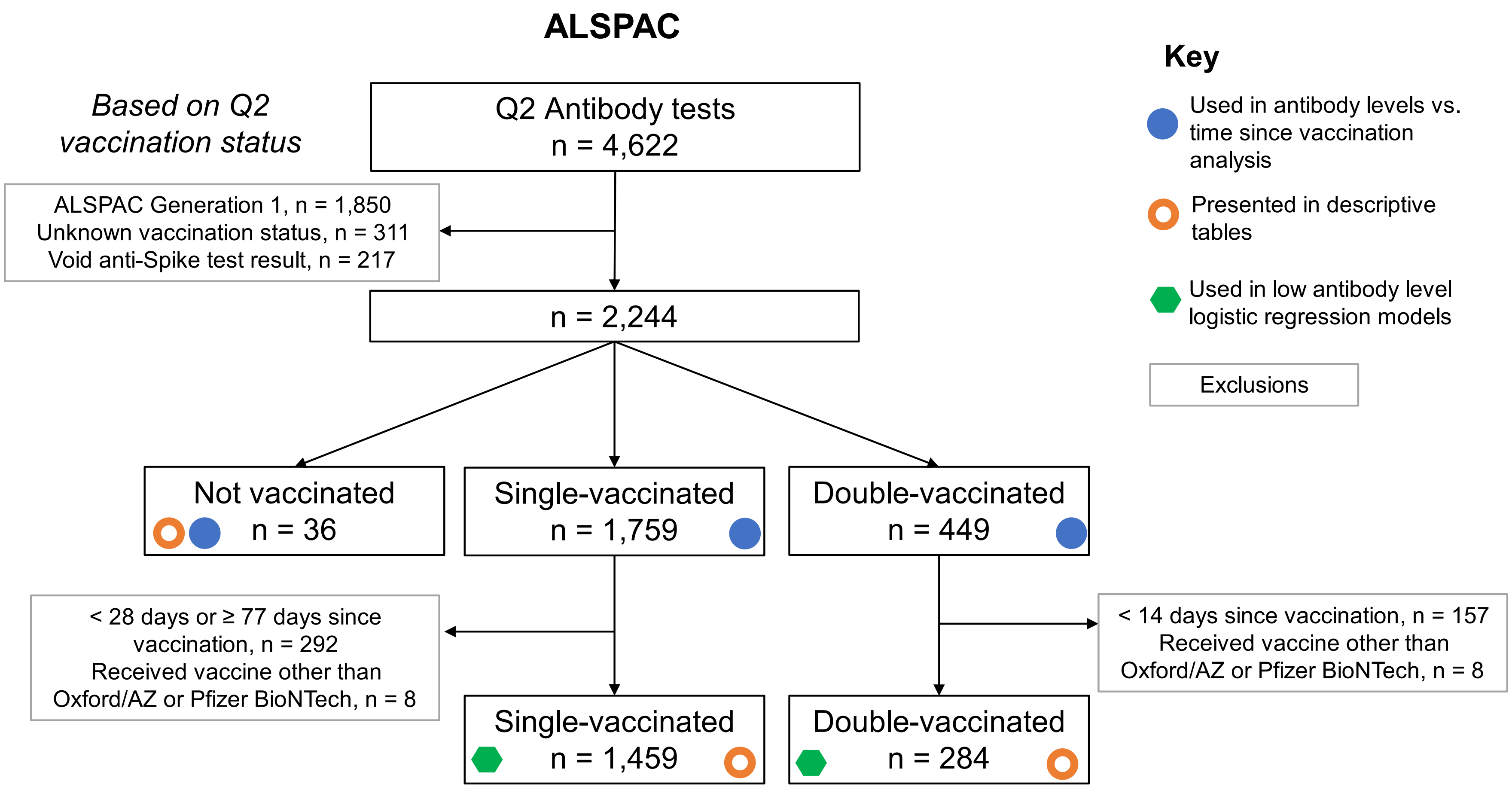

Figure S 3. Flow chart showing identification of analysis samples from Q2 antibody testing within ALSPAC. The use of groups of individuals in various analyses is highlighted with symbols.

#### Variable information

Detailed variable information is provided in supplementary spreadsheet file.

Table S 1. Information on origin of variables used in TwinsUK and ALSPAC analysis.

#### Extended sample characteristics

Extended sample characteristics are also provided in supplementary spreadsheet file for easier viewing.

Table S 2. Anti-Spike antibody level values and characteristics for individuals from TwinsUK sampled in Q2 and Q4 antibody collections. Individuals are stratified by vaccination status at time of sampling. Data shown for individuals sampled at least 4 (2) weeks after first (second or third) vaccination. The antibody level assay range is 0.4 to 250 BAU/mL for Q2 results and 0.4 to 25,000 BAU/mL for Q4 results, with a positive threshold of 0.8 BAU/mL. Categories with fewer than 5 individuals are suppressed.

| **Cohort** | **TwinsUK** | | | | | | | **ALSPAC** | | | |
|---|---|---|---|---|---|---|---|---|---|---|---|
| **Testing period** | **Q2** | **Q4** | **Q2** | | | **Q4** | | **Q2** | | | |
| **Vaccination status** | **All results** | **All results** | **Not vaccinated** | **Single-vaccinated** | **Double-vaccinated** | **Double-vaccinated** | **Triple-vaccinated** | **All results** | **Not vaccinated** | **Single-vaccinated** | **Double-vaccinated** |
| n | 4256 | 3575 | 330 | 1375 | 748 | 691 | 1937 | 1779 | 36 | 1459 | 284 |
| Anti-Spike antibody level value (BAU/mL): Median (IQR) | 80.78 (18.55, 250.0) | 10403.0 (3510.0, 20224.0) | 0.4 (0.4, 0.4) | 53.3 (22.72, 121.2) | 250.0 (250.0, 250.0) | 1317.0 (337.0, 5202.5) | 13694.0 (8153.0, 23543.0) | 58.93 (21.25, 247.5) | 10.53 (0.4, 48.69) | 43.42 (17.98, 106.65) | 250.0 (250.0, 250.0) |
| Anti-Spike antibody level value (BAU/mL): 5th & 10th percentile | 0.4, 0.4 | 134.6, 401.4 | 0.4, 0.4 | 4.58, 9.59 | 152.83, 250.0 | 73.7, 112.0 | 3446.0, 5036.0 | 3.19, 7.28 | 0.4, 0.4 | 3.2, 6.51 | 155.61, 250.0 |
| Anti-Spike antibody status: Positive, n (%) | 3372/3912 (86.2%) | 3423/3445 (99.4%) | 79/330 (23.9%) | 1357/1375 (98.7%) | 745/748 (99.6%) | 690/691 (99.9%) | 1936/1937 (99.9%) | 1745/1779 (98.1%) | 23/36 (63.9%) | 1440/1459 (98.7%) | 282/284 (99.3%) |
| Age (years): Median (IQR) | 63.0 (49.0, 72.0) | 63.0 (51.0, 72.0) | 38.0 (31.0, 44.0) | 63.0 (56.0, 69.0) | 70.0 (56.0, 77.0) | 49.0 (38.0, 59.0) | 69.0 (60.0, 74.0) | 60.0 (57.0, 62.0) | 57.5 (52.75, 62.25) | 60.0 (57.0, 63.0) | 59.0 (56.0, 61.0) |
| Sex: Male, n (%) | 518/4255 (12.2%) | 447/3574 (12.5%) | 48/330 (14.5%) | 178/1375 (12.9%) | 88/748 (11.8%) | 103/691 (14.9%) | 225/1937 (11.6%) | 451/1779 (25.4%) | 8/36 (22.2%) | 397/1459 (27.2%) | 46/284 (16.2%) |
| Ethnicity: Other than White, n (%) | 118/4219 (2.8%) | 96/3536 (2.7%) | 16/329 (4.9%) | 29/1368 (2.1%) | 19/739 (2.6%) | 26/686 (3.8%) | 39/1914 (2.0%) | 26/1779 (1.5%) | < 5 | 20/1459 (1.4%) | 5/284 (1.8%) |
| Local area deprivation, IMD decile: Median (IQR) | 7.0 (5.0, 9.0) | 7.0 (5.0, 9.0) | 7.0 (5.0, 9.0) | 8.0 (5.0, 9.0) | 7.0 (5.0, 9.0) | 7.0 (5.0, 9.0) | 8.0 (5.0, 9.0) | | | | |
| Local area deprivation, IMD: Most deprived 40% (decile 1-4), n (%) | 766/4242 (18.1%) | 640/3564 (18.0%) | 81/330 (24.5%) | 214/1371 (15.6%) | 124/748 (16.6%) | 151/690 (21.9%) | 298/1933 (15.4%) | 384/1213 (31.7%) | 9/24 (37.5%) | 305/971 (31.4%) | 70/218 (32.1%) |
| Highest educational attainment: NVQ level 3 or lower, n (%) | 1472/3493 (42.1%) | 1253/3025 (41.4%) | 49/224 (21.9%) | 497/1206 (41.2%) | 275/652 (42.2%) | 171/525 (32.6%) | 763/1732 (44.1%) | 1176/1694 (69.4%) | 30/36 (83.3%) | 936/1388 (67.4%) | 210/270 (77.8%) |
| Zygosity: Monozygotic, n (%) | 2722/4253 (64.0%) | 2280/3573 (63.8%) | 248/328 (75.6%) | 883/1375 (64.2%) | 459/748 (61.4%) | 490/689 (71.1%) | 1170/1937 (60.4%) | | | | |
| Weeks since first vaccination: Median (IQR) | 10.0 (6.0, 12.0) | 42.0 (38.0, 45.0) | -5.0 (-8.0, -3.0) | 8.0 (6.0, 9.0) | | | | | | 6.0 (5.0, 8.0) | |
| First vaccination received: AZD1222, n (%) | 2124/3591 (59.1%) | 1980/3378 (58.6%) | 70/275 (25.5%) | 1103/1374 (80.3%) | | | | | | 1235/1459 (84.6%) | |

| | | | | | | | | | | | |
|---|---|---|---|---|---|---|---|---|---|---|---|
| First vaccination received: BNT162b2, n (%) | 1410/3591 (39.3%) | 1336/3378 (39.6%) | 170/275 (61.8%) | 266/1374 (19.4%) | | | | | | 224/1459 (15.4%) | |
| First vaccination received: Other, n (%) | 57/3591 (1.6%) | 62/3378 (1.8%) | 35/275 (12.7%) | 5/1374 (0.4%) | | | | | | | |
| Weeks since second vaccination: Median (IQR) | -1.0 (-4.0, 2.0) | 32.0 (28.0, 34.0) | | | 3.0 (2.0, 5.0) | 25.0 (20.0, 28.0) | 33.0 (31.0, 35.0) | | | | 4.0 (2.0, 6.0) |
| Second vaccination received: AZD1222, n (%) | 1858/3266 (56.9%) | 1888/3275 (57.6%) | | | 212/748 (28.3%) | 411/691 (59.5%) | 1065/1934 (55.1%) | | | | 50/284 (17.6%) |
| Second vaccination received: BNT162b2, n (%) | 1357/3266 (41.5%) | 1330/3275 (40.6%) | | | 532/748 (71.1%) | 241/691 (34.9%) | 858/1934 (44.4%) | | | | 234/284 (82.4%) |
| Second vaccination received: Other, n (%) | 51/3266 (1.6%) | 57/3275 (1.7%) | | | < 5 | 39/691 (5.6%) | 11/1934 (0.6%) | | | | |
| Weeks since third vaccination: Median (IQR) | -28.0 (-30.0, -26.0) | 5.0 (3.0, 7.0) | | | | | 5.0 (4.0, 8.0) | | | | |
| Third vaccination received: mRNA-1273, n (%) | 293/2149 (13.6%) | 337/2400 (14.0%) | | | | | 203/1903 (10.7%) | | | | |
| Third vaccination received: BNT162b2, n (%) | 1828/2149 (85.1%) | 2026/2400 (84.4%) | | | | | 1677/1903 (88.1%) | | | | |
| Third vaccination received: Other, n (%) | 28/2149 (1.3%) | 37/2400 (1.5%) | | | | | 23/1903 (1.2%) | | | | |
| SARS-CoV-2 infection status (serology-based) at time of antibody testing: Evidence of natural infection, n (%) | 891/4190 (21.3%) | 977/3561 (27.4%) | 98/330 (29.7%) | 304/1375 (22.1%) | 157/748 (21.0%) | 245/691 (35.5%) | 464/1937 (24.0%) | 187/1757 (10.6%) | 23/36 (63.9%) | 133/1438 (9.2%) | 31/283 (11.0%) |
| SARS-CoV-2 infection status (self-reported), Q2: Unsure, n (%) | 93/4092 (2.3%) | 60/3428 (1.8%) | 11/320 (3.4%) | 43/1365 (3.2%) | 9/739 (1.2%) | 12/662 (1.8%) | 30/1882 (1.6%) | | | | |
| SARS-CoV-2 infection status (self-reported), Q2: Suspected case, n (%) | 477/4092 (11.7%) | 399/3428 (11.6%) | 35/320 (10.9%) | 183/1365 (13.4%) | 67/739 (9.1%) | 81/662 (12.2%) | 197/1882 (10.5%) | 302/1675 (18.0%) | 5/33 (15.2%) | 240/1374 (17.5%) | 57/268 (21.3%) |
| SARS-CoV-2 infection status (self-reported), Q2: Confirmed case, n (%) | 597/4092 (14.6%) | 492/3428 (14.4%) | 57/320 (17.8%) | 218/1365 (16.0%) | 112/739 (15.2%) | 107/662 (16.2%) | 256/1882 (13.6%) | 40/1675 (2.4%) | < 5 | 29/1374 (2.1%) | 11/268 (4.1%) |
| SARS-CoV-2 infection status (self-reported), Q4: Unsure, n (%) | 147/4134 (3.6%) | 128/3543 (3.6%) | 19/330 (5.8%) | 67/1375 (4.9%) | 18/748 (2.4%) | 30/691 (4.3%) | 65/1936 (3.4%) | | | | |
| SARS-CoV-2 infection status (self-reported), Q4: Suspected case, n (%) | 478/4134 (11.6%) | 404/3543 (11.4%) | 34/330 (10.3%) | 183/1375 (13.3%) | 70/748 (9.4%) | 78/691 (11.3%) | 204/1936 (10.5%) | | | | |
| SARS-CoV-2 infection status (self-reported), Q4: Confirmed case, n (%) | 817/4134 (19.8%) | 751/3543 (21.2%) | 92/330 (27.9%) | 306/1375 (22.3%) | 145/748 (19.4%) | 202/691 (29.2%) | 357/1936 (18.4%) | | | | |
| Anti-Nucleocapsid antibody status, Q2: Positive, n (%) | 460/3893 (11.8%) | 333/2887 (11.5%) | 60/329 (18.2%) | 156/1368 (11.4%) | 87/743 (11.7%) | 85/565 (15.0%) | 160/1624 (9.9%) | 167/1757 (9.5%) | < 5 | 133/1438 (9.2%) | 31/283 (11.0%) |
| Anti-Nucleocapsid antibody status, Q4: Positive, n (%) | 524/2998 (17.5%) | 618/3447 (17.9%) | 80/290 (27.6%) | 197/1130 (17.4%) | 95/602 (15.8%) | 179/691 (25.9%) | 263/1937 (13.6%) | | | | |
| Frailty Index: Frail, n (%) | 390/3316 (11.8%) | 312/2854 (10.9%) | 9/195 (4.6%) | 111/1138 (9.8%) | 90/622 (14.5%) | 45/478 (9.4%) | 192/1665 (11.5%) | | | | |
| Frailty Index: Very frail, n (%) | 112/3316 (3.4%) | 91/2854 (3.2%) | < 5 | 35/1138 (3.1%) | 23/622 (3.7%) | 7/478 (1.5%) | 56/1665 (3.4%) | | | | |

| | | | | | | | | | | | |
|---|---|---|---|---|---|---|---|---|---|---|---|
| Frailty (PRISMA-7): Frail, n (%) | | | | | | | | 50/1779 (2.8%) | < 5 | 40/1459 (2.7%) | 10/284 (3.5%) |
| Advised on "Shielded Patient List": Yes, n (%) | 341/4109 (8.3%) | 279/3530 (7.9%) | 8/329 (2.4%) | 82/1374 (6.0%) | 86/748 (11.5%) | 23/691 (3.3%) | 190/1936 (9.8%) | 67/1754 (3.8%) | < 5 | 45/1443 (3.1%) | 22/276 (8.0%) |
| Prescribed immunosuppressant medication: Yes, n (%) | 345/2824 (12.2%) | 291/2444 (11.9%) | 9/169 (5.3%) | 117/963 (12.1%) | 64/540 (11.9%) | 37/394 (9.4%) | 192/1462 (13.1%) | | | | |
| Immunocompromised: Yes, n (%) | | | | | | | | 54/1332 (4.1%) | < 5 | 39/1092 (3.6%) | 15/213 (7.0%) |
| Self-rated health: Poor, Fair, n (%) | 357/4082 (8.7%) | 290/3407 (8.5%) | 15/316 (4.7%) | 134/1364 (9.8%) | 60/737 (8.1%) | 42/656 (6.4%) | 168/1871 (9.0%) | 167/1778 (9.4%) | < 5 | 137/1459 (9.4%) | 28/283 (9.9%) |
| BMI: Median (IQR) | 24.75 (22.15, 27.99) | 24.76 (22.2, 27.98) | 22.72 (20.94, 25.42) | 24.86 (22.27, 28.28) | 24.99 (22.23, 28.07) | 23.91 (21.62, 27.58) | 24.87 (22.3, 27.87) | 25.7 (23.23, 28.7) | 25.63 (23.13, 28.04) | 25.71 (23.25, 28.77) | 25.65 (23.02, 28.6) |
| Number of selected comorbidities: 1+, n (%) | 1345/3047 (44.1%) | 1134/2617 (43.3%) | 46/190 (24.2%) | 452/1056 (42.8%) | 288/555 (51.9%) | 144/452 (31.9%) | 717/1506 (47.6%) | 387/1289 (30.0%) | 6/26 (23.1%) | 314/1058 (29.7%) | 67/205 (32.7%) |
| Comorbidity: Anxiety or Stress Disorder: Yes, n (%) | 474/3071 (15.4%) | 389/2635 (14.8%) | 29/177 (16.4%) | 165/1059 (15.6%) | 81/576 (14.1%) | 70/441 (15.9%) | 213/1548 (13.8%) | 131/1321 (9.9%) | < 5 | 114/1083 (10.5%) | 16/211 (7.6%) |
| Comorbidity: Depression: Yes, n (%) | 387/3039 (12.7%) | 328/2618 (12.5%) | 22/174 (12.6%) | 134/1051 (12.7%) | 70/574 (12.2%) | 54/437 (12.4%) | 180/1533 (11.7%) | 65/1330 (4.9%) | < 5 | 55/1092 (5.0%) | 9/211 (4.3%) |
| Anxiety (HADS) score up to Q2: Median (IQR) | 6.0 (3.0, 9.0) | 6.0 (3.0, 9.0) | 7.0 (4.0, 11.0) | 6.0 (3.0, 10.0) | 6.0 (3.0, 9.0) | 7.0 (4.0, 10.0) | 6.0 (3.0, 9.0) | | | | |
| Anxiety (HADS) score up to Q2: 8-10, mild, n (%) | 874/4074 (21.5%) | 730/3420 (21.3%) | 75/319 (23.5%) | 298/1361 (21.9%) | 153/736 (20.8%) | 143/661 (21.6%) | 392/1878 (20.9%) | | | | |
| Anxiety (HADS) score up to Q2: 11+, moderate, severe, n (%) | 752/4074 (18.5%) | 601/3420 (17.6%) | 82/319 (25.7%) | 262/1361 (19.3%) | 120/736 (16.3%) | 137/661 (20.7%) | 276/1878 (14.7%) | | | | |
| Depression (HADS) score up to Q2: Median (IQR) | 5.0 (2.0, 8.0) | 5.0 (2.0, 7.0) | 5.0 (3.0, 8.0) | 5.0 (2.0, 8.0) | 4.0 (2.0, 7.0) | 5.0 (3.0, 8.0) | 4.0 (2.0, 7.0) | | | | |
| Depression (HADS) score up to Q2: 8-10, mild, n (%) | 627/4076 (15.4%) | 511/3419 (14.9%) | 54/319 (16.9%) | 227/1362 (16.7%) | 112/736 (15.2%) | 95/660 (14.4%) | 275/1878 (14.6%) | | | | |
| Depression (HADS) score up to Q2: 11+, moderate, severe, n (%) | 398/4076 (9.8%) | 331/3419 (9.7%) | 36/319 (11.3%) | 151/1362 (11.1%) | 60/736 (8.2%) | 85/660 (12.9%) | 146/1878 (7.8%) | | | | |
| Anxiety (HADS) score up to Q4: Median (IQR) | 7.0 (4.0, 10.0) | 7.0 (4.0, 10.0) | 8.0 (5.0, 11.0) | 7.0 (4.0, 10.0) | 6.0 (3.0, 9.0) | 7.0 (5.0, 10.5) | 6.0 (3.0, 9.0) | | | | |
| Anxiety (HADS) score up to Q4: 8-10, mild, n (%) | 900/4121 (21.8%) | 776/3541 (21.9%) | 75/329 (22.8%) | 309/1372 (22.5%) | 160/746 (21.4%) | 144/691 (20.8%) | 431/1937 (22.3%) | | | | |
| Anxiety (HADS) score up to Q4: 11+, moderate, severe, n (%) | 846/4121 (20.5%) | 715/3541 (20.2%) | 98/329 (29.8%) | 297/1372 (21.6%) | 138/746 (18.5%) | 173/691 (25.0%) | 327/1937 (16.9%) | | | | |
| Depression (HADS) score up to Q4: Median (IQR) | 5.0 (3.0, 8.0) | 5.0 (3.0, 8.0) | 6.0 (3.0, 8.0) | 5.0 (3.0, 8.0) | 5.0 (3.0, 8.0) | 6.0 (3.0, 8.0) | 5.0 (3.0, 8.0) | | | | |
| Depression (HADS) score up to Q4: 8-10, mild, n (%) | 693/4124 (16.8%) | 598/3541 (16.9%) | 60/329 (18.2%) | 244/1373 (17.8%) | 129/746 (17.3%) | 106/691 (15.3%) | 331/1937 (17.1%) | | | | |
| Depression (HADS) score up to Q4: 11+, moderate, severe, n (%) | 455/4124 (11.0%) | 395/3541 (11.2%) | 46/329 (14.0%) | 175/1373 (12.7%) | 67/746 (9.0%) | 105/691 (15.2%) | 174/1937 (9.0%) | | | | |

#### Shielded patient list criteria

Summary of UK government "Shielded Patient List" criteria used to identify individuals at high or moderate risk of developing complications from coronavirus (COVID-19). Adapted from https://digital.nhs.uk/coronavirus/shielded-patient-list/risk-criteria.

High risk criteria:

- Solid organ transplant recipients
- People with severe respiratory conditions including all cystic fibrosis, severe asthma and severe chronic obstructive pulmonary (COPD)
- People with rare diseases and inborn errors of metabolism that significantly increase the risk of infections (such as Severe combined immunodeficiency (SCID), homozygous sickle cell)
- People on immunosuppression therapies sufficient to significantly increase risk of infection
- People who have problems with their spleen, for example have had a splenectomy
- Adults with Down's syndrome
- Adults on dialysis with kidney impairment (Stage 5 Chronic Kidney Disease)
- Women who are pregnant with significant heart disease, congenital or acquired
- People with cancer who are undergoing active chemotherapy
- people with lung cancer who are undergoing radical radiotherapy
- people with cancers of the blood or bone marrow such as leukaemia, lymphoma or myeloma who are at any stage of treatment
- people having immunotherapy or other continuing antibody treatments for cancer
- people having other targeted cancer treatments which can affect the immune system, such as protein kinase inhibitors or PARP inhibitors
- people who have had bone marrow or stem cell transplants in the last 6 months, or who are still taking immunosuppression drugs

Moderate risk criteria

Individuals who met the criteria that made them eligible for the annual flu vaccination (except those aged 65 to 69 years old inclusive who have no other qualifying conditions), or did not meet the CMO criteria for the high risk group for COVID-19, including those identified by the COVID-19 Population Risk Assessment. This included patients aged 70 or older (regardless of medical conditions), or patients under 70 years old who:

- have chronic (long-term) respiratory disease, such as asthma, chronic obstructive pulmonary disease (COPD), emphysema or bronchitis
- have chronic heart disease, such as heart failure
- have chronic kidney disease (Stage 1 to 4)
- have chronic liver disease, such as hepatitis
- have a chronic neurological condition, such as Parkinson's disease, motor neurone disease, multiple sclerosis (MS), a learning disability or cerebral palsy
- have diabetes
- have a weakened immune system caused by a medical condition or medications such as steroid tablets or chemotherapy
- are seriously overweight (a BMI of 40 or above)
- are pregnant

**Notes:**

Some speciality organisations (Association of British Neurologists, British Society of Gastroenterology (liver and IBD), The renal association, British Society for Rheumatology, British Association of Dermatologists, British Thoracic Society) had developed decision-support tools to help identify patients. Please note that this is guidance, and ultimately the decision to add a person to the high risk category would be on a case-by-case basis.

Lists are non-exhaustive, with the decision to add a person to the shielding patient list able to be done on a case-by-case basis.

For adults, the moderate risk criteria were comprised of anyone usually instructed to get a flu jab as an adult each year on medical grounds (except those aged 65 to 69 years old inclusive who have no other qualifying conditions).

#### TwinsUK SARS-CoV-2 infection descriptive statistics

Table S 3. SARS-CoV-2 infection prevalence rates, split by selected socio-demographic variables, for TwinsUK Q4 antibody testing participants. P-values are generated from chi-square test of independence on cross tabulation of counts for the socio-demographic variable of interest and all categories (including those not presented) of the SARS-CoV-2 infection variable.

| Variable | SARS-CoV-2 infection status (self-reported): Suspected case | SARS-CoV-2 infection status (self-reported): Confirmed case | SARS-CoV-2 infection status (serology-based): Evidence of natural infection | Anti-Nucleocapsid antibody status: Positive |
|---|---|---|---|---|
| Overall | 404/3543 (11.4%) | 751/3543 (21.2%) | 977/3560 (27.4%) | 618/3447 (17.9%) |
| Sex: Female | 362/3101 (11.7%) [p = 0.41] | 650/3101 (21.0%) [p = 0.41] | 837/3113 (26.9%) [p = 0.03] | 530/3016 (17.6%) [p = 0.18] |
| Sex: Male | 41/441 (9.3%) | 101/441 (22.9%) | 139/446 (31.2%) | 87/430 (20.2%) |
| Age: 18-29 | 14/131 (10.7%) [p < 0.0001] | 44/131 (33.6%) [p < 0.0001] | 60/133 (45.1%) [p < 0.0001] | 48/130 (36.9%) [p < 0.0001] |
| Age: 30-39 | 43/292 (14.7%) | 93/292 (31.8%) | 116/298 (38.9%) | 80/290 (27.6%) |
| Age: 40-49 | 57/376 (15.2%) | 107/376 (28.5%) | 141/380 (37.1%) | 107/374 (28.6%) |
| Age: 50-59 | 94/671 (14.0%) | 183/671 (27.3%) | 224/675 (33.2%) | 168/658 (25.5%) |
| Age: 60-69 | 109/904 (12.1%) | 175/904 (19.4%) | 228/905 (25.2%) | 140/884 (15.8%) |
| Age: 70-79 | 74/962 (7.7%) | 130/962 (13.5%) | 174/964 (18.0%) | 67/928 (7.2%) |
| Age: 80+ | 13/207 (6.3%) | 19/207 (9.2%) | 34/205 (16.6%) | 8/183 (4.4%) |
| Ethnicity: Other than white | 12/96 (12.5%) [p = 0.74] | 22/96 (22.9%) [p = 0.74] | 44/96 (45.8%) [p = 0.0001] | 21/96 (21.9%) [p = 0.30] |
| Ethnicity: White | 385/3411 (11.3%) | 724/3411 (21.2%) | 925/3426 (27.0%) | 589/3314 (17.8%) |
| IMD: Quintile 1 (most deprived 20%) | 29/206 (14.1%) [p = 0.002] | 60/206 (29.1%) [p = 0.002] | 71/210 (33.8%) [p = 0.21] | 56/200 (28.0%) [p = 0.004] |
| IMD: Quintile 2 | 54/423 (12.8%) | 77/423 (18.2%) | 120/426 (28.2%) | 77/409 (18.8%) |
| IMD: Quintile 3 | 95/725 (13.1%) | 147/725 (20.3%) | 189/724 (26.1%) | 116/697 (16.6%) |
| IMD: Quintile 4 | 118/918 (12.9%) | 185/918 (20.2%) | 244/927 (26.3%) | 153/900 (17.0%) |
| IMD: Quintile 5 (least deprived 20%) | 107/1261 (8.5%) | 281/1261 (22.3%) | 351/1262 (27.8%) | 215/1230 (17.5%) |
| RUC: Rural | 106/911 (11.6%) [p = 0.07] | 168/911 (18.4%) [p = 0.07] | 203/912 (22.3%) [p < 0.0001] | 125/884 (14.1%) [p = 0.0005] |
| RUC: Urban | 296/2603 (11.4%) | 581/2603 (22.3%) | 770/2617 (29.4%) | 490/2532 (19.4%) |

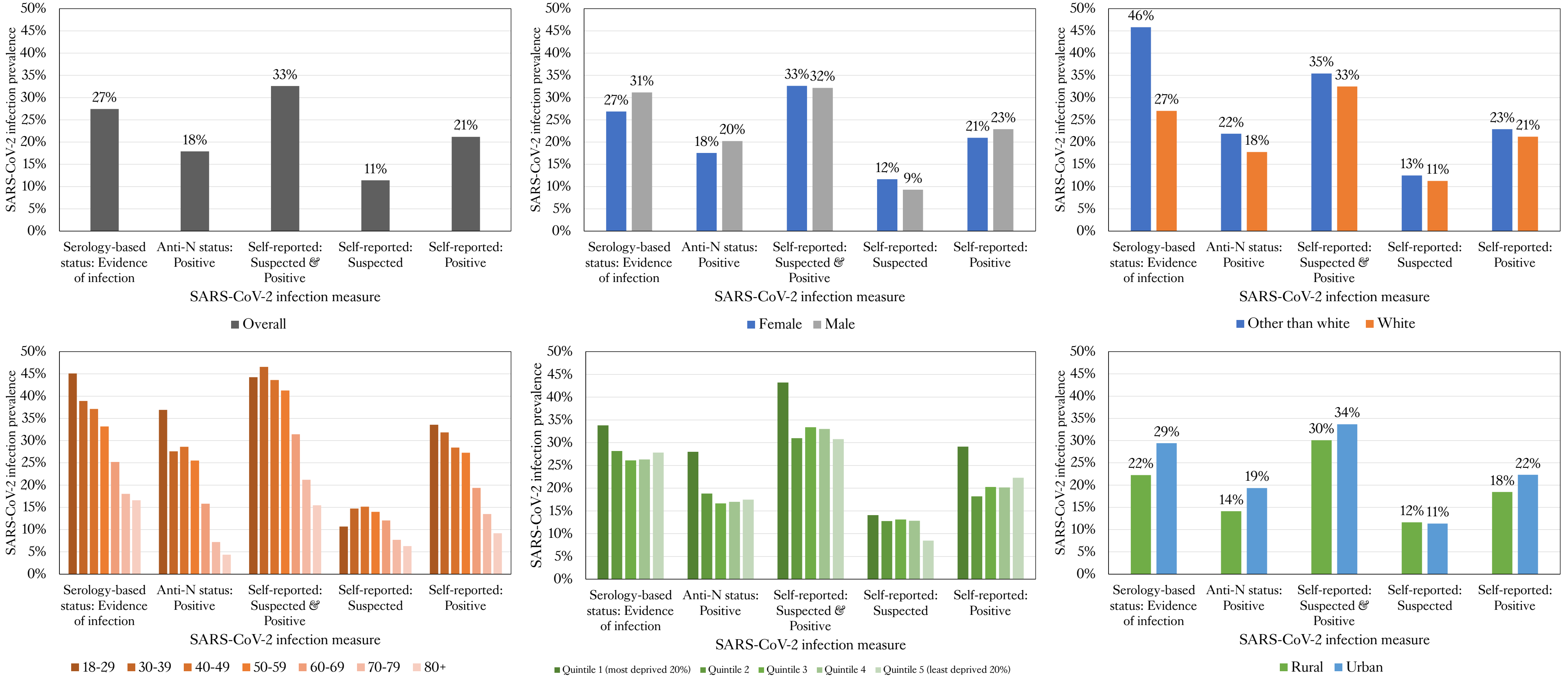

Figure S 4. Prevalence of SARS-CoV-2 infection for serology-based and self-reported measures of infection, for all individuals sampled in TwinsUK Q4 antibody testing, overall and split by socio-demographic variables: age, sex, ethnicity, local area deprivation (IMD), and rural-urban classification. Anti-N: Anti-Nucleocapsid.

#### All-cohort antibody testing summary

Table S 4. Summary counts and seropositivity rates from Q2 antibody testing conducted across 11 UK-based longitudinal cohorts. TwinsUK Q4 testing results also shown for completeness. Cohorts listed in ascending order of average age.

| | | | | Individual antibody results, n (%*) | | | Combined antibody results, n (%*) | | |
|---|---|---|---|---|---|---|---|---|---|
| **Cohort** | **Area** | **Characteristics** | **Mean age (years)** | **n** | **Anti-Nucleocapsid positive** | **Anti-Spike positive** | **Anti-Spike & anti-Nucleocapsid negative** | **Anti-Spike positive only** | **Anti-Spike & anti-Nucleocapsid positive** |
| **Millennium Cohort Study (MCS): Parents** | **Nationwide** | **Birth cohort, family structure (parent & child)** | 53^ | 2266 | 283 (13.6%) | 1888 (90.2%) | 201 (9.7%) | 1587 (76.6%) | 283 (13.7%) |
| **Millennium Cohort Study (MCS): Children** | | | 20 | 1141 | 229 (22.5%) | 427 (41.8%) | 590 (58.2%) | 197 (19.4%) | 226 (22.3%) |
| **Next Steps** | **Nationwide** | **Younger population** | 31 | 1267 | 166 (14.5%) | 514 (44.5%) | 632 (55.7%) | 339 (29.9%) | 162 (14.3%) |
| **ALSPAC (Avon Longitudinal Study of Parents and Children): G0 (parents)** | **Bristol** | **Birth cohort, family structure (parent & child), local population** | 60 | 2772 | 248 (9.8%) | 2463 (96.4%) | 91 (3.6%) | 2183 (86.6%) | 247 (9.8%) |
| **ALSPAC (Avon Longitudinal Study of Parents and Children): G1 (children)** | | | 29 | 1850 | 245 (14.6%) | 709 (42.1%) | 970 (58.1%) | 455 (27.2%) | 242 (14.5%) |
| **Understanding Society (USoc)** | **Nationwide** | **Nationally representative, family structure (households)** | 49 | 6667 | 599 (10%) | 4793 (79.3%) | 1239 (20.7%) | 4145 (69.3%) | 591 (9.9%) |
| **British Cohort Study 1970 (1970 BCS)** | **Nationwide** | **Birth cohort** | 51 | 2547 | 261 (11.2%) | 2230 (95%) | 117 (5%) | 1955 (83.9%) | 257 (11%) |
| **TwinsUK [Q2]** | **Nationwide** | **Family structure (twin pairs)** | 59 | 4256 | 460 (11.8%) | 3372 (86.2%) | 535 (13.8%) | 2889 (74.4%) | 457 (11.8%) |
| **TwinsUK [Q4]** | | | 60 | 3575 | 618 (17.9%) | 3423 (99.4%) | 22 (0.6%) | 2805 (81.4%) | 618 (17.9%) |
| **Extended Cohort for E-health, Environment and DNA (EXCEED)** | **Leicester** | **Local population** | 62 | 2411 | 186 (8.5%) | 2100 (94.6%) | 119 (5.4%) | 1883 (86.1%) | 186 (8.5%) |
| **National Child Development Study (1958 NCDS)** | **Nationwide** | **Birth cohort** | 63 | 3223 | 244 (8.4%) | 2850 (97.7%) | 68 (2.4%) | 2573 (89.2%) | 244 (8.5%) |
| **English longitudinal Study of Ageing (ELSA)** | **Nationwide** | **Older population** | 70 | 3513 | 258 (8.8%) | 2847 (96.8%) | 89 (3.1%) | 2553 (88.1%) | 252 (8.7%) |
| **Southall and Brent Revisited (SABRE)** | **London** | **Ethnicity diversity** | 74 | 176 | 13 (9.6%) | 139 (98.6%) | < 5 | 120 (89.6%) | 12 (9%) |
| **National Survey of Health and Development (1946 NSHD)** | **Nationwide** | **Birth cohort** | 75 | 896 | 36 (4.7%) | 769 (96.7%) | 26 (3.4%) | 710 (92%) | 36 (4.7%) |
| **Total [Q2]** | | | | 40816 | 4306 (11.7%) | 31896 (85.8%) | 5236 (14.2%) | 27283 (74.1%) | 4270 (11.6%) |

Notes: * Void results excluded in calculation of quoted percentages. ^ MCS cohort parents mean age is a best estimate based on a mean age of 50 of all parents participating in MCS Age 17 Sweep undertaken in 2018, https://cls.ucl.ac.uk/cls-studies/millennium-cohort-study/mcs-age-17-sweep/.

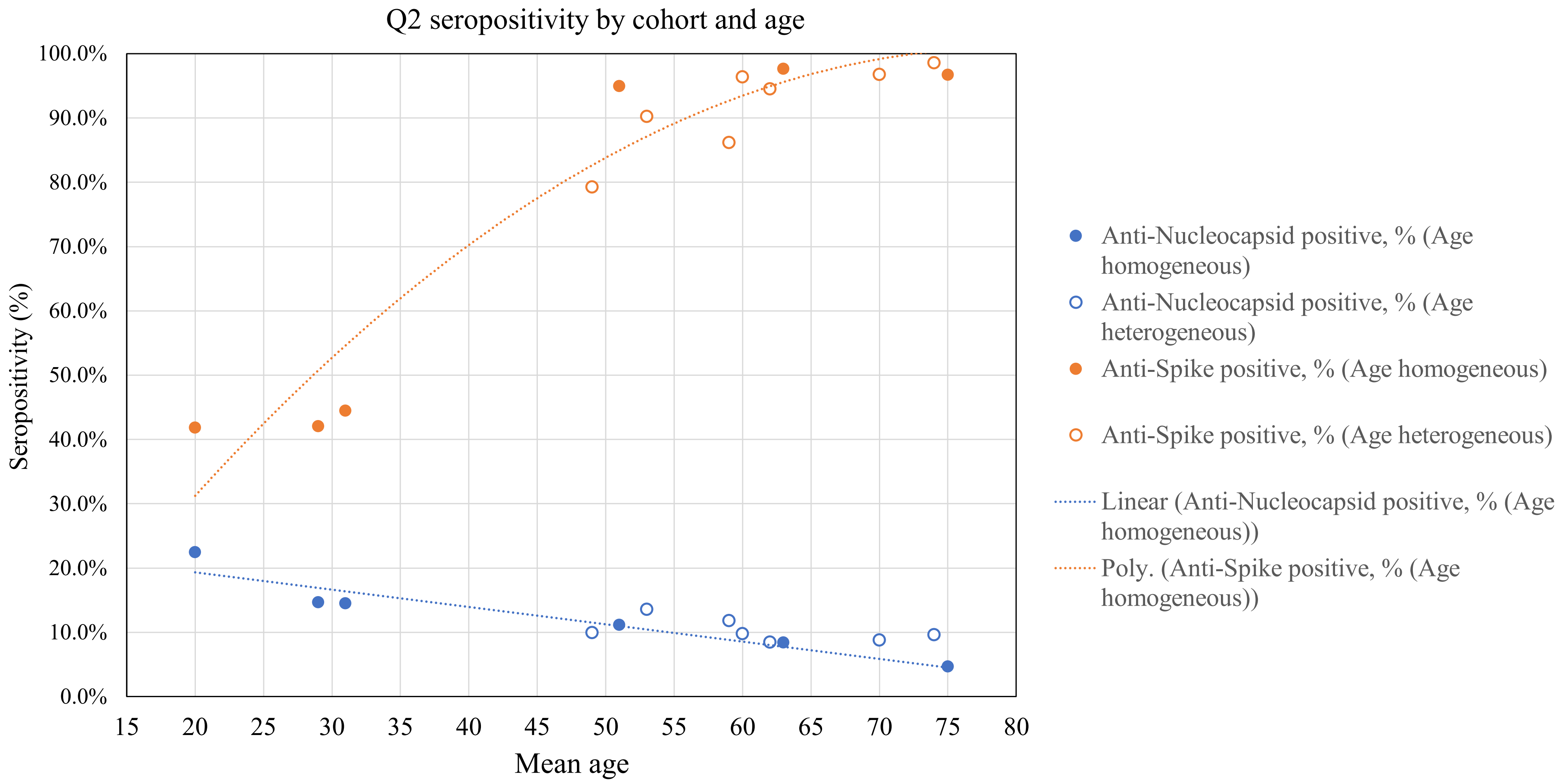

Figure S 5. Seropositivity rates versus mean age from Q2 antibody testing conducted across 11 UK-based longitudinal cohorts. Age homogeneous birth cohorts and age heterogeneous cohorts are plotted as separate series. Linear and polynomial trend lines are added to age-homogenous cohort data to guide the eye.

#### Antibody level descriptive statistics

Descriptive statistics of anti-Spike antibody levels are also provided in supplementary spreadsheet file for easier viewing.

Table S 5. Anti-Spike antibody levels and weeks since most recent vaccination within TwinsUK and ALSPAC individuals, stratified by vaccination status at Q2 and Q4 antibody testing, split by various variables. The antibody level assay range is 0.4 to 250 BAU/mL for Q2 results and 0.4 to 25,000 BAU/mL for Q4 results, with a positive threshold of 0.8 BAU/mL.

| **Cohort** | **TwinsUK** | | | | **ALSPAC** | |
|---|---|---|---|---|---|---|
| **Testing period** | **Q2** | | **Q4** | | **Q2** | |
| **Vaccination status** | **Single-vaccinated** | **Double-vaccinated** | **Double-vaccinated** | **Triple-vaccinated** | **Single-vaccinated** | **Double-vaccinated** |
| Age group: 18-49, Anti-Spike level | n = 145, Median: 52.58, IQR: (19.25, 180.6), 5%: 4.57, 10%: 8.22 | n = 123, Median: 250.0, IQR: (250.0, 250.0), 5%: 250.0, 10%: 250.0 | n = 352, Median: 2130.5, IQR: (485.0, 5662.75), 5%: 91.76, 10%: 201.5 | n = 215, Median: 12844.0, IQR: (8257.0, 22363.5), 5%: 3310.8, 10%: 4771.0 | n = 8, Median: 167.77, IQR: (57.98, 250.0), 5%: 21.52, 10%: 25.04 | n = 6, Median: 250.0, IQR: (250.0, 250.0), 5%: 250.0, 10%: 250.0 |
| Age group: 18-49, Weeks since vaccination | Median: 6.0, IQR: (5.0, 8.0) | Median: 5.0, IQR: (3.0, 6.5) | Median: 21.0, IQR: (18.0, 24.0) | Median: 6.0, IQR: (4.0, 9.0) | Median: 6.0, IQR: (5.0, 7.0) | Median: 6.0, IQR: (2.75, 12.25) |
| Age group: 50-69, Anti-Spike level | n = 944, Median: 52.97, IQR: (20.84, 124.9), 5%: 4.01, 10%: 8.73 | n = 249, Median: 250.0, IQR: (250.0, 250.0), 5%: 146.86, 10%: 250.0 | n = 293, Median: 701.0, IQR: (236.0, 3692.0), 5%: 66.94, 10%: 87.28 | n = 815, Median: 15607.0, IQR: (9223.0, 25000.0), 5%: 3946.2, 10%: 5342.6 | n = 1445, Median: 43.28, IQR: (17.82, 105.1), 5%: 3.2, 10%: 6.49 | n = 268, Median: 250.0, IQR: (250.0, 250.0), 5%: 156.69, 10%: 250.0 |
| Age group: 50-69, Weeks since vaccination | Median: 7.0, IQR: (5.0, 9.0) | Median: 4.0, IQR: (2.0, 6.0) | Median: 27.0, IQR: (25.0, 29.0) | Median: 5.0, IQR: (3.0, 7.0) | Median: 6.0, IQR: (5.0, 8.0) | Median: 4.0, IQR: (3.0, 6.0) |
| Age group: 70+, Anti-Spike level | n = 286, Median: 56.87, IQR: (27.99, 108.88), 5%: 7.2, 10%: 13.55 | n = 376, Median: 250.0, IQR: (250.0, 250.0), 5%: 132.8, 10%: 250.0 | n = 46, Median: 3807.5, IQR: (456.25, 12092.5), 5%: 83.45, 10%: 111.15 | n = 907, Median: 12569.0, IQR: (7354.0, 21218.5), 5%: 3128.9, 10%: 4835.0 | n = 6, Median: 72.51, IQR: (18.69, 120.88), 5%: 13.82, 10%: 14.3 | n = 10, Median: 250.0, IQR: (250.0, 250.0), 5%: 124.86, 10%: 227.25 |
| Age group: 70+, Weeks since vaccination | Median: 10.0, IQR: (9.0, 10.0) | Median: 3.0, IQR: (2.0, 4.0) | Median: 31.5, IQR: (29.0, 33.0) | Median: 6.0, IQR: (4.0, 8.0) | Median: 9.0, IQR: (9.0, 9.75) | Median: 2.5, IQR: (2.0, 3.75) |
| Sex: Female, Anti-Spike level | n = 1197, Median: 57.03, IQR: (24.64, 124.9), 5%: 4.78, 10%: 9.88 | n = 660, Median: 250.0, IQR: (250.0, 250.0), 5%: 147.24, 10%: 250.0 | n = 588, Median: 1162.0, IQR: (321.0, 5011.5), 5%: 73.9, 10%: 110.8 | n = 1712, Median: 13803.0, IQR: (8201.5, 23702.75), 5%: 3438.6, 10%: 5027.1 | n = 1062, Median: 45.15, IQR: (19.2, 108.02), 5%: 3.37, 10%: 7.37 | n = 238, Median: 250.0, IQR: (250.0, 250.0), 5%: 245.0, 10%: 250.0 |
| Sex: Female, Weeks since vaccination | Median: 8.0, IQR: (6.0, 9.0) | Median: 3.0, IQR: (2.0, 5.0) | Median: 25.0, IQR: (20.0, 28.0) | Median: 5.0, IQR: (4.0, 8.0) | Median: 6.0, IQR: (5.0, 8.0) | Median: 4.0, IQR: (2.0, 6.0) |
| Sex: Male, Anti-Spike level | n = 178, Median: 38.9, IQR: (15.61, 99.0), 5%: 3.39, 10%: 7.34 | n = 88, Median: 250.0, IQR: (250.0, 250.0), 5%: 196.51, 10%: 250.0 | n = 103, Median: 2349.0, IQR: (410.0, 5971.0), 5%: 66.84, 10%: 132.4 | n = 225, Median: 13220.0, IQR: (7794.0, 21949.0), 5%: 4085.6, 10%: 5325.8 | n = 397, Median: 39.47, IQR: (16.34, 98.81), 5%: 2.82, 10%: 5.93 | n = 46, Median: 250.0, IQR: (250.0, 250.0), 5%: 34.21, 10%: 110.76 |
| Sex: Male, Weeks since vaccination | Median: 8.0, IQR: (6.0, 9.0) | Median: 3.0, IQR: (2.0, 6.0) | Median: 25.0, IQR: (18.0, 28.0) | Median: 6.0, IQR: (4.0, 8.0) | Median: 7.0, IQR: (5.0, 8.0) | Median: 4.0, IQR: (2.25, 6.75) |

| | | | | | | |
|---|---|---|---|---|---|---|
| Ethnicity: White, Anti-Spike level | n = 1339, Median: 53.18, IQR: (22.72, 121.2), 5%: 4.72, 10%: 9.54 | n = 720, Median: 250.0, IQR: (250.0, 250.0), 5%: 147.65, 10%: 250.0 | n = 660, Median: 1272.0, IQR: (324.25, 5262.5), 5%: 73.5, 10%: 108.0 | n = 1875, Median: 13704.0, IQR: (8199.0, 23491.5), 5%: 3497.6, 10%: 5058.0 | n = 1434, Median: 42.82, IQR: (17.81, 104.4), 5%: 3.2, 10%: 6.49 | n = 279, Median: 250.0, IQR: (250.0, 250.0), 5%: 159.66, 10%: 250.0 |
| Ethnicity: White, Weeks since vaccination | Median: 8.0, IQR: (6.0, 9.0) | Median: 3.0, IQR: (2.0, 5.0) | Median: 25.0, IQR: (20.0, 28.0) | Median: 5.0, IQR: (4.0, 8.0) | Median: 6.0, IQR: (5.0, 8.0) | Median: 4.0, IQR: (2.0, 6.0) |
| Ethnicity: Other than White, Anti-Spike level | n = 29, Median: 70.85, IQR: (43.27, 138.8), 5%: 7.61, 10%: 18.61 | n = 19, Median: 250.0, IQR: (250.0, 250.0), 5%: 250.0, 10%: 250.0 | n = 26, Median: 2576.5, IQR: (880.5, 4011.5), 5%: 332.75, 10%: 503.5 | n = 39, Median: 13992.0, IQR: (7064.0, 25000.0), 5%: 3138.3, 10%: 4557.8 | n = 20, Median: 104.45, IQR: (38.22, 250.0), 5%: 12.8, 10%: 15.96 | n = 5, Median: 250.0, IQR: (250.0, 250.0), 5%: 105.53, 10%: 141.65 |
| Ethnicity: Other than White, Weeks since vaccination | Median: 6.0, IQR: (5.0, 8.0) | Median: 4.0, IQR: (3.0, 6.0) | Median: 22.5, IQR: (19.0, 28.0) | Median: 6.0, IQR: (3.5, 9.0) | Median: 7.0, IQR: (5.0, 8.25) | Median: 7.0, IQR: (5.0, 7.0) |
| Local area deprivation, IMD: Least deprived 60% (decile 5-10), Anti-Spike level | n = 1157, Median: 52.32, IQR: (23.04, 119.2), 5%: 5.0, 10%: 10.0 | n = 624, Median: 250.0, IQR: (250.0, 250.0), 5%: 161.07, 10%: 250.0 | n = 539, Median: 1251.0, IQR: (323.5, 5132.5), 5%: 73.85, 10%: 108.0 | n = 1635, Median: 13728.0, IQR: (8248.5, 23222.0), 5%: 3543.4, 10%: 5058.4 | n = 666, Median: 44.08, IQR: (19.06, 110.4), 5%: 3.05, 10%: 6.54 | n = 148, Median: 250.0, IQR: (250.0, 250.0), 5%: 238.0, 10%: 250.0 |
| Local area deprivation, IMD: Least deprived 60% (decile 5-10), Weeks since vaccination | Median: 8.0, IQR: (6.0, 9.0) | Median: 3.0, IQR: (2.0, 5.0) | Median: 25.0, IQR: (20.0, 28.0) | Median: 5.0, IQR: (4.0, 8.0) | Median: 6.0, IQR: (5.0, 7.0) | Median: 4.0, IQR: (3.0, 6.0) |
| Local area deprivation, IMD: Most deprived 40% (decile 1-4), Anti-Spike level | n = 214, Median: 58.87, IQR: (20.36, 147.02), 5%: 3.58, 10%: 6.52 | n = 124, Median: 250.0, IQR: (250.0, 250.0), 5%: 95.82, 10%: 250.0 | n = 151, Median: 1616.0, IQR: (399.5, 5481.0), 5%: 73.7, 10%: 127.0 | n = 298, Median: 13542.5, IQR: (7234.5, 25000.0), 5%: 3058.25, 10%: 4825.6 | n = 305, Median: 45.21, IQR: (18.97, 107.1), 5%: 3.42, 10%: 7.4 | n = 70, Median: 250.0, IQR: (250.0, 250.0), 5%: 250.0, 10%: 250.0 |
| Local area deprivation, IMD: Most deprived 40% (decile 1-4), Weeks since vaccination | Median: 8.0, IQR: (6.0, 9.0) | Median: 3.0, IQR: (2.0, 5.0) | Median: 25.0, IQR: (19.5, 29.0) | Median: 5.0, IQR: (3.0, 8.0) | Median: 6.0, IQR: (5.0, 7.0) | Median: 3.0, IQR: (2.0, 6.0) |
| Highest educational attainment: NVQ level 3 or lower, Anti-Spike level | n = 497, Median: 55.56, IQR: (24.64, 119.8), 5%: 5.39, 10%: 10.8 | n = 275, Median: 250.0, IQR: (250.0, 250.0), 5%: 62.1, 10%: 181.34 | n = 171, Median: 951.0, IQR: (296.0, 5211.5), 5%: 64.05, 10%: 100.0 | n = 763, Median: 13768.0, IQR: (8100.5, 23719.5), 5%: 3388.4, 10%: 4898.0 | n = 936, Median: 45.78, IQR: (17.99, 115.85), 5%: 2.76, 10%: 5.62 | n = 210, Median: 250.0, IQR: (250.0, 250.0), 5%: 163.08, 10%: 250.0 |
| Highest educational attainment: NVQ level 3 or lower, Weeks since vaccination | Median: 8.0, IQR: (6.0, 10.0) | Median: 3.0, IQR: (2.0, 5.0) | Median: 27.0, IQR: (24.5, 30.0) | Median: 5.0, IQR: (4.0, 8.0) | Median: 6.0, IQR: (5.0, 8.0) | Median: 4.0, IQR: (3.0, 6.0) |
| Highest educational attainment: NVQ level 4 or higher, Anti-Spike level | n = 709, Median: 52.58, IQR: (22.76, 117.8), 5%: 4.57, 10%: 9.63 | n = 377, Median: 250.0, IQR: (250.0, 250.0), 5%: 226.0, 10%: 250.0 | n = 354, Median: 1485.0, IQR: (353.5, 5717.5), 5%: 73.76, 10%: 106.6 | n = 969, Median: 13589.0, IQR: (8089.0, 23543.0), 5%: 3570.2, 10%: 5072.0 | n = 452, Median: 39.61, IQR: (18.4, 94.4), 5%: 5.08, 10%: 8.56 | n = 60, Median: 250.0, IQR: (250.0, 250.0), 5%: 148.9, 10%: 244.08 |
| Highest educational attainment: NVQ level 4 or higher, Weeks since vaccination | Median: 8.0, IQR: (6.0, 9.0) | Median: 3.0, IQR: (2.0, 5.0) | Median: 25.0, IQR: (20.0, 28.0) | Median: 5.0, IQR: (3.0, 8.0) | Median: 7.0, IQR: (5.0, 8.0) | Median: 4.0, IQR: (2.0, 7.0) |
| First vaccination received: AZD1222, Anti-Spike level | n = 1103, Median: 48.45, IQR: (19.8, 116.05), 5%: 3.89, 10%: 7.89 | | | | n = 1235, Median: 39.26, IQR: (16.42, 103.45), 5%: 2.82, 10%: 5.8 | n = 50, Median: 250.0, IQR: (163.6, 250.0), 5%: 22.95, 10%: 79.62 |
| First vaccination received: AZD1222, Weeks since vaccination | Median: 7.0, IQR: (5.0, 9.0) | | | | Median: 6.0, IQR: (5.0, 7.0) | Median: 3.0, IQR: (2.0, 4.0) |

| | | | | | | |
|---|---|---|---|---|---|---|
| First vaccination received: BNT162b2, Anti-Spike level | n = 266, Median: 74.33, IQR: (35.96, 145.3), 5%: 13.41, 10%: 19.29 | | | | n = 224, Median: 59.66, IQR: (31.04, 118.18), 5%: 9.29, 10%: 16.14 | n = 234, Median: 250.0, IQR: (250.0, 250.0), 5%: 250.0, 10%: 250.0 |
| First vaccination received: BNT162b2, Weeks since vaccination | Median: 9.0, IQR: (8.0, 10.0) | | | | Median: 8.0, IQR: (7.0, 9.0) | Median: 4.0, IQR: (3.0, 7.0) |
| First vaccination received: Other, Anti-Spike level | n = 5, Median: 57.86, IQR: (42.05, 107.0), 5%: 39.03, 10%: 39.79 | | | | | |
| First vaccination received: Other, Weeks since vaccination | Median: 10.0, IQR: (8.0, 10.0) | | | | | |
| Second vaccination received: AZD1222, Anti-Spike level | | n = 212, Median: 250.0, IQR: (250.0, 250.0), 5%: 91.47, 10%: 171.93 | n = 411, Median: 459.0, IQR: (216.5, 2721.0), 5%: 60.45, 10%: 83.4 | n = 1065, Median: 14358.0, IQR: (8523.0, 23950.0), 5%: 3636.4, 10%: 5260.2 | | |
| Second vaccination received: AZD1222, Weeks since vaccination | | Median: 3.0, IQR: (2.0, 3.25) | Median: 26.0, IQR: (24.0, 28.0) | Median: 5.0, IQR: (3.0, 7.0) | | |
| Second vaccination received: BNT162b2, Anti-Spike level | | n = 532, Median: 250.0, IQR: (250.0, 250.0), 5%: 250.0, 10%: 250.0 | n = 241, Median: 3174.0, IQR: (1424.0, 6406.0), 5%: 571.0, 10%: 731.0 | n = 858, Median: 13118.0, IQR: (7619.25, 22570.5), 5%: 3312.9, 10%: 4782.6 | | |
| Second vaccination received: BNT162b2, Weeks since vaccination | | Median: 4.0, IQR: (2.75, 6.0) | Median: 20.0, IQR: (17.0, 30.0) | Median: 7.0, IQR: (4.0, 9.0) | | |
| Second vaccination received: Other, Anti-Spike level | | n < 5, Median: 250.0, IQR: (229.42, 250.0), 5%: 180.04, 10%: 192.39 | n = 39, Median: 4222.0, IQR: (3107.0, 7033.0), 5%: 1382.9, 10%: 2231.2 | n = 11, Median: 14195.0, IQR: (10306.5, 21298.0), 5%: 7422.0, 10%: 8391.0 | | |
| Second vaccination received: Other, Weeks since vaccination | | Median: 8.0, IQR: (5.5, 10.0) | Median: 19.0, IQR: (16.0, 21.5) | Median: 5.0, IQR: (4.0, 7.5) | | |
| Third vaccination received: mRNA-1273, Anti-Spike level | | | | n = 203, Median: 22154.0, IQR: (14025.0, 25000.0), 5%: 6129.1, 10%: 8709.4 | | |
| Third vaccination received: mRNA-1273, Weeks since vaccination | | | | Median: 3.0, IQR: (2.0, 5.0) | | |
| Third vaccination received: BNT162b2, Anti-Spike level | | | | n = 1677, Median: 13170.0, IQR: (7743.0, 22100.0), 5%: 3262.0, 10%: 4865.4 | | |
| Third vaccination received: BNT162b2, Weeks since vaccination | | | | Median: 6.0, IQR: (4.0, 8.0) | | |
| Third vaccination received: Other, Anti-Spike level | | | | n = 23, Median: 16475.0, IQR: (8934.0, 23189.5), 5%: 1489.5, 10%: 6515.6 | | |

| | | | | | | |
|---|---|---|---|---|---|---|
| Third vaccination received: Other, Weeks since vaccination | | | | Median: 5.0, IQR: (4.0, 7.5) | | |
| SARS-CoV-2 infection status (serology-based) at time of antibody testing: No evidence of natural infection, Anti-Spike level | n = 1071, Median: 43.94, IQR: (19.72, 87.77), 5%: 4.04, 10%: 8.7 | n = 591, Median: 250.0, IQR: (250.0, 250.0), 5%: 136.35, 10%: 250.0 | n = 444, Median: 603.5, IQR: (237.75, 2460.0), 5%: 61.08, 10%: 84.41 | n = 1471, Median: 12883.0, IQR: (7551.0, 21130.5), 5%: 3227.0, 10%: 4828.0 | n = 1305, Median: 38.2, IQR: (16.29, 83.52), 5%: 2.93, 10%: 6.07 | n = 252, Median: 250.0, IQR: (250.0, 250.0), 5%: 152.24, 10%: 250.0 |
| SARS-CoV-2 infection status (serology-based) at time of antibody testing: No evidence of natural infection, Weeks since vaccination | Median: 8.0, IQR: (6.0, 9.0) | Median: 3.0, IQR: (2.0, 5.0) | Median: 25.0, IQR: (20.0, 28.0) | Median: 5.0, IQR: (4.0, 8.0) | Median: 6.0, IQR: (5.0, 8.0) | Median: 4.0, IQR: (2.0, 6.0) |
| SARS-CoV-2 infection status (serology-based) at time of antibody testing: Evidence of natural infection, Anti-Spike level | n = 304, Median: 250.0, IQR: (52.84, 250.0), 5%: 7.68, 10%: 18.84 | n = 157, Median: 250.0, IQR: (250.0, 250.0), 5%: 250.0, 10%: 250.0 | n = 245, Median: 5051.0, IQR: (1750.0, 13098.0), 5%: 247.4, 10%: 465.8 | n = 464, Median: 18238.0, IQR: (10392.25, 25000.0), 5%: 4690.5, 10%: 6661.0 | n = 133, Median: 250.0, IQR: (250.0, 250.0), 5%: 155.18, 10%: 250.0 | n = 31, Median: 250.0, IQR: (250.0, 250.0), 5%: 250.0, 10%: 250.0 |
| SARS-CoV-2 infection status (serology-based) at time of antibody testing: Evidence of natural infection, Weeks since vaccination | Median: 8.0, IQR: (6.0, 9.0) | Median: 3.0, IQR: (2.0, 5.0) | Median: 25.0, IQR: (20.0, 28.0) | Median: 5.0, IQR: (3.0, 8.0) | Median: 6.0, IQR: (5.0, 8.0) | Median: 4.0, IQR: (3.0, 6.0) |
| SARS-CoV-2 infection status (self-reported), Q2: No infection, Anti-Spike level | n = 921, Median: 46.19, IQR: (20.35, 92.63), 5%: 3.69, 10%: 8.42 | n = 551, Median: 250.0, IQR: (250.0, 250.0), 5%: 119.45, 10%: 250.0 | n = 462, Median: 989.5, IQR: (293.0, 4610.75), 5%: 66.45, 10%: 100.1 | n = 1399, Median: 13408.0, IQR: (7612.0, 23167.5), 5%: 3245.4, 10%: 4823.6 | n = 1105, Median: 40.23, IQR: (17.09, 94.07), 5%: 3.03, 10%: 6.21 | n = 200, Median: 250.0, IQR: (250.0, 250.0), 5%: 154.52, 10%: 250.0 |
| SARS-CoV-2 infection status (self-reported), Q2: No infection, Weeks since vaccination | Median: 8.0, IQR: (6.0, 10.0) | Median: 3.0, IQR: (2.0, 5.0) | Median: 25.0, IQR: (20.0, 28.0) | Median: 6.0, IQR: (4.0, 8.0) | Median: 6.0, IQR: (5.0, 8.0) | Median: 4.0, IQR: (2.0, 6.0) |
| SARS-CoV-2 infection status (self-reported), Q2: Unsure, Anti-Spike level | n = 43, Median: 43.81, IQR: (17.67, 107.7), 5%: 8.96, 10%: 10.85 | n = 9, Median: 250.0, IQR: (250.0, 250.0), 5%: 190.06, 10%: 230.02 | n = 12, Median: 826.5, IQR: (328.75, 3462.0), 5%: 201.15, 10%: 253.3 | n = 30, Median: 20072.5, IQR: (14218.75, 25000.0), 5%: 6379.3, 10%: 7422.6 | | |
| SARS-CoV-2 infection status (self-reported), Q2: Unsure, Weeks since vaccination | Median: 8.0, IQR: (5.5, 10.0) | Median: 3.0, IQR: (3.0, 7.0) | Median: 24.0, IQR: (16.75, 25.5) | Median: 3.5, IQR: (3.0, 6.0) | | |
| SARS-CoV-2 infection status (self-reported), Q2: Suspected case, Anti-Spike level | n = 183, Median: 50.41, IQR: (20.83, 101.35), 5%: 5.1, 10%: 7.92 | n = 67, Median: 250.0, IQR: (250.0, 250.0), 5%: 239.29, 10%: 250.0 | n = 81, Median: 705.0, IQR: (250.0, 5873.0), 5%: 62.7, 10%: 76.2 | n = 197, Median: 13677.0, IQR: (8263.0, 23933.0), 5%: 3830.0, 10%: 5050.8 | n = 240, Median: 55.47, IQR: (19.76, 153.4), 5%: 3.14, 10%: 7.03 | n = 57, Median: 250.0, IQR: (250.0, 250.0), 5%: 207.89, 10%: 250.0 |
| SARS-CoV-2 infection status (self-reported), Q2: Suspected case, Weeks since vaccination | Median: 8.0, IQR: (5.5, 9.0) | Median: 3.0, IQR: (2.0, 5.0) | Median: 26.0, IQR: (21.0, 29.0) | Median: 6.0, IQR: (4.0, 8.0) | Median: 6.0, IQR: (5.0, 8.0) | Median: 3.0, IQR: (3.0, 7.0) |
| SARS-CoV-2 infection status (self-reported), Q2: Confirmed case, Anti-Spike level | n = 218, Median: 250.0, IQR: (57.28, 250.0), 5%: 11.66, 10%: 23.07 | n = 112, Median: 250.0, IQR: (250.0, 250.0), 5%: 250.0, 10%: 250.0 | n = 107, Median: 3390.0, IQR: (1253.5, 6194.5), 5%: 302.0, 10%: 431.2 | n = 256, Median: 15752.0, IQR: (10174.75, 24951.25), 5%: 5717.75, 10%: 6730.0 | n = 29, Median: 250.0, IQR: (250.0, 250.0), 5%: 107.84, 10%: 250.0 | n = 11, Median: 250.0, IQR: (250.0, 250.0), 5%: 250.0, 10%: 250.0 |

| | | | | | | |
|---|---|---|---|---|---|---|
| SARS-CoV-2 infection status (self-reported), Q2: Confirmed case, Weeks since vaccination | Median: 7.0, IQR: (5.0, 9.0) | Median: 4.0, IQR: (3.0, 6.0) | Median: 25.0, IQR: (20.0, 29.0) | Median: 5.0, IQR: (3.0, 8.0) | Median: 5.0, IQR: (5.0, 7.0) | Median: 3.0, IQR: (2.0, 5.0) |
| SARS-CoV-2 infection status (self-reported), Q4: No infection, Anti-Spike level | n = 819, Median: 47.06, IQR: (21.18, 94.3), 5%: 4.06, 10%: 8.88 | n = 515, Median: 250.0, IQR: (250.0, 250.0), 5%: 132.12, 10%: 250.0 | n = 381, Median: 614.0, IQR: (238.0, 2624.0), 5%: 63.6, 10%: 87.1 | n = 1310, Median: 12876.0, IQR: (7374.25, 21687.0), 5%: 3123.9, 10%: 4737.3 | | |
| SARS-CoV-2 infection status (self-reported), Q4: No infection, Weeks since vaccination | Median: 8.0, IQR: (6.0, 10.0) | Median: 3.0, IQR: (2.0, 5.0) | Median: 25.0, IQR: (20.0, 28.0) | Median: 6.0, IQR: (4.0, 8.0) | | |
| SARS-CoV-2 infection status (self-reported), Q4: Unsure, Anti-Spike level | n = 67, Median: 41.75, IQR: (18.82, 86.56), 5%: 4.61, 10%: 10.17 | n = 18, Median: 250.0, IQR: (250.0, 250.0), 5%: 137.65, 10%: 220.03 | n = 30, Median: 646.5, IQR: (272.75, 3521.0), 5%: 105.81, 10%: 138.2 | n = 65, Median: 17892.0, IQR: (10187.0, 25000.0), 5%: 3924.6, 10%: 6240.4 | | |
| SARS-CoV-2 infection status (self-reported), Q4: Unsure, Weeks since vaccination | Median: 8.0, IQR: (5.5, 10.0) | Median: 4.0, IQR: (3.0, 6.75) | Median: 24.5, IQR: (21.0, 27.75) | Median: 4.0, IQR: (3.0, 6.0) | | |
| SARS-CoV-2 infection status (self-reported), Q4: Suspected case, Anti-Spike level | n = 183, Median: 48.54, IQR: (19.56, 101.35), 5%: 5.04, 10%: 7.11 | n = 70, Median: 250.0, IQR: (250.0, 250.0), 5%: 241.58, 10%: 250.0 | n = 78, Median: 715.5, IQR: (244.0, 3822.25), 5%: 62.31, 10%: 75.51 | n = 204, Median: 13453.5, IQR: (8192.0, 23315.0), 5%: 3739.0, 10%: 5033.0 | | |
| SARS-CoV-2 infection status (self-reported), Q4: Suspected case, Weeks since vaccination | Median: 8.0, IQR: (6.0, 9.0) | Median: 3.0, IQR: (2.0, 5.0) | Median: 25.0, IQR: (20.25, 27.75) | Median: 5.5, IQR: (3.0, 8.0) | | |
| SARS-CoV-2 infection status (self-reported), Q4: Confirmed case, Anti-Spike level | n = 306, Median: 150.0, IQR: (40.05, 250.0), 5%: 7.26, 10%: 12.61 | n = 145, Median: 250.0, IQR: (250.0, 250.0), 5%: 250.0, 10%: 250.0 | n = 202, Median: 5851.5, IQR: (2217.5, 15805.5), 5%: 395.0, 10%: 813.3 | n = 357, Median: 18091.0, IQR: (10524.0, 25000.0), 5%: 5743.2, 10%: 7116.8 | | |
| SARS-CoV-2 infection status (self-reported), Q4: Confirmed case, Weeks since vaccination | Median: 7.0, IQR: (5.0, 9.0) | Median: 4.0, IQR: (3.0, 6.0) | Median: 25.0, IQR: (21.0, 29.0) | Median: 5.0, IQR: (3.0, 8.0) | | |
| Frailty Index: Healthy, Anti-Spike level | n = 388, Median: 52.1, IQR: (24.83, 130.9), 5%: 5.06, 10%: 10.51 | n = 189, Median: 250.0, IQR: (250.0, 250.0), 5%: 158.22, 10%: 250.0 | n = 241, Median: 1364.0, IQR: (317.0, 5154.0), 5%: 66.4, 10%: 97.7 | n = 503, Median: 14607.0, IQR: (8607.5, 24247.0), 5%: 4473.4, 10%: 5464.6 | | |
| Frailty Index: Healthy, Weeks since vaccination | Median: 7.0, IQR: (6.0, 9.0) | Median: 4.0, IQR: (3.0, 5.0) | Median: 24.0, IQR: (19.0, 27.0) | Median: 5.0, IQR: (3.0, 7.0) | | |
| Frailty Index: Pre-frail, Anti-Spike level | n = 604, Median: 52.48, IQR: (22.08, 114.2), 5%: 4.74, 10%: 9.87 | n = 320, Median: 250.0, IQR: (250.0, 250.0), 5%: 178.23, 10%: 250.0 | n = 185, Median: 1127.0, IQR: (307.0, 5172.0), 5%: 74.86, 10%: 125.0 | n = 914, Median: 13481.5, IQR: (8037.5, 22984.75), 5%: 3441.1, 10%: 5063.2 | | |
| Frailty Index: Pre-frail, Weeks since vaccination | Median: 8.0, IQR: (6.0, 10.0) | Median: 3.0, IQR: (2.0, 5.0) | Median: 26.0, IQR: (23.0, 30.0) | Median: 5.0, IQR: (4.0, 8.0) | | |

| | | | | | | |
|---|---|---|---|---|---|---|
| Frailty Index: Frail, Anti-Spike level | n = 111, Median: 55.46, IQR: (25.4, 105.4), 5%: 2.69, 10%: 7.75 | n = 90, Median: 250.0, IQR: (250.0, 250.0), 5%: 215.44, 10%: 250.0 | n = 45, Median: 1022.0, IQR: (438.0, 5767.0), 5%: 93.04, 10%: 138.8 | n = 192, Median: 13585.5, IQR: (7378.75, 22779.5), 5%: 3160.1, 10%: 4540.8 | | |
| Frailty Index: Frail, Weeks since vaccination | Median: 8.0, IQR: (6.5, 9.0) | Median: 3.0, IQR: (2.0, 5.0) | Median: 29.0, IQR: (26.0, 32.0) | Median: 6.0, IQR: (4.0, 8.0) | | |
| Frailty Index: Very frail, Anti-Spike level | n = 35, Median: 48.9, IQR: (20.84, 120.15), 5%: 4.83, 10%: 9.74 | n = 23, Median: 250.0, IQR: (119.25, 250.0), 5%: 7.88, 10%: 36.36 | n = 7, Median: 2444.0, IQR: (884.0, 5069.5), 5%: 469.8, 10%: 528.6 | n = 56, Median: 12848.5, IQR: (6776.75, 22534.0), 5%: 2372.5, 10%: 4015.0 | | |
| Frailty Index: Very frail, Weeks since vaccination | Median: 9.0, IQR: (7.0, 10.0) | Median: 4.0, IQR: (2.0, 5.5) | Median: 28.0, IQR: (26.0, 28.5) | Median: 6.0, IQR: (4.75, 8.0) | | |
| Frailty (PRISMA-7): Healthy, Anti-Spike level | | | | | n = 1419, Median: 43.42, IQR: (17.89, 107.55), 5%: 3.24, 10%: 6.55 | n = 274, Median: 250.0, IQR: (250.0, 250.0), 5%: 165.2, 10%: 250.0 |
| Frailty (PRISMA-7): Healthy, Weeks since vaccination | | | | | Median: 6.0, IQR: (5.0, 8.0) | Median: 4.0, IQR: (2.0, 6.0) |
| Frailty (PRISMA-7): Frail, Anti-Spike level | | | | | n = 40, Median: 42.0, IQR: (19.33, 70.04), 5%: 2.0, 10%: 4.09 | n = 10, Median: 250.0, IQR: (143.95, 250.0), 5%: 59.11, 10%: 78.79 |
| Frailty (PRISMA-7): Frail, Weeks since vaccination | | | | | Median: 7.0, IQR: (6.0, 8.0) | Median: 3.0, IQR: (2.25, 4.0) |
| Advised on "Shielded Patient List": No, Anti-Spike level | n = 1292, Median: 54.74, IQR: (23.28, 120.6), 5%: 5.04, 10%: 10.0 | n = 662, Median: 250.0, IQR: (250.0, 250.0), 5%: 179.03, 10%: 250.0 | n = 668, Median: 1316.5, IQR: (333.25, 5187.25), 5%: 73.9, 10%: 112.7 | n = 1746, Median: 13997.5, IQR: (8358.5, 23888.5), 5%: 3970.5, 10%: 5272.0 | n = 1398, Median: 43.82, IQR: (18.26, 107.0), 5%: 3.4, 10%: 6.78 | n = 254, Median: 250.0, IQR: (250.0, 250.0), 5%: 183.01, 10%: 250.0 |
| Advised on "Shielded Patient List": No, Weeks since vaccination | Median: 8.0, IQR: (6.0, 9.0) | Median: 3.0, IQR: (2.0, 5.0) | Median: 25.0, IQR: (20.0, 28.0) | Median: 5.0, IQR: (3.25, 8.0) | Median: 6.0, IQR: (5.0, 8.0) | Median: 4.0, IQR: (2.0, 6.0) |
| Advised on "Shielded Patient List": Yes, Anti-Spike level | n = 82, Median: 43.78, IQR: (13.26, 137.85), 5%: 0.56, 10%: 2.47 | n = 86, Median: 250.0, IQR: (250.0, 250.0), 5%: 24.56, 10%: 56.74 | n = 23, Median: 1801.0, IQR: (385.5, 5767.5), 5%: 62.34, 10%: 113.96 | n = 190, Median: 11244.0, IQR: (6057.5, 19538.0), 5%: 754.5, 10%: 1989.7 | n = 45, Median: 40.75, IQR: (9.39, 64.46), 5%: 0.4, 10%: 0.76 | n = 22, Median: 250.0, IQR: (250.0, 250.0), 5%: 2.35, 10%: 43.13 |
| Advised on "Shielded Patient List": Yes, Weeks since vaccination | Median: 9.0, IQR: (7.0, 10.0) | Median: 3.0, IQR: (2.0, 5.0) | Median: 29.0, IQR: (26.0, 32.0) | Median: 6.5, IQR: (4.0, 8.0) | Median: 8.0, IQR: (7.0, 9.0) | Median: 4.0, IQR: (3.0, 6.75) |
| Prescribed immunosuppressant medication: No, Anti-Spike level | n = 846, Median: 52.0, IQR: (22.6, 117.18), 5%: 5.03, 10%: 10.04 | n = 476, Median: 250.0, IQR: (250.0, 250.0), 5%: 165.75, 10%: 250.0 | n = 357, Median: 1161.0, IQR: (311.0, 4883.0), 5%: 73.48, 10%: 100.88 | n = 1270, Median: 14144.0, IQR: (8210.0, 23532.5), 5%: 3724.75, 10%: 5058.3 | | |
| Prescribed immunosuppressant medication: No, Weeks since vaccination | Median: 8.0, IQR: (6.0, 9.0) | Median: 3.0, IQR: (2.0, 5.0) | Median: 25.0, IQR: (21.0, 28.0) | Median: 5.0, IQR: (4.0, 8.0) | | |
| Prescribed immunosuppressant medication: Yes, Anti-Spike level | n = 117, Median: 47.94, IQR: (27.12, 112.0), 5%: 3.48, 10%: 10.0 | n = 64, Median: 250.0, IQR: (250.0, 250.0), 5%: 7.32, 10%: 44.74 | n = 37, Median: 1938.0, IQR: (851.0, 7639.0), 5%: 106.0, 10%: 258.4 | n = 192, Median: 12432.0, IQR: (7046.0, 20654.0), 5%: 1371.1, 10%: 3589.1 | | |

| | | | | | | |
|---|---|---|---|---|---|---|
| Prescribed immunosuppressant medication: Yes, Weeks since vaccination | Median: 9.0, IQR: (7.0, 10.0) | Median: 3.0, IQR: (2.0, 4.25) | Median: 28.0, IQR: (24.0, 32.0) | Median: 5.0, IQR: (4.0, 8.0) | | |
| Immunocompromised: No, Anti-Spike level | | | | | n = 1053, Median: 42.26, IQR: (18.25, 101.0), 5%: 3.39, 10%: 6.59 | n = 198, Median: 250.0, IQR: (250.0, 250.0), 5%: 189.29, 10%: 250.0 |
| Immunocompromised: No, Weeks since vaccination | | | | | Median: 6.0, IQR: (5.0, 8.0) | Median: 4.0, IQR: (2.0, 6.0) |
| Immunocompromised: Yes, Anti-Spike level | | | | | n = 39, Median: 38.89, IQR: (6.9, 61.0), 5%: 0.68, 10%: 0.91 | n = 15, Median: 250.0, IQR: (250.0, 250.0), 5%: 27.72, 10%: 90.3 |
| Immunocompromised: Yes, Weeks since vaccination | | | | | Median: 8.0, IQR: (7.0, 9.0) | Median: 4.0, IQR: (2.0, 4.0) |
| Self-rated health: Poor, Fair, Anti-Spike level | n = 134, Median: 57.2, IQR: (20.61, 118.72), 5%: 1.75, 10%: 3.21 | n = 60, Median: 250.0, IQR: (250.0, 250.0), 5%: 20.31, 10%: 99.87 | n = 42, Median: 1855.5, IQR: (593.5, 16280.0), 5%: 141.4, 10%: 244.5 | n = 168, Median: 13431.0, IQR: (6789.0, 24415.75), 5%: 1237.35, 10%: 4072.7 | n = 137, Median: 39.75, IQR: (20.29, 101.0), 5%: 2.27, 10%: 5.67 | n = 28, Median: 250.0, IQR: (250.0, 250.0), 5%: 77.94, 10%: 140.84 |
| Self-rated health: Poor, Fair, Weeks since vaccination | Median: 9.0, IQR: (7.0, 10.0) | Median: 3.0, IQR: (2.0, 4.25) | Median: 27.0, IQR: (24.25, 29.0) | Median: 6.0, IQR: (4.0, 8.0) | Median: 6.0, IQR: (5.0, 8.0) | Median: 3.5, IQR: (2.75, 7.0) |
| Self-rated health: Good, Very Good, Excellent, Anti-Spike level | n = 1230, Median: 53.1, IQR: (23.0, 121.7), 5%: 5.14, 10%: 10.02 | n = 677, Median: 250.0, IQR: (250.0, 250.0), 5%: 174.24, 10%: 250.0 | n = 614, Median: 1308.5, IQR: (322.75, 5091.75), 5%: 72.67, 10%: 105.3 | n = 1703, Median: 13851.0, IQR: (8243.0, 23522.0), 5%: 3609.5, 10%: 5111.4 | n = 1322, Median: 43.68, IQR: (17.75, 107.18), 5%: 3.37, 10%: 6.52 | n = 255, Median: 250.0, IQR: (250.0, 250.0), 5%: 161.88, 10%: 250.0 |
| Self-rated health: Good, Very Good, Excellent, Weeks since vaccination | Median: 8.0, IQR: (6.0, 9.0) | Median: 3.0, IQR: (2.0, 5.0) | Median: 25.0, IQR: (20.0, 28.0) | Median: 5.0, IQR: (4.0, 8.0) | Median: 6.0, IQR: (5.0, 8.0) | Median: 4.0, IQR: (2.0, 6.0) |
| Number of selected comorbidities: 0, Anti-Spike level | n = 604, Median: 52.52, IQR: (25.51, 125.1), 5%: 4.81, 10%: 9.99 | n = 267, Median: 250.0, IQR: (250.0, 250.0), 5%: 223.01, 10%: 250.0 | n = 308, Median: 1459.0, IQR: (312.5, 5848.25), 5%: 72.04, 10%: 104.1 | n = 789, Median: 13574.0, IQR: (8251.0, 23727.0), 5%: 3872.0, 10%: 5068.8 | n = 744, Median: 42.08, IQR: (17.7, 97.25), 5%: 3.37, 10%: 6.3 | n = 138, Median: 250.0, IQR: (250.0, 250.0), 5%: 189.89, 10%: 250.0 |
| Number of selected comorbidities: 0, Weeks since vaccination | Median: 8.0, IQR: (6.0, 9.0) | Median: 3.0, IQR: (2.0, 5.0) | Median: 25.0, IQR: (20.0, 28.0) | Median: 5.0, IQR: (3.0, 7.0) | Median: 6.0, IQR: (5.0, 7.0) | Median: 3.0, IQR: (2.0, 6.0) |
| Number of selected comorbidities: 1+, Anti-Spike level | n = 452, Median: 51.8, IQR: (20.14, 114.2), 5%: 4.41, 10%: 9.87 | n = 288, Median: 250.0, IQR: (250.0, 250.0), 5%: 97.91, 10%: 248.59 | n = 144, Median: 850.5, IQR: (354.0, 3639.0), 5%: 84.35, 10%: 131.6 | n = 717, Median: 13768.0, IQR: (7833.0, 23953.0), 5%: 3135.4, 10%: 4869.0 | n = 314, Median: 41.8, IQR: (19.28, 108.02), 5%: 3.01, 10%: 8.0 | n = 67, Median: 250.0, IQR: (250.0, 250.0), 5%: 183.75, 10%: 250.0 |
| Number of selected comorbidities: 1+, Weeks since vaccination | Median: 8.0, IQR: (6.0, 10.0) | Median: 3.0, IQR: (2.0, 5.0) | Median: 26.0, IQR: (24.0, 30.0) | Median: 6.0, IQR: (4.0, 8.0) | Median: 7.0, IQR: (5.0, 8.0) | Median: 4.0, IQR: (3.0, 6.5) |
| Comorbidity: Anxiety or Stress Disorder: No, Anti-Spike level | n = 894, Median: 52.29, IQR: (24.12, 125.5), 5%: 4.96, 10%: 10.0 | n = 495, Median: 250.0, IQR: (250.0, 250.0), 5%: 144.44, 10%: 250.0 | n = 371, Median: 1301.0, IQR: (317.5, 5457.5), 5%: 71.55, 10%: 97.7 | n = 1335, Median: 13574.0, IQR: (8055.0, 22890.0), 5%: 3448.1, 10%: 4974.2 | n = 969, Median: 42.55, IQR: (17.74, 100.3), 5%: 3.1, 10%: 6.14 | n = 195, Median: 250.0, IQR: (250.0, 250.0), 5%: 160.26, 10%: 250.0 |
| Comorbidity: Anxiety or Stress Disorder: No, Weeks since vaccination | Median: 8.0, IQR: (6.0, 10.0) | Median: 3.0, IQR: (2.0, 5.0) | Median: 25.0, IQR: (21.0, 29.0) | Median: 5.0, IQR: (4.0, 8.0) | Median: 6.0, IQR: (5.0, 8.0) | Median: 4.0, IQR: (2.0, 6.0) |

| | | | | | | |
|---|---|---|---|---|---|---|
| Comorbidity: Anxiety or Stress Disorder: Yes, Anti-Spike level | n = 165, Median: 60.86, IQR: (21.39, 116.8), 5%: 4.28, 10%: 9.5 | n = 81, Median: 250.0, IQR: (250.0, 250.0), 5%: 174.9, 10%: 250.0 | n = 70, Median: 790.0, IQR: (396.5, 3271.0), 5%: 101.35, 10%: 137.5 | n = 213, Median: 15039.0, IQR: (9191.0, 25000.0), 5%: 4215.2, 10%: 5718.2 | n = 114, Median: 39.25, IQR: (19.28, 101.38), 5%: 5.71, 10%: 9.7 | n = 16, Median: 250.0, IQR: (250.0, 250.0), 5%: 227.55, 10%: 250.0 |
| Comorbidity: Anxiety or Stress Disorder: Yes, Weeks since vaccination | Median: 8.0, IQR: (6.0, 9.0) | Median: 3.0, IQR: (2.0, 5.0) | Median: 26.0, IQR: (23.0, 28.0) | Median: 6.0, IQR: (4.0, 7.0) | Median: 6.5, IQR: (6.0, 8.0) | Median: 5.0, IQR: (3.75, 8.0) |
| Comorbidity: Depression: No, Anti-Spike level | n = 917, Median: 54.01, IQR: (24.28, 126.5), 5%: 4.99, 10%: 10.1 | n = 504, Median: 250.0, IQR: (250.0, 250.0), 5%: 151.27, 10%: 250.0 | n = 383, Median: 1301.0, IQR: (321.5, 5602.0), 5%: 67.54, 10%: 98.56 | n = 1353, Median: 13700.0, IQR: (8160.0, 23484.0), 5%: 3685.6, 10%: 5058.2 | n = 1037, Median: 42.55, IQR: (17.82, 100.3), 5%: 3.26, 10%: 6.5 | n = 202, Median: 250.0, IQR: (250.0, 250.0), 5%: 160.32, 10%: 250.0 |
| Comorbidity: Depression: No, Weeks since vaccination | Median: 8.0, IQR: (6.0, 10.0) | Median: 3.0, IQR: (2.0, 5.0) | Median: 25.0, IQR: (21.0, 29.0) | Median: 5.0, IQR: (4.0, 8.0) | Median: 6.0, IQR: (5.0, 8.0) | Median: 4.0, IQR: (2.0, 6.0) |
| Comorbidity: Depression: Yes, Anti-Spike level | n = 134, Median: 48.42, IQR: (20.4, 108.5), 5%: 5.1, 10%: 9.93 | n = 70, Median: 250.0, IQR: (250.0, 250.0), 5%: 98.54, 10%: 170.31 | n = 54, Median: 1157.5, IQR: (377.25, 3339.25), 5%: 121.2, 10%: 242.8 | n = 180, Median: 14426.5, IQR: (7579.75, 24805.5), 5%: 2673.1, 10%: 4952.9 | n = 55, Median: 39.73, IQR: (23.68, 100.94), 5%: 2.92, 10%: 4.4 | n = 9, Median: 250.0, IQR: (250.0, 250.0), 5%: 250.0, 10%: 250.0 |
| Comorbidity: Depression: Yes, Weeks since vaccination | Median: 8.0, IQR: (6.0, 9.0) | Median: 3.0, IQR: (2.0, 5.0) | Median: 26.0, IQR: (22.0, 28.75) | Median: 5.0, IQR: (3.0, 7.0) | Median: 6.0, IQR: (5.0, 7.0) | Median: 4.0, IQR: (3.0, 6.0) |
| Anxiety (HADS) score up to Q2: 8-10, mild, Anti-Spike level | n = 298, Median: 52.36, IQR: (24.94, 120.52), 5%: 6.87, 10%: 10.58 | n = 153, Median: 250.0, IQR: (250.0, 250.0), 5%: 164.98, 10%: 250.0 | n = 143, Median: 808.0, IQR: (318.0, 4259.0), 5%: 94.66, 10%: 146.6 | n = 392, Median: 13739.5, IQR: (7930.5, 24174.25), 5%: 3611.8, 10%: 5222.6 | | |
| Anxiety (HADS) score up to Q2: 8-10, mild, Weeks since vaccination | Median: 8.0, IQR: (6.0, 9.0) | Median: 4.0, IQR: (3.0, 5.0) | Median: 25.0, IQR: (20.5, 27.5) | Median: 5.0, IQR: (4.0, 8.0) | | |
| Anxiety (HADS) score up to Q2: 11+, moderate, severe, Anti-Spike level | n = 262, Median: 58.33, IQR: (24.18, 125.35), 5%: 4.42, 10%: 10.64 | n = 120, Median: 250.0, IQR: (250.0, 250.0), 5%: 156.53, 10%: 250.0 | n = 137, Median: 1750.0, IQR: (373.0, 6057.0), 5%: 65.62, 10%: 101.8 | n = 276, Median: 14713.0, IQR: (8193.75, 22705.0), 5%: 3705.5, 10%: 5261.0 | | |
| Anxiety (HADS) score up to Q2: 11+, moderate, severe, Weeks since vaccination | Median: 8.0, IQR: (6.0, 9.0) | Median: 3.0, IQR: (2.0, 6.0) | Median: 25.0, IQR: (20.0, 28.0) | Median: 6.0, IQR: (4.0, 8.0) | | |
| Depression (HADS) score up to Q2: 8-10, mild, Anti-Spike level | n = 227, Median: 56.11, IQR: (25.54, 108.85), 5%: 5.13, 10%: 10.15 | n = 112, Median: 250.0, IQR: (250.0, 250.0), 5%: 119.76, 10%: 243.97 | n = 95, Median: 1301.0, IQR: (314.0, 5997.0), 5%: 72.43, 10%: 121.0 | n = 275, Median: 15186.0, IQR: (8866.5, 23795.0), 5%: 2872.7, 10%: 5317.4 | | |
| Depression (HADS) score up to Q2: 8-10, mild, Weeks since vaccination | Median: 8.0, IQR: (6.0, 9.0) | Median: 3.0, IQR: (2.0, 5.0) | Median: 25.0, IQR: (20.0, 29.0) | Median: 5.0, IQR: (3.5, 7.0) | | |
| Depression (HADS) score up to Q2: 11+, moderate, severe, Anti-Spike level | n = 151, Median: 58.17, IQR: (25.22, 125.3), 5%: 4.5, 10%: 10.02 | n = 60, Median: 250.0, IQR: (250.0, 250.0), 5%: 242.6, 10%: 250.0 | n = 85, Median: 1057.0, IQR: (311.0, 3963.0), 5%: 67.06, 10%: 90.94 | n = 146, Median: 14243.0, IQR: (7839.0, 24008.75), 5%: 3717.75, 10%: 4951.0 | | |
| Depression (HADS) score up to Q2: 11+, moderate, severe, Weeks since vaccination | Median: 8.0, IQR: (6.0, 9.0) | Median: 4.0, IQR: (2.0, 7.0) | Median: 25.0, IQR: (20.0, 29.0) | Median: 5.0, IQR: (3.0, 7.0) | | |

| | | | | |
|---|---|---|---|---|
| Anxiety (HADS) score up to Q4: 8-10, mild, Anti-Spike level | n = 309, Median: 52.3, IQR: (24.52, 120.6), 5%: 6.97, 10%: 10.6 | n = 160, Median: 250.0, IQR: (250.0, 250.0), 5%: 159.41, 10%: 250.0 | n = 144, Median: 747.0, IQR: (318.0, 3762.75), 5%: 91.02, 10%: 131.6 | n = 431, Median: 13480.0, IQR: (7929.0, 23890.0), 5%: 3646.0, 10%: 5281.0 |
| Anxiety (HADS) score up to Q4: 8-10, mild, Weeks since vaccination | Median: 8.0, IQR: (6.0, 9.0) | Median: 4.0, IQR: (2.75, 5.0) | Median: 25.0, IQR: (20.0, 27.0) | Median: 5.0, IQR: (4.0, 8.0) |
| Anxiety (HADS) score up to Q4: 11+, moderate, severe, Anti-Spike level | n = 297, Median: 57.38, IQR: (23.51, 120.6), 5%: 4.37, 10%: 10.04 | n = 138, Median: 250.0, IQR: (250.0, 250.0), 5%: 101.06, 10%: 244.7 | n = 173, Median: 1628.0, IQR: (366.0, 6905.0), 5%: 72.56, 10%: 106.4 | n = 327, Median: 15042.0, IQR: (8254.5, 23350.0), 5%: 3740.7, 10%: 5261.8 |
| Anxiety (HADS) score up to Q4: 11+, moderate, severe, Weeks since vaccination | Median: 8.0, IQR: (6.0, 9.0) | Median: 3.0, IQR: (2.0, 6.0) | Median: 24.0, IQR: (19.0, 28.0) | Median: 6.0, IQR: (4.0, 8.0) |
| Depression (HADS) score up to Q4: 8-10, mild, Anti-Spike level | n = 244, Median: 55.09, IQR: (22.38, 110.9), 5%: 3.4, 10%: 8.83 | n = 129, Median: 250.0, IQR: (250.0, 250.0), 5%: 118.82, 10%: 236.86 | n = 106, Median: 1332.5, IQR: (401.5, 5759.5), 5%: 99.3, 10%: 136.0 | n = 331, Median: 14658.0, IQR: (8692.0, 24706.5), 5%: 3202.5, 10%: 5281.0 |
| Depression (HADS) score up to Q4: 8-10, mild, Weeks since vaccination | Median: 8.0, IQR: (6.0, 9.0) | Median: 3.0, IQR: (2.0, 5.0) | Median: 25.0, IQR: (20.0, 28.0) | Median: 5.0, IQR: (4.0, 7.5) |
| Depression (HADS) score up to Q4: 11+, moderate, severe, Anti-Spike level | n = 175, Median: 57.51, IQR: (22.77, 122.05), 5%: 4.53, 10%: 10.01 | n = 67, Median: 250.0, IQR: (250.0, 250.0), 5%: 146.4, 10%: 250.0 | n = 105, Median: 1057.0, IQR: (292.0, 4208.0), 5%: 61.84, 10%: 90.94 | n = 174, Median: 14862.5, IQR: (8167.25, 24193.0), 5%: 3838.75, 10%: 5096.7 |
| Depression (HADS) score up to Q4: 11+, moderate, severe, Weeks since vaccination | Median: 8.0, IQR: (6.0, 9.0) | Median: 4.0, IQR: (2.0, 6.0) | Median: 25.0, IQR: (20.0, 29.0) | Median: 5.0, IQR: (3.0, 7.0) |

#### Post-vaccination infection descriptive statistics

Post-vaccination infection descriptive statistics are also provided in supplementary spreadsheet file for easier viewing.

Table S 6. Descriptive statistics relating to post-vaccination infections within TwinsUK, within groups of individuals with varying vaccination status at Q2 and Q4 testing.

| Vaccination status | All who are vaccinated once or more at Q4 | Single-vaccinated at Q2 | Double-vaccinated at Q2 |
|---|---|---|---|
| Q2 anti-Spike antibody level value (BAU/mL): Data not available, n (%) | 267/3539 (7.5%) | 0/1375 (0.0%) | 0/748 (0.0%) |
| Q2 anti-Spike antibody level value (BAU/mL): Median (IQR) | 90.42 (24.65, 250.0) | 53.3 (22.72, 121.2) | 250.0 (250.0, 250.0) |
| Post-vaccination infection between Q2 and Q4 testing: Data not available, n (%) | 570/3539 (16.1%) | 213/1375 (15.5%) | 124/748 (16.6%) |
| Post-vaccination infection between Q2 and Q4 testing: Yes, n (%) | 252/2969 (8.5%) | 105/1162 (9.0%) | 45/624 (7.2%) |
| Post-vaccination infection between Q2 and Q4 testing: No: Q2 Anti-Spike level | Median: 91.56 IQR:(25.46, 250.0), 5%: 0.4, 10%: 0.85 | Median: 57.17 IQR:(23.7, 129.4), 5%: 5.08, 10%: 10.0 | Median: 250.0 IQR:(250.0, 250.0), 5%: 136.47, 10%: 250.0 |
| Post-vaccination infection between Q2 and Q4 testing: Yes: Q2 Anti-Spike level | Median: 48.49 IQR:(8.92, 234.95), 5%: 0.4, 10%: 0.4 | Median: 40.04 IQR:(13.72, 81.82), 5%: 2.54, 10%: 5.11 | Median: 250.0 IQR:(250.0, 250.0), 5%: 149.3, 10%: 240.82 |
| Post-vaccination infection at any time: Data not available, n (%) | 546/3539 (15.4%) | 205/1375 (14.9%) | 114/748 (15.2%) |
| Post-vaccination infection at any time: Yes, n (%) | 276/2993 (9.2%) | 113/1170 (9.7%) | 55/634 (8.7%) |
| Post-vaccination infection, exact date of infection unknown: Yes, n | 32 | 9 | 6 |
| Post-vaccination infection while single-vaccinated: Yes, n | 34 | 12 | 8 |
| Post-vaccination infection while double-vaccinated: Yes, n | 171 | 83 | 29 |
| Post-vaccination infection while triple-vaccinated: Yes, n | 39 | 9 | 12 |
| Post-vaccination infection while single-vaccinated: Weeks since first vaccination: Median (IQR) | 5.5 (1.25, 9.0) | 4.0 (1.75, 9.5) | 1.0 (0.0, 4.0) |
| Post-vaccination infection while double-vaccinated: Weeks since second vaccination: Median (IQR) | 19.0 (11.5, 23.0) | 19.0 (11.0, 23.0) | 23.0 (20.0, 26.0) |
| Post-vaccination infection while triple-vaccinated: Weeks since third vaccination: Median (IQR) | 5.0 (1.0, 7.5) | 1.0 (1.0, 2.0) | 9.5 (7.75, 10.5) |
| Post-vaccination infection while single-vaccinated: Days since first vaccination: Median (IQR) | 41.5 (12.25, 64.75) | 29.5 (14.75, 68.0) | 7.5 (4.25, 28.5) |
| Post-vaccination infection while double-vaccinated: Days since second vaccination: Median (IQR) | 136.0 (83.5, 167.0) | 136.0 (78.5, 164.5) | 167.0 (143.0, 187.0) |
| Post-vaccination infection while triple-vaccinated: Days since third vaccination: Median (IQR) | 37.0 (9.0, 57.0) | 10.0 (9.0, 16.0) | 69.0 (57.75, 78.0) |
| Post-vaccination infection while single-vaccinated: Date & likely variant: 1. Before May 2021: Alpha, n | 22 | 8 | 8 |

| | | | |
|---|---|---|---|
| Post-vaccination infection while single-vaccinated: Date & likely variant: 2. May-Dec 2021: Delta, n | 9 | < 5 | < 5 |
| Post-vaccination infection while single-vaccinated: Date & likely variant: 3. After Dec 2021: Omicron, n | < 5 | < 5 | < 5 |
| Post-vaccination infection while double-vaccinated: Date & likely variant: 1. Before May 2021: Alpha, n | < 5 | < 5 | < 5 |
| Post-vaccination infection while double-vaccinated: Date & likely variant: 2. May-Dec 2021: Delta, n | 158 | 80 | 26 |
| Post-vaccination infection while double-vaccinated: Date & likely variant: 3. After Dec 2021: Omicron, n | 11 | < 5 | < 5 |
| Post-vaccination infection while triple-vaccinated: Date & likely variant: 1. Before May 2021: Alpha, n | < 5 | < 5 | < 5 |
| Post-vaccination infection while triple-vaccinated: Date & likely variant: 2. May-Dec 2021: Delta, n | 17 | 5 | < 5 |
| Post-vaccination infection while triple-vaccinated: Date & likely variant: 3. After Dec 2021: Omicron, n | 22 | < 5 | 9 |
| Post-vaccination infection while single-vaccinated: UK 7-day rolling case rate at time of infection: Median (IQR) | 104.45 (57.5, 387.5) | 57.05 (37.97, 65.22) | 236.05 (130.65, 525.28) |
| Post-vaccination infection while double-vaccinated: UK 7-day rolling case rate at time of infection: Median (IQR) | 373.1 (350.1, 442.85) | 392.6 (352.5, 443.45) | 362.5 (309.9, 406.4) |
| Post-vaccination infection while triple-vaccinated: UK 7-day rolling case rate at time of infection: Median (IQR) | 507.5 (444.35, 1214.65) | 448.7 (445.1, 530.5) | 1017.05 (472.18, 1697.02) |

#### Factors associated with post-vaccination infection

Table S 7. Logistic regression model results, testing for association between post-vaccination infection and anti-Spike antibody levels recorded in Q2 after first and second SARS-CoV-2 vaccination within TwinsUK. Results present odds ratios, unadjusted 95% confidence intervals, and p-values adjusted for multiple testing. Results based on fewer than 3 individuals having post-vaccination infection are suppressed.

| Control variables | Test variable | Sample: Single-vaccinated at Q2 | Sample: Double-vaccinated at Q2 |
|---|---|---|---|
| | | Outcome: Post-vaccination infection after Q2 | Outcome: Post-vaccination infection after Q2 |
| Adjusted for: Age, Weeks since vaccination | Q2 Antibody level: 1. 0.4-250 BAU/mL | | 1.65 (0.61, 4.43), p = 0.64 |
| Adjusted for: Age, Weeks since vaccination | Q2 Antibody level: 1. 0.4-50 BAU/mL | 2.64 (1.26, 5.54), p = 0.04 | Suppressed |
| Adjusted for: Age, Weeks since vaccination | Q2 Antibody level: 2. 50-150 BAU/mL | 2.47 (1.12, 5.45), p = 0.04 | Suppressed |
| Adjusted for: Age, Weeks since vaccination | Q2 Antibody level: 3. 150-250 BAU/mL | Suppressed | Suppressed |
| Adjusted for: Age, Weeks since vaccination | Q2 Antibody level: Quintile 1 (lowest 20%) | 2.99 (1.45, 6.17), p = 0.02 | |
| Adjusted for: Age, Weeks since vaccination | Q2 Antibody level: Quintile 2 | 2.14 (0.99, 4.65), p = 0.06 | |
| Adjusted for: Age, Weeks since vaccination | Q2 Antibody level: Quintile 3 | 2.4 (1.1, 5.2), p = 0.04 | |
| Adjusted for: Age, Weeks since vaccination | Q2 Antibody level: Quintile 4 | 2.49 (1.15, 5.43), p = 0.04 | |
| Adjusted for: Weeks since vaccination | Q2 Antibody level: 1. 0.4-50 BAU/mL | 2.45 (1.17, 5.13), p = 0.06 | |
| Adjusted for: Weeks since vaccination | Q2 Antibody level: 2. 50-150 BAU/mL | 2.3 (1.04, 5.07), p = 0.06 | |
| Adjusted for: Weeks since vaccination | Q2 Antibody level: 3. 150-250 BAU/mL | Suppressed | |
| Adjusted for: Weeks since vaccination | Q2 Antibody level: Quintile 1 (lowest 20%) | 2.85 (1.39, 5.86), p = 0.03 | |
| Adjusted for: Weeks since vaccination | Q2 Antibody level: Quintile 2 | 2.04 (0.94, 4.43), p = 0.08 | |
| Adjusted for: Weeks since vaccination | Q2 Antibody level: Quintile 3 | 2.26 (1.04, 4.92), p = 0.06 | |
| Adjusted for: Weeks since vaccination | Q2 Antibody level: Quintile 4 | 2.39 (1.1, 5.22), p = 0.06 | |
| No control variables | Q2 Antibody level: 1. 0.4-250 BAU/mL | | 1.29 (0.49, 3.43), p = 0.73 |
| No control variables | Q2 Antibody level: 1. 0.4-50 BAU/mL | 2.39 (1.16, 4.92), p = 0.06 | Suppressed |
| No control variables | Q2 Antibody level: 2. 50-150 BAU/mL | 1.91 (0.89, 4.09), p = 0.11 | Suppressed |
| No control variables | Q2 Antibody level: 3. 150-250 BAU/mL | Suppressed | Suppressed |
| No control variables | Q2 Antibody level: Quintile 1 (lowest 20%) | 3.23 (1.58, 6.58), p = 0.009 | |
| No control variables | Q2 Antibody level: Quintile 2 | 1.97 (0.92, 4.21), p = 0.11 | |
| No control variables | Q2 Antibody level: Quintile 3 | 1.95 (0.92, 4.15), p = 0.11 | |
| No control variables | Q2 Antibody level: Quintile 4 | 2.04 (0.96, 4.33), p = 0.11 | |

Results of logistic regression analysis testing association between post-vaccination infection and socio-demographic, SARS-CoV-2 vaccination and SARS-CoV-2 infection variables are also provided in supplementary spreadsheet file for easier viewing.

Table S 8. Logistic regression model results, testing for association between post-vaccination infection and socio-demographic, SARS-CoV-2 vaccination, and SARS-CoV-2 infection variables for TwinsUK individuals who participated in antibody testing at Q2 and one or both of Q4 antibody testing and Q4 questionnaire, who reported one or more vaccination reported by Q4. Results present odds ratios, unadjusted 95% confidence intervals, and p-values adjusted for multiple testing. Results based on fewer than 3 individuals having post-vaccination infection are suppressed. Variables with adjusted p-values < 0.05 are highlighted in bold.

| Control variables | Domain | Test variable | Outcome: post-vaccination infection at any time |
|---|---|---|---|
| Adjusted for: Age | COVID-19 infection | Anti-Nucleocapsid antibody status: Positive | 0.55 (0.34, 0.89), p = 0.17 |
| Adjusted for: Age | COVID-19 infection | SARS-CoV-2 infection status (self-reported): Confirmed case | 0.92 (0.64, 1.32), p = 0.86 |
| Adjusted for: Age | COVID-19 infection | SARS-CoV-2 infection status (self-reported): Suspected case | 0.73 (0.47, 1.13), p = 0.75 |
| Adjusted for: Age | COVID-19 infection | SARS-CoV-2 infection status (self-reported): Suspected or confirmed case | 0.84 (0.62, 1.13), p = 0.75 |
| Adjusted for: Age | COVID-19 infection | SARS-CoV-2 infection status (self-reported): Unsure | 1.26 (0.56, 2.84), p = 0.86 |
| Adjusted for: Age | COVID-19 infection | SARS-CoV-2 infection status (serology-based): Evidence of natural infection | **0.46 (0.32, 0.67), p = 0.0009** |
| Adjusted for: Age | COVID-19 vaccination | First vaccination received: Other | 0.83 (0.35, 1.97), p = 0.87 |
| Adjusted for: Age | COVID-19 vaccination | First vaccination received: AZD1222 (Oxford/AZ) | 1.36 (1.03, 1.78), p = 0.27 |
| Adjusted for: Age | Socio-demographics | Deprivation (IMD): Most deprived 40% (decile 1-4) | 0.91 (0.65, 1.27), p = 0.86 |
| Adjusted for: Age | Socio-demographics | Employment status: In education | 0.82 (0.35, 1.9), p = 0.86 |
| Adjusted for: Age | Socio-demographics | Employment status: Looking after home or family (unpaid care) | 0.8 (0.4, 1.59), p = 0.85 |
| Adjusted for: Age | Socio-demographics | Employment status: Maternity leave | Suppressed |
| Adjusted for: Age | Socio-demographics | Employment status: Other | Suppressed |
| Adjusted for: Age | Socio-demographics | Employment status: Permanently (or long-term) sick or disabled | Suppressed |
| Adjusted for: Age | Socio-demographics | Employment status: Retired | **0.5 (0.34, 0.75), p = 0.01** |
| Adjusted for: Age | Socio-demographics | Employment status: Self-employed | 0.88 (0.55, 1.4), p = 0.86 |
| Adjusted for: Age | Socio-demographics | Employment status: Semi-retired/part-time employment | 1.89 (0.53, 6.75), p = 0.81 |
| Adjusted for: Age | Socio-demographics | Employment status: Unemployed | Suppressed |

| | | | |
|---|---|---|---|
| Adjusted for: Age | Socio-demographics | Employment status: Unpaid/voluntary work | 0.36 (0.11, 1.17), p = 0.69 |
| Adjusted for: Age | Socio-demographics | Ethnicity: Other than white | 0.99 (0.47, 2.1), p = 0.99 |
| Adjusted for: Age | Socio-demographics | IMD: -1 decile (increasing deprivation) | 0.98 (0.93, 1.03), p = 0.81 |
| Adjusted for: Age | Socio-demographics | Highest educational attainment: NVQ level 3 or lower | 1.13 (0.85, 1.49), p = 0.81 |
| Adjusted for: Age | Socio-demographics | RUC: Urban | 1.01 (0.76, 1.35), p = 0.97 |
| Adjusted for: Age, SARS-CoV-2 infection status (serology-based) | COVID-19 infection | Anti-Nucleocapsid antibody status: Positive | 1.27 (0.63, 2.58), p = 0.83 |
| Adjusted for: Age, SARS-CoV-2 infection status (serology-based) | COVID-19 infection | SARS-CoV-2 infection status (self-reported): Confirmed case | 1.66 (1.07, 2.57), p = 0.32 |
| Adjusted for: Age, SARS-CoV-2 infection status (serology-based) | COVID-19 infection | SARS-CoV-2 infection status (self-reported): Suspected case | 0.78 (0.5, 1.21), p = 0.83 |
| Adjusted for: Age, SARS-CoV-2 infection status (serology-based) | COVID-19 infection | SARS-CoV-2 infection status (self-reported): Suspected or confirmed case | 1.11 (0.81, 1.53), p = 0.84 |
| Adjusted for: Age, SARS-CoV-2 infection status (serology-based) | COVID-19 infection | SARS-CoV-2 infection status (self-reported): Unsure | 1.28 (0.56, 2.9), p = 0.84 |
| Adjusted for: Age, SARS-CoV-2 infection status (serology-based) | COVID-19 vaccination | First vaccination received: Other | 0.85 (0.35, 2.08), p = 0.92 |
| Adjusted for: Age, SARS-CoV-2 infection status (serology-based) | COVID-19 vaccination | First vaccination received: AZD1222 (Oxford/AZ) | 1.29 (0.98, 1.69), p = 0.75 |
| Adjusted for: Age, SARS-CoV-2 infection status (serology-based) | Socio-demographics | Deprivation (IMD): Most deprived 40% (decile 1-4) | 0.92 (0.65, 1.29), p = 0.84 |
| Adjusted for: Age, SARS-CoV-2 infection status (serology-based) | Socio-demographics | Employment status: In education | 0.76 (0.32, 1.85), p = 0.84 |
| Adjusted for: Age, SARS-CoV-2 infection status (serology-based) | Socio-demographics | Employment status: Looking after home or family (unpaid care) | 0.82 (0.41, 1.62), p = 0.84 |
| Adjusted for: Age, SARS-CoV-2 infection status (serology-based) | Socio-demographics | Employment status: Maternity leave | Suppressed |
| Adjusted for: Age, SARS-CoV-2 infection status (serology-based) | Socio-demographics | Employment status: Other | Suppressed |
| Adjusted for: Age, SARS-CoV-2 infection status (serology-based) | Socio-demographics | Employment status: Permanently (or long-term) sick or disabled | Suppressed |
| Adjusted for: Age, SARS-CoV-2 infection status (serology-based) | Socio-demographics | Employment status: Retired | **0.49 (0.33, 0.74), p = 0.01** |
| Adjusted for: Age, SARS-CoV-2 infection status (serology-based) | Socio-demographics | Employment status: Self-employed | 0.83 (0.52, 1.32), p = 0.83 |
| Adjusted for: Age, SARS-CoV-2 infection status (serology-based) | Socio-demographics | Employment status: Semi-retired/part-time employment | 1.84 (0.52, 6.54), p = 0.83 |
| Adjusted for: Age, SARS-CoV-2 infection status (serology-based) | Socio-demographics | Employment status: Unemployed | Suppressed |
| Adjusted for: Age, SARS-CoV-2 infection status (serology-based) | Socio-demographics | Employment status: Unpaid/voluntary work | 0.36 (0.11, 1.18), p = 0.75 |

| | | | |
|---|---|---|---|
| Adjusted for: Age, SARS-CoV-2 infection status (serology-based) | Socio-demographics | Ethnicity: Other than white | 1.09 (0.51, 2.35), p = 0.93 |
| Adjusted for: Age, SARS-CoV-2 infection status (serology-based) | Socio-demographics | IMD: -1 decile (increasing deprivation) | 0.98 (0.93, 1.03), p = 0.83 |
| Adjusted for: Age, SARS-CoV-2 infection status (serology-based) | Socio-demographics | Highest educational attainment: NVQ level 3 or lower | 1.15 (0.87, 1.52), p = 0.83 |
| Adjusted for: Age, SARS-CoV-2 infection status (serology-based) | Socio-demographics | RUC: Urban | 1.03 (0.77, 1.38), p = 0.93 |
| Adjusted for: Age, Sex, SARS-CoV-2 infection status (serology-based) | COVID-19 infection | Anti-Nucleocapsid antibody status: Positive | 1.28 (0.63, 2.59), p = 0.86 |
| Adjusted for: Age, Sex, SARS-CoV-2 infection status (serology-based) | COVID-19 infection | SARS-CoV-2 infection status (self-reported): Confirmed case | 1.67 (1.08, 2.6), p = 0.26 |
| Adjusted for: Age, Sex, SARS-CoV-2 infection status (serology-based) | COVID-19 infection | SARS-CoV-2 infection status (self-reported): Suspected case | 0.79 (0.51, 1.23), p = 0.84 |
| Adjusted for: Age, Sex, SARS-CoV-2 infection status (serology-based) | COVID-19 infection | SARS-CoV-2 infection status (self-reported): Suspected or confirmed case | 1.13 (0.82, 1.55), p = 0.86 |
| Adjusted for: Age, Sex, SARS-CoV-2 infection status (serology-based) | COVID-19 infection | SARS-CoV-2 infection status (self-reported): Unsure | 1.28 (0.56, 2.93), p = 0.86 |
| Adjusted for: Age, Sex, SARS-CoV-2 infection status (serology-based) | COVID-19 vaccination | First vaccination received: Other | 0.8 (0.33, 1.95), p = 0.87 |
| Adjusted for: Age, Sex, SARS-CoV-2 infection status (serology-based) | COVID-19 vaccination | First vaccination received: AZD1222 (Oxford/AZ) | 1.28 (0.98, 1.68), p = 0.63 |
| Adjusted for: Age, Sex, SARS-CoV-2 infection status (serology-based) | Socio-demographics | Deprivation (IMD): Most deprived 40% (decile 1-4) | 0.92 (0.65, 1.29), p = 0.87 |
| Adjusted for: Age, Sex, SARS-CoV-2 infection status (serology-based) | Socio-demographics | Employment status: In education | 0.76 (0.31, 1.85), p = 0.86 |
| Adjusted for: Age, Sex, SARS-CoV-2 infection status (serology-based) | Socio-demographics | Employment status: Looking after home or family (unpaid care) | 0.86 (0.43, 1.71), p = 0.87 |
| Adjusted for: Age, Sex, SARS-CoV-2 infection status (serology-based) | Socio-demographics | Employment status: Maternity leave | Suppressed |
| Adjusted for: Age, Sex, SARS-CoV-2 infection status (serology-based) | Socio-demographics | Employment status: Other | Suppressed |
| Adjusted for: Age, Sex, SARS-CoV-2 infection status (serology-based) | Socio-demographics | Employment status: Permanently (or long-term) sick or disabled | Suppressed |
| Adjusted for: Age, Sex, SARS-CoV-2 infection status (serology-based) | Socio-demographics | Employment status: Retired | **0.5 (0.33, 0.74), p = 0.01** |
| Adjusted for: Age, Sex, SARS-CoV-2 infection status (serology-based) | Socio-demographics | Employment status: Self-employed | 0.82 (0.51, 1.31), p = 0.85 |
| Adjusted for: Age, Sex, SARS-CoV-2 infection status (serology-based) | Socio-demographics | Employment status: Semi-retired/part-time employment | 1.73 (0.51, 5.93), p = 0.85 |
| Adjusted for: Age, Sex, SARS-CoV-2 infection status (serology-based) | Socio-demographics | Employment status: Unemployed | Suppressed |
| Adjusted for: Age, Sex, SARS-CoV-2 infection status (serology-based) | Socio-demographics | Employment status: Unpaid/voluntary work | 0.36 (0.11, 1.19), p = 0.68 |

| | | | |
|---|---|---|---|
| Adjusted for: Age, Sex, SARS-CoV-2 infection status (serology-based) | Socio-demographics | Ethnicity: Other than white | 1.11 (0.51, 2.39), p = 0.93 |
| Adjusted for: Age, Sex, SARS-CoV-2 infection status (serology-based) | Socio-demographics | IMD: -1 decile (increasing deprivation) | 0.98 (0.93, 1.03), p = 0.86 |
| Adjusted for: Age, Sex, SARS-CoV-2 infection status (serology-based) | Socio-demographics | Highest educational attainment: NVQ level 3 or lower | 1.19 (0.89, 1.57), p = 0.84 |
| Adjusted for: Age, Sex, SARS-CoV-2 infection status (serology-based) | Socio-demographics | RUC: Urban | 1.02 (0.76, 1.37), p = 0.99 |
| Adjusted for: Age, Sex, SARS-CoV-2 infection status (serology-based) | Socio-demographics | Sex: Male | 1.49 (1.06, 2.12), p = 0.26 |
| No control variables | COVID-19 infection | Anti-Nucleocapsid antibody status: Positive | 0.71 (0.45, 1.12), p = 0.42 |
| No control variables | COVID-19 infection | SARS-CoV-2 infection status (self-reported): Confirmed case | 1.05 (0.74, 1.48), p = 0.89 |
| No control variables | COVID-19 infection | SARS-CoV-2 infection status (self-reported): Suspected case | 0.81 (0.53, 1.25), p = 0.67 |
| No control variables | COVID-19 infection | SARS-CoV-2 infection status (self-reported): Suspected or confirmed case | 0.95 (0.71, 1.26), p = 0.86 |
| No control variables | COVID-19 infection | SARS-CoV-2 infection status (self-reported): Unsure | 1.46 (0.65, 3.27), p = 0.68 |
| No control variables | COVID-19 infection | SARS-CoV-2 infection status (serology-based): Evidence of natural infection | **0.53 (0.37, 0.77), p = 0.006** |
| No control variables | COVID-19 vaccination | First vaccination received: Other | 1.4 (0.58, 3.35), p = 0.75 |
| No control variables | COVID-19 vaccination | First vaccination received: AZD1222 (Oxford/AZ) | 1.18 (0.91, 1.53), p = 0.52 |
| No control variables | Socio-demographics | Age: 50-59 | 0.94 (0.68, 1.31), p = 0.86 |
| No control variables | Socio-demographics | Age: 60-69 | **0.49 (0.35, 0.69), p = 0.0007** |
| No control variables | Socio-demographics | Age: 70-79 | **0.31 (0.21, 0.45), p < 0.0001** |
| No control variables | Socio-demographics | Age: 80+ | **0.18 (0.07, 0.44), p = 0.002** |
| No control variables | Socio-demographics | Deprivation (IMD): Most deprived 40% (decile 1-4) | 1.0 (0.72, 1.39), p = 0.99 |
| No control variables | Socio-demographics | Employment status: In education | 1.16 (0.51, 2.63), p = 0.86 |
| No control variables | Socio-demographics | Employment status: Looking after home or family (unpaid care) | 0.69 (0.35, 1.35), p = 0.59 |
| No control variables | Socio-demographics | Employment status: Maternity leave | Suppressed |
| No control variables | Socio-demographics | Employment status: Other | Suppressed |
| No control variables | Socio-demographics | Employment status: Permanently (or long-term) sick or disabled | Suppressed |
| No control variables | Socio-demographics | Employment status: Retired | **0.36 (0.27, 0.49), p < 0.0001** |
| No control variables | Socio-demographics | Employment status: Self-employed | 0.8 (0.51, 1.26), p = 0.66 |

| | | | |
|---|---|---|---|
| No control variables | Socio-demographics | Employment status: Semi-retired/part-time employment | 1.41 (0.4, 4.98), p = 0.83 |
| No control variables | Socio-demographics | Employment status: Unemployed | Suppressed |
| No control variables | Socio-demographics | Employment status: Unpaid/voluntary work | 0.28 (0.09, 0.9), p = 0.12 |
| No control variables | Socio-demographics | Ethnicity: Other than white | 1.35 (0.64, 2.85), p = 0.75 |
| No control variables | Socio-demographics | IMD: -1 decile (increasing deprivation) | 0.99 (0.94, 1.05), p = 0.9 |
| No control variables | Socio-demographics | Highest educational attainment: NVQ level 3 or lower | 0.85 (0.65, 1.11), p = 0.55 |
| No control variables | Socio-demographics | RUC: Urban | 1.11 (0.83, 1.47), p = 0.76 |

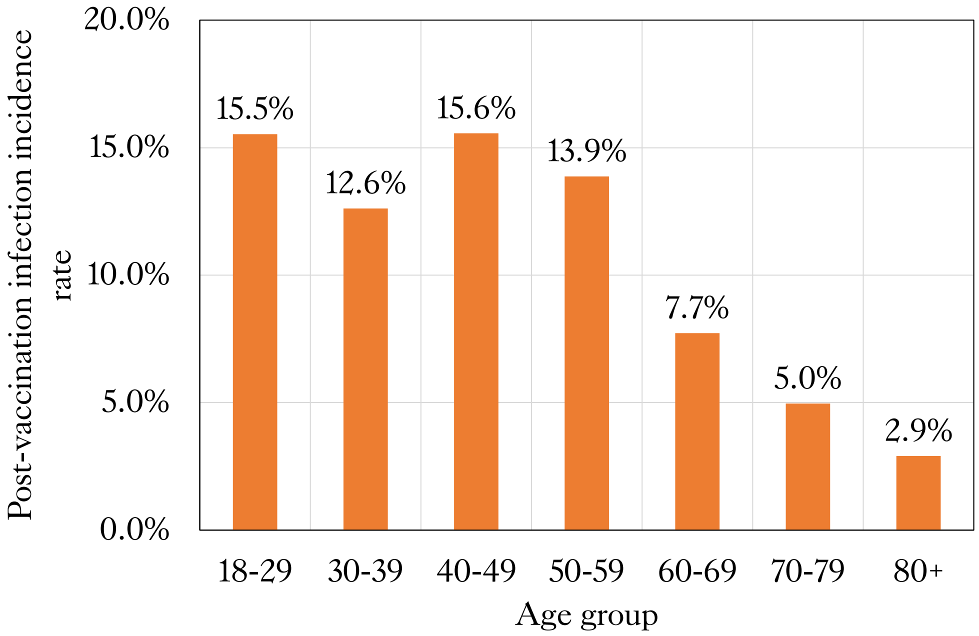

Figure S 6. Incidence of post-vaccination infection for TwinsUK individuals, split by age group.

### Factors associated with low antibody levels: logistic regression results

Results of logistic regression analysis testing association with low anti-Spike antibody levels are also provided in supplementary spreadsheet file for easier viewing.

Table S 9. Logistic regression model results, testing for association with low anti-Spike antibody levels after first, second and third SARS-CoV-2 vaccination within TwinsUK and ALSPAC at Q2 or Q4 testing. Results present odds ratios, unadjusted 95% confidence intervals, and p-values adjusted for multiple testing. Results based on fewer than 3 individuals being in the low antibody level group are suppressed. Sets of adjustment variables included in addition to the exposure variable in each model were age, sex, most recent vaccine received and weeks since most recent vaccination, aside from cases where the effect of adjustment variables were themselves tested. In these cases, all other adjustment variables within the given set were included in addition to the adjustment variable being tested. Variables with adjusted p-values < 0.05 are highlighted in bold.

| | **Cohort** | **TwinsUK** | | | | | | | | **ALSPAC** | | |
|---|---|---|---|---|---|---|---|---|---|---|---|---|
| | **Testing period** | **Q2** | | | | **Q4** | | | | **Q2** | | |
| | **Number of vaccinations received** | **1** | | **2** | | **2** | | **3** | | **1** | | **2** |
| | **Outcome threshold** | **Lowest 10%** | **Lowest 5%** | **Lowest 5%** | **Lowest 8%** | **Lowest 10%** | **Lowest 5%** | **Lowest 10%** | **Lowest 5%** | **Lowest 10%** | **Lowest 5%** | **Lowest 8%** |
| **Variable group** | **Variable** | | | | | | | | | | | |
| COVID-19 at-risk | Advised on "Shielded Patient List": Yes | **4.03 (2.2, 7.42), p = 0.0001** | **4.39 (2.14, 9.0), p = 0.0005** | **3.56 (1.67, 7.59), p = 0.009** | **3.0 (1.55, 5.8), p = 0.009** | 0.96 (0.21, 4.36), p = 0.96 | Suppressed | **2.42 (1.62, 3.64), p = 0.0002** | **3.16 (1.92, 5.21), p < 0.0001** | **4.13 (1.79, 9.51), p = 0.02** | **8.68 (3.39, 22.23), p = 0.0003** | 2.23 (0.58, 8.65), p = 0.78 |
| COVID-19 at-risk | Frailty (PRISMA-7 assessment): Above threshold | | | | | | | | | 1.78 (0.71, 4.43), p = 0.55 | 1.63 (0.48, 5.55), p = 0.62 | 1.95 (0.34, 11.37), p = 0.78 |
| COVID-19 at-risk | Frailty Index: 2. Pre-frail | 1.22 (0.83, 1.78), p = 0.62 | 1.19 (0.7, 2.04), p = 0.77 | 0.72 (0.33, 1.61), p = 0.6 | 0.83 (0.44, 1.59), p = 0.7 | 0.69 (0.37, 1.32), p = 0.54 | 0.68 (0.29, 1.61), p = 0.61 | 0.92 (0.64, 1.32), p = 0.87 | 1.04 (0.64, 1.7), p = 0.92 | | | |
| COVID-19 at-risk | Frailty Index: 3. Frail | 1.5 (0.77, 2.93), p = 0.55 | 1.96 (0.85, 4.51), p = 0.35 | 0.65 (0.2, 2.13), p = 0.63 | 0.62 (0.24, 1.63), p = 0.55 | 0.57 (0.18, 1.79), p = 0.59 | Suppressed | 1.13 (0.66, 1.93), p = 0.87 | 1.09 (0.51, 2.31), p = 0.91 | | | |
| COVID-19 at-risk | Frailty Index: 4. Very frail | 1.39 (0.39, 4.94), p = 0.75 | Suppresse d | **5.55 (1.66, 18.57), p = 0.03** | **6.64 (2.32, 18.99), p = 0.004** | Suppressed | Suppressed | 1.81 (0.83, 3.94), p = 0.4 | 1.72 (0.61, 4.86), p = 0.59 | | | |
| COVID-19 at-risk | Prescribed immunosuppressant medication | 1.11 (0.55, 2.24), p = 0.87 | 1.55 (0.68, 3.52), p = 0.64 | **4.2 (1.86, 9.5), p = 0.006** | **3.65 (1.77, 7.51), p = 0.004** | 0.78 (0.2, 2.99), p = 0.85 | Suppressed | 1.46 (0.91, 2.37), p = 0.38 | 2.1 (1.18, 3.75), p = 0.06 | | | |

| | | | | | | | | | | | | |
|---|---|---|---|---|---|---|---|---|---|---|---|---|
| COVID-19 at-risk | Self-reported immunosuppressed | | | | | | | | | **6.2 (2.65, 14.5), p = 0.001** | **6.83 (2.59, 18.02), p = 0.002** | 1.82 (0.36, 9.17), p = 0.78 |
| COVID-19 infection | Anti-Nucleocapsid antibody status: Positive | Suppressed | Suppressed | Suppressed | Suppressed | Suppressed | Suppressed | **0.1 (0.04, 0.29), p = 0.0002** | Suppressed | Suppressed | Suppressed | Suppressed |
| COVID-19 infection | SARS-CoV-2 infection status (self-reported): Confirmed case | **0.28 (0.13, 0.59), p = 0.007** | 0.35 (0.14, 0.9), p = 0.15 | Suppressed | Suppressed | Suppressed | Suppressed | **0.25 (0.13, 0.45), p = 0.0001** | **0.32 (0.14, 0.71), p = 0.03** | Suppressed | Suppressed | Suppressed |
| COVID-19 infection | SARS-CoV-2 infection status (self-reported): Suspected case | 1.16 (0.71, 1.9), p = 0.72 | 0.7 (0.33, 1.5), p = 0.74 | 0.89 (0.26, 3.02), p = 0.92 | 0.77 (0.26, 2.29), p = 0.74 | 1.18 (0.57, 2.41), p = 0.82 | 1.06 (0.41, 2.72), p = 0.95 | 0.82 (0.5, 1.35), p = 0.79 | 0.78 (0.4, 1.53), p = 0.73 | 0.81 (0.5, 1.32), p = 0.78 | 1.02 (0.55, 1.91), p = 0.97 | 0.61 (0.16, 2.43), p = 0.78 |
| COVID-19 infection | SARS-CoV-2 infection status (self-reported): Suspected or confirmed case | 0.65 (0.42, 0.99), p = 0.23 | 0.51 (0.27, 0.95), p = 0.15 | 0.3 (0.09, 0.98), p = 0.13 | 0.35 (0.14, 0.86), p = 0.09 | **0.3 (0.16, 0.57), p = 0.001** | **0.26 (0.1, 0.65), p = 0.01** | **0.44 (0.3, 0.67), p = 0.0008** | 0.49 (0.28, 0.84), p = 0.05 | 0.69 (0.42, 1.12), p = 0.54 | 0.87 (0.47, 1.63), p = 0.84 | 0.56 (0.14, 2.18), p = 0.78 |
| COVID-19 infection | SARS-CoV-2 infection status (self-reported): Unsure | 0.6 (0.19, 1.93), p = 0.63 | Suppressed | Suppressed | Suppressed | Suppressed | Suppressed | 0.68 (0.28, 1.65), p = 0.77 | 0.86 (0.27, 2.79), p = 0.91 | | | |
| COVID-19 infection | SARS-CoV-2 infection status (serology-based): Evidence of natural infection | **0.49 (0.29, 0.82), p = 0.04** | 0.65 (0.34, 1.27), p = 0.51 | 0.37 (0.11, 1.25), p = 0.26 | 0.58 (0.26, 1.32), p = 0.36 | **0.09 (0.03, 0.26), p < 0.0001** | Suppressed | **0.45 (0.28, 0.71), p = 0.004** | **0.42 (0.22, 0.81), p = 0.05** | Suppressed | Suppressed | Suppressed |
| COVID-19 vaccination | First vaccination received: Other | Suppressed | Suppressed | | | | | | | | | |
| COVID-19 vaccination | First vaccination received: AZD1222 | **3.05 (1.46, 6.39), p = 0.02** | 2.5 (0.99, 6.31), p = 0.21 | | | | | | | 3.23 (1.36, 7.68), p = 0.09 | 3.33 (0.99, 11.2), p = 0.25 | |
| COVID-19 vaccination | Second vaccination received: Other | | | Suppressed | Suppressed | Suppressed | Suppressed | Suppressed | Suppressed | | | |
| COVID-19 vaccination | Second vaccination received: AZD1222 | | | **2.96 (1.4, 6.25), p = 0.03** | **4.65 (2.44, 8.86), p < 0.0001** | **45.69 (5.61, 372.28), p = 0.001** | **20.55 (2.43, 173.46), p = 0.01** | 1.13 (0.82, 1.56), p = 0.8 | 1.18 (0.77, 1.83), p = 0.73 | | | **20.34 (6.39, 64.68), p < 0.0001** |
| COVID-19 vaccination | Third vaccination received: mRNA-1273 | | | | | | | 0.34 (0.13, 0.86), p = 0.11 | Suppressed | | | |
| COVID-19 | Third vaccination received: Other | | | | | | | Suppressed | Suppressed | | | |

| | | | | | | | | | | | | |
|---|---|---|---|---|---|---|---|---|---|---|---|---|
| vaccination | | | | | | | | | | | | |
| COVID-19 vaccination | Weeks since first vaccination: +1 week | **0.83 (0.75, 0.92), p = 0.007** | 0.87 (0.75, 1.0), p = 0.21 | | | | | | | 0.83 (0.73, 0.95), p = 0.09 | 0.88 (0.73, 1.05), p = 0.36 | |
| COVID-19 vaccination | Weeks since second vaccination: +1 week | | | 1.09 (0.98, 1.21), p = 0.25 | **1.14 (1.04, 1.24), p = 0.02** | 1.02 (0.96, 1.09), p = 0.7 | 1.01 (0.92, 1.11), p = 0.93 | | | | | 1.13 (0.97, 1.32), p = 0.78 |
| COVID-19 vaccination | Weeks since third vaccination: +1 week | | | | | | | **1.24 (1.18, 1.29), p < 0.0001** | **1.22 (1.16, 1.29), p < 0.0001** | | | |
| Comorbidities | # Selected comorbidities: 1 | 1.23 (0.8, 1.88), p = 0.63 | 1.0 (0.54, 1.85), p = 0.99 | 1.0 (0.45, 2.22), p = 1.0 | 1.19 (0.64, 2.21), p = 0.7 | 0.72 (0.35, 1.49), p = 0.62 | 0.57 (0.2, 1.68), p = 0.54 | 0.99 (0.68, 1.43), p = 0.95 | 1.11 (0.68, 1.8), p = 0.86 | 0.92 (0.56, 1.51), p = 0.87 | 1.35 (0.72, 2.55), p = 0.54 | 0.45 (0.08, 2.44), p = 0.78 |
| Comorbidities | # Selected comorbidities: 2 | 0.89 (0.37, 2.14), p = 0.89 | 1.41 (0.53, 3.77), p = 0.76 | 0.67 (0.18, 2.53), p = 0.7 | 0.88 (0.34, 2.26), p = 0.86 | Suppressed | Suppressed | 0.92 (0.51, 1.66), p = 0.92 | 1.5 (0.76, 2.97), p = 0.55 | 1.43 (0.48, 4.28), p = 0.79 | 2.32 (0.65, 8.2), p = 0.39 | 3.18 (0.5, 20.04), p = 0.78 |
| Comorbidities | # Selected comorbidities: 3 | 4.27 (1.13, 16.07), p = 0.17 | 5.62 (1.32, 23.92), p = 0.11 | **7.68 (2.17, 27.14), p = 0.01** | **5.21 (1.5, 18.02), p = 0.04** | Suppressed | Suppressed | 2.78 (1.09, 7.12), p = 0.14 | 2.12 (0.6, 7.53), p = 0.55 | Suppressed | Suppressed | Suppressed |
| Comorbidities | # Selected comorbidities: 4+ | Suppressed | Suppressed | | | | | Suppressed | Suppressed | Suppressed | Suppressed | |
| Comorbidities | Comorbidity domain: Arthritis (any) | 1.56 (1.0, 2.45), p = 0.24 | 1.25 (0.67, 2.33), p = 0.76 | 1.22 (0.58, 2.58), p = 0.73 | 1.16 (0.62, 2.16), p = 0.74 | 0.72 (0.28, 1.85), p = 0.7 | Suppressed | 1.36 (0.96, 1.94), p = 0.32 | 1.43 (0.89, 2.29), p = 0.46 | | | |
| Comorbidities | Comorbidity domain: Cardiac Disease | 1.62 (0.65, 4.07), p = 0.62 | 1.46 (0.44, 4.92), p = 0.77 | 1.69 (0.59, 4.83), p = 0.5 | 1.51 (0.61, 3.72), p = 0.6 | Suppressed | Suppressed | 0.74 (0.37, 1.47), p = 0.77 | 0.56 (0.2, 1.57), p = 0.55 | | | |
| Comorbidities | Comorbidity domain: Cardiac Risk Factors | 0.82 (0.53, 1.27), p = 0.63 | 0.88 (0.5, 1.57), p = 0.89 | 1.14 (0.56, 2.33), p = 0.83 | 1.5 (0.85, 2.66), p = 0.34 | 0.55 (0.23, 1.29), p = 0.44 | 0.68 (0.22, 2.14), p = 0.69 | 1.06 (0.76, 1.49), p = 0.91 | 1.08 (0.69, 1.69), p = 0.88 | | | |
| Comorbidities | Comorbidity domain: Neurological Disease | 1.24 (0.77, 1.99), p = 0.63 | 1.25 (0.66, 2.36), p = 0.76 | 1.67 (0.75, 3.73), p = 0.38 | 1.35 (0.67, 2.72), p = 0.61 | 0.56 (0.24, 1.28), p = 0.44 | Suppressed | 0.96 (0.62, 1.48), p = 0.92 | 0.92 (0.52, 1.65), p = 0.91 | Suppressed | Suppressed | Suppressed |
| Comorbidities | Comorbidity domain: Subjective Memory Impairment | 0.64 (0.3, 1.34), p = 0.55 | 0.46 (0.14, 1.5), p = 0.51 | 1.24 (0.35, 4.41), p = 0.85 | 1.04 (0.34, 3.19), p = 0.95 | 1.02 (0.37, 2.84), p = 0.96 | Suppressed | 1.29 (0.72, 2.32), p = 0.77 | 1.57 (0.75, 3.29), p = 0.55 | | | |

| | | | | | | | | | | | | |
|---|---|---|---|---|---|---|---|---|---|---|---|---|
| Comorbidities | Comorbidity: Anxiety or Depression | 1.05 (0.65, 1.7), p = 0.93 | 1.02 (0.53, 1.96), p = 0.99 | 1.65 (0.74, 3.68), p = 0.39 | 1.46 (0.74, 2.89), p = 0.48 | 0.54 (0.23, 1.24), p = 0.43 | Suppressed | 1.06 (0.69, 1.62), p = 0.92 | 1.16 (0.67, 2.02), p = 0.81 | 0.81 (0.42, 1.58), p = 0.79 | 0.86 (0.36, 2.09), p = 0.88 | Suppressed |
| Comorbidities | Comorbidity: Anxiety or Stress Disorder | 1.13 (0.65, 1.97), p = 0.8 | 1.16 (0.56, 2.42), p = 0.9 | 0.94 (0.32, 2.79), p = 0.94 | 0.7 (0.27, 1.86), p = 0.65 | 0.61 (0.23, 1.61), p = 0.57 | Suppressed | 0.72 (0.41, 1.25), p = 0.62 | 0.72 (0.34, 1.52), p = 0.69 | 0.59 (0.26, 1.33), p = 0.55 | 0.88 (0.34, 2.3), p = 0.88 | Suppressed |
| Comorbidities | Comorbidity: Asthma | 0.98 (0.57, 1.69), p = 0.95 | 1.12 (0.56, 2.24), p = 0.9 | 1.55 (0.64, 3.75), p = 0.5 | 2.09 (1.05, 4.14), p = 0.11 | 0.7 (0.31, 1.6), p = 0.63 | Suppressed | 1.26 (0.83, 1.93), p = 0.64 | 0.97 (0.54, 1.74), p = 0.94 | 1.29 (0.63, 2.64), p = 0.79 | 0.88 (0.3, 2.59), p = 0.88 | Suppressed |
| Comorbidities | Comorbidity: Atrial Fibrillation | Suppressed | Suppressed | Suppressed | 0.96 (0.27, 3.41), p = 0.95 | Suppressed | Suppressed | 0.43 (0.16, 1.17), p = 0.34 | Suppressed | | | |
| Comorbidities | Comorbidity: Cancer (any) | 1.34 (0.69, 2.63), p = 0.63 | 1.1 (0.43, 2.83), p = 0.91 | 2.44 (1.07, 5.57), p = 0.11 | 2.13 (1.05, 4.31), p = 0.11 | 0.74 (0.2, 2.71), p = 0.82 | Suppressed | 1.37 (0.85, 2.2), p = 0.52 | 1.22 (0.64, 2.31), p = 0.76 | Suppressed | Suppressed | Suppressed |
| Comorbidities | Comorbidity: Lung disease | 1.42 (0.43, 4.68), p = 0.72 | 2.85 (0.85, 9.52), p = 0.32 | 2.45 (0.57, 10.47), p = 0.39 | 2.87 (0.88, 9.34), p = 0.2 | Suppressed | Suppressed | 1.16 (0.51, 2.66), p = 0.91 | Suppressed | Suppressed | Suppressed | Suppressed |
| Comorbidities | Comorbidity: Depression | 0.98 (0.53, 1.8), p = 0.95 | 0.71 (0.28, 1.8), p = 0.76 | 1.97 (0.75, 5.18), p = 0.32 | 1.86 (0.82, 4.2), p = 0.3 | 0.41 (0.12, 1.4), p = 0.44 | Suppressed | 1.2 (0.71, 2.03), p = 0.84 | 1.54 (0.8, 2.95), p = 0.54 | 1.23 (0.53, 2.86), p = 0.85 | 1.33 (0.45, 3.92), p = 0.78 | Suppressed |
| Comorbidities | Comorbidity: Diabetes (any) | 1.49 (0.42, 5.31), p = 0.72 | 2.93 (0.82, 10.47), p = 0.33 | 2.11 (0.62, 7.16), p = 0.39 | 1.26 (0.37, 4.33), p = 0.79 | Suppressed | Suppressed | 0.52 (0.15, 1.83), p = 0.69 | Suppressed | 2.1 (0.68, 6.49), p = 0.55 | 2.8 (0.77, 10.17), p = 0.35 | Suppressed |
| Comorbidities | Comorbidity: Heart disease (CHD or Heart failure) | Suppressed | Suppressed | Suppressed | Suppressed | | | 1.16 (0.35, 3.79), p = 0.92 | 1.82 (0.51, 6.56), p = 0.66 | Suppressed | Suppressed | 2.75 (0.43, 17.7), p = 0.78 |
| Comorbidities | Comorbidity: High Cholesterol | 0.81 (0.48, 1.35), p = 0.65 | 1.02 (0.54, 1.94), p = 0.99 | 1.63 (0.79, 3.37), p = 0.35 | 1.56 (0.85, 2.87), p = 0.33 | Suppressed | Suppressed | 0.92 (0.63, 1.33), p = 0.87 | 0.74 (0.45, 1.24), p = 0.55 | | | |
| Comorbidities | Comorbidity: Hypertension | 1.4 (0.86, 2.29), p = 0.49 | 2.04 (1.14, 3.66), p = 0.11 | 0.71 (0.31, 1.63), p = 0.6 | 0.81 (0.42, 1.58), p = 0.68 | 0.87 (0.28, 2.68), p = 0.89 | Suppressed | 1.04 (0.71, 1.53), p = 0.92 | 1.2 (0.73, 1.99), p = 0.73 | 1.25 (0.72, 2.18), p = 0.78 | 1.76 (0.89, 3.48), p = 0.35 | 1.32 (0.33, 5.34), p = 0.88 |
| Comorbidities | Comorbidity: Osteoporosis | 1.06 (0.53, 2.1), p = 0.94 | 1.3 (0.57, 2.95), p = 0.77 | 1.38 (0.53, 3.59), p = 0.66 | 1.66 (0.79, 3.5), p = 0.35 | Suppressed | Suppressed | 1.18 (0.75, 1.85), p = 0.8 | 1.16 (0.64, 2.12), p = 0.82 | | | |
| Comorbidities | Comorbidity: Rheumatoid Arthritis | 1.61 (0.63, 4.12), p = 0.62 | Suppressed | 3.85 (0.94, 15.74), p = 0.16 | 2.39 (0.57, 10.02), p = 0.42 | Suppressed | Suppressed | **3.03 (1.35, 6.82), p = 0.04** | **4.04 (1.62, 10.04), p = 0.02** | | | |

| | | | | | | | | | | | | |
|---|---|---|---|---|---|---|---|---|---|---|---|---|
| Comorbidities | Comorbidity: Stroke | 3.2 (0.95, 10.72), p = 0.26 | Suppressed | Suppressed | Suppressed | Suppressed | Suppressed | 0.88 (0.26, 3.06), p = 0.92 | Suppressed | | | |
| General health | BMI: +1 kg/m^2 | 1.03 (0.99, 1.07), p = 0.4 | 1.05 (1.0, 1.1), p = 0.19 | 1.05 (0.98, 1.13), p = 0.31 | 1.03 (0.96, 1.09), p = 0.61 | 1.03 (0.98, 1.08), p = 0.44 | 1.03 (0.97, 1.1), p = 0.56 | 0.97 (0.93, 1.02), p = 0.55 | 0.98 (0.93, 1.04), p = 0.76 | 0.96 (0.91, 1.0), p = 0.33 | 0.92 (0.86, 0.98), p = 0.13 | 0.97 (0.86, 1.09), p = 0.85 |
| General health | Self-rated health: -1 rating (5-point scale, decreasing health) | **1.35 (1.11, 1.64), p = 0.02** | 1.45 (1.09, 1.92), p = 0.08 | **1.65 (1.14, 2.41), p = 0.04** | **1.55 (1.14, 2.1), p = 0.03** | 0.86 (0.65, 1.14), p = 0.56 | 0.97 (0.67, 1.42), p = 0.94 | **1.3 (1.11, 1.52), p = 0.01** | **1.38 (1.11, 1.7), p = 0.02** | 1.13 (0.94, 1.36), p = 0.55 | 1.39 (1.09, 1.77), p = 0.09 | 1.29 (0.75, 2.24), p = 0.78 |
| General health | Self-rated health: 1. Poor | 3.02 (0.9, 10.2), p = 0.31 | 3.68 (0.99, 13.7), p = 0.21 | **28.85 (3.8, 219.11), p = 0.009** | **41.09 (8.36, 201.89), p < 0.0001** | Suppressed | Suppressed | Suppressed | Suppressed | 2.06 (0.71, 5.93), p = 0.55 | **5.69 (1.78, 18.2), p = 0.05** | Suppressed |
| General health | Self-rated health: 2. Fair | **2.86 (1.49, 5.46), p = 0.01** | **2.97 (1.37, 6.41), p = 0.05** | 5.81 (1.4, 24.17), p = 0.07 | 2.89 (0.94, 8.83), p = 0.17 | Suppressed | Suppressed | **2.62 (1.41, 4.85), p = 0.01** | **3.27 (1.47, 7.24), p = 0.02** | 1.12 (0.51, 2.43), p = 0.87 | 1.76 (0.62, 4.99), p = 0.49 | 2.62 (0.42, 16.24), p = 0.78 |
| General health | Self-rated health: 3. Good | 1.38 (0.82, 2.32), p = 0.55 | 1.08 (0.54, 2.15), p = 0.91 | 2.6 (0.69, 9.8), p = 0.31 | 2.0 (0.8, 4.98), p = 0.3 | 1.14 (0.53, 2.46), p = 0.86 | 1.39 (0.51, 3.84), p = 0.69 | **1.91 (1.17, 3.12), p = 0.05** | 2.07 (1.07, 4.0), p = 0.14 | 1.25 (0.74, 2.11), p = 0.78 | 1.74 (0.81, 3.75), p = 0.36 | 1.32 (0.24, 7.17), p = 0.91 |
| General health | Self-rated health: 4. Very Good | 1.01 (0.63, 1.64), p = 0.95 | 0.61 (0.31, 1.2), p = 0.42 | 3.36 (0.94, 12.07), p = 0.16 | 2.19 (0.89, 5.34), p = 0.21 | 0.92 (0.5, 1.72), p = 0.89 | 0.85 (0.36, 2.02), p = 0.86 | **1.84 (1.16, 2.92), p = 0.05** | 1.89 (1.0, 3.55), p = 0.21 | 0.99 (0.62, 1.58), p = 0.98 | 1.26 (0.62, 2.55), p = 0.7 | 1.16 (0.27, 4.97), p = 0.96 |
| General health | Self-rated health: Poor, Fair | **2.61 (1.55, 4.38), p = 0.004** | **3.68 (1.99, 6.8), p = 0.0004** | **3.21 (1.41, 7.32), p = 0.03** | **2.69 (1.29, 5.57), p = 0.04** | Suppressed | Suppressed | 1.54 (0.95, 2.49), p = 0.32 | 1.8 (0.97, 3.33), p = 0.25 | 1.26 (0.7, 2.26), p = 0.78 | 1.97 (0.98, 3.94), p = 0.25 | 2.1 (0.51, 8.6), p = 0.78 |
| Mental health | Anxiety (GAD-7): +1 score (21-point scale) | | | | | | | | | 1.05 (1.0, 1.1), p = 0.29 | 1.05 (0.98, 1.12), p = 0.36 | 1.09 (0.93, 1.27), p = 0.78 |
| Mental health | Anxiety (GAD-7): Above threshold | | | | | | | | | 1.27 (0.8, 2.02), p = 0.64 | 1.14 (0.6, 2.16), p = 0.85 | 1.89 (0.48, 7.42), p = 0.78 |
| Mental health | Anxiety (HADS) score: +1 score (21-point scale) | 0.98 (0.93, 1.02), p = 0.61 | 0.96 (0.9, 1.03), p = 0.64 | 1.01 (0.93, 1.1), p = 0.89 | 1.03 (0.96, 1.1), p = 0.62 | 1.03 (0.97, 1.1), p = 0.6 | 1.06 (0.98, 1.15), p = 0.33 | 0.96 (0.93, 1.0), p = 0.16 | 0.97 (0.92, 1.02), p = 0.54 | | | |
| Mental health | Anxiety (HADS): Above threshold, 11+ | 0.8 (0.5, 1.28), p = 0.63 | 0.86 (0.47, 1.58), p = 0.86 | 1.08 (0.42, 2.78), p = 0.93 | 1.28 (0.61, 2.71), p = 0.66 | 1.38 (0.75, 2.54), p = 0.56 | 1.36 (0.61, 3.04), p = 0.66 | 0.71 (0.46, 1.11), p = 0.4 | 0.78 (0.42, 1.46), p = 0.73 | | | |

| | | | | | | | | | | | | |
|---|---|---|---|---|---|---|---|---|---|---|---|---|
| Mental health | Anxiety (HADS): Above threshold, 8-10 | 0.65 (0.4, 1.06), p = 0.31 | 0.37 (0.17, 0.83), p = 0.11 | 1.06 (0.46, 2.41), p = 0.94 | 1.17 (0.6, 2.29), p = 0.74 | 0.62 (0.3, 1.27), p = 0.44 | 0.66 (0.24, 1.79), p = 0.64 | 0.71 (0.47, 1.06), p = 0.34 | 0.85 (0.5, 1.45), p = 0.76 | | | |
| Mental health | Depression (HADS): +1 score (21-point scale) | 1.02 (0.97, 1.07), p = 0.65 | 1.02 (0.96, 1.09), p = 0.76 | 1.05 (0.96, 1.15), p = 0.5 | 1.04 (0.97, 1.12), p = 0.48 | 1.05 (0.98, 1.11), p = 0.44 | 1.08 (0.99, 1.18), p = 0.16 | 0.99 (0.94, 1.03), p = 0.84 | 0.99 (0.93, 1.05), p = 0.88 | | | |
| Mental health | Depression (HADS): Above threshold, 11+ | 0.98 (0.55, 1.74), p = 0.95 | 1.07 (0.49, 2.3), p = 0.93 | 1.05 (0.3, 3.66), p = 0.96 | 0.61 (0.18, 2.1), p = 0.61 | 1.57 (0.8, 3.08), p = 0.44 | 2.04 (0.87, 4.78), p = 0.22 | 0.88 (0.49, 1.56), p = 0.87 | 0.72 (0.3, 1.75), p = 0.73 | | | |
| Mental health | Depression (HADS): Above threshold, 8-10 | 0.91 (0.55, 1.5), p = 0.84 | 0.98 (0.5, 1.91), p = 0.99 | 1.33 (0.53, 3.32), p = 0.7 | 1.35 (0.64, 2.86), p = 0.61 | 0.55 (0.22, 1.34), p = 0.44 | 0.62 (0.18, 2.14), p = 0.66 | 0.8 (0.52, 1.25), p = 0.73 | 1.14 (0.66, 1.98), p = 0.84 | | | |
| Mental health | Depression (SMFQ): +1 score (26-point scale) | | | | | | | | | 1.05 (1.0, 1.11), p = 0.33 | 1.05 (0.98, 1.12), p = 0.36 | 1.06 (0.9, 1.26), p = 0.78 |
| Mental health | Depression (SMFQ): Above threshold | | | | | | | | | 1.74 (0.68, 4.42), p = 0.59 | 2.1 (0.68, 6.47), p = 0.39 | Suppressed |
| Socio-demographics | Age: +1 year | 0.99 (0.97, 1.01), p = 0.61 | 1.0 (0.97, 1.02), p = 0.9 | 1.02 (1.0, 1.04), p = 0.08 | 1.02 (1.0, 1.04), p = 0.12 | 1.0 (0.97, 1.03), p = 0.96 | 1.0 (0.97, 1.04), p = 0.91 | 1.01 (1.0, 1.02), p = 0.62 | 1.01 (0.99, 1.02), p = 0.76 | 0.94 (0.89, 0.99), p = 0.14 | 0.92 (0.86, 0.98), p = 0.13 | 1.04 (0.93, 1.16), p = 0.78 |
| Socio-demographics | Age: 18-49 | | | | | | | | | | | Suppressed |
| Socio-demographics | Age: 40-49 | | | | | | | | | Suppressed | Suppressed | |
| Socio-demographics | Age: 50-59 | 0.83 (0.48, 1.46), p = 0.72 | 1.32 (0.59, 2.92), p = 0.76 | 3.09 (0.3, 31.37), p = 0.5 | 1.06 (0.25, 4.4), p = 0.95 | 0.79 (0.42, 1.5), p = 0.7 | 1.3 (0.55, 3.09), p = 0.7 | 1.02 (0.55, 1.89), p = 0.95 | 0.9 (0.39, 2.09), p = 0.91 | | | |
| Socio-demographics | Age: 60-69 | 0.7 (0.39, 1.26), p = 0.55 | 0.75 (0.32, 1.76), p = 0.76 | 9.79 (1.25, 76.59), p = 0.1 | 3.81 (1.3, 11.13), p = 0.07 | 0.79 (0.35, 1.77), p = 0.77 | 0.84 (0.25, 2.79), p = 0.86 | 0.95 (0.55, 1.65), p = 0.92 | 0.84 (0.4, 1.8), p = 0.84 | 0.76 (0.52, 1.13), p = 0.55 | 0.69 (0.4, 1.18), p = 0.38 | 2.72 (0.87, 8.49), p = 0.78 |
| Socio-demographics | Age: 70-79 | 0.72 (0.32, 1.61), p = 0.65 | 0.81 (0.26, 2.5), p = 0.9 | 8.02 (1.06, 60.54), p = 0.13 | 3.14 (1.12, 8.78), p = 0.11 | 0.93 (0.3, 2.89), p = 0.95 | Suppressed | 1.12 (0.68, 1.86), p = 0.87 | 1.04 (0.53, 2.05), p = 0.94 | Suppressed | Suppressed | 0.41 (0.03, 5.54), p = 0.78 |
| Socio-demographics | Age: 80+ | Suppressed | Suppressed | 5.37 (0.63, 45.51), p = 0.27 | 2.15 (0.68, 6.76), p = 0.36 | Suppressed | Suppressed | 1.07 (0.56, 2.07), p = 0.92 | 1.14 (0.49, 2.67), p = 0.9 | | | |

| | | | | | | | | | | | | |
|---|---|---|---|---|---|---|---|---|---|---|---|---|
| Socio-demographics | Deprivation (IMD): Middle 40% (decile 4-7) | 0.73 (0.49, 1.08), p = 0.39 | 0.8 (0.47, 1.36), p = 0.76 | 2.4 (1.14, 5.07), p = 0.08 | 1.88 (1.04, 3.37), p = 0.11 | 1.15 (0.66, 2.01), p = 0.82 | 1.34 (0.63, 2.87), p = 0.66 | 1.08 (0.78, 1.5), p = 0.87 | 1.33 (0.86, 2.06), p = 0.54 | | | |
| Socio-demographics | Deprivation (IMD): Most deprived 30% (decile 1-3) | 1.29 (0.73, 2.28), p = 0.63 | 1.32 (0.61, 2.85), p = 0.76 | 3.44 (1.17, 10.07), p = 0.09 | 2.45 (0.97, 6.21), p = 0.17 | 1.21 (0.55, 2.68), p = 0.82 | 1.26 (0.43, 3.76), p = 0.86 | 0.83 (0.46, 1.53), p = 0.87 | 1.43 (0.7, 2.92), p = 0.61 | | | |
| Socio-demographics | Deprivation (IMD): Most deprived 40% (decile 1-4) | 1.49 (0.94, 2.36), p = 0.31 | 1.6 (0.87, 2.93), p = 0.39 | 1.92 (0.84, 4.39), p = 0.27 | 1.29 (0.61, 2.74), p = 0.66 | 0.97 (0.51, 1.84), p = 0.96 | 1.16 (0.5, 2.68), p = 0.86 | 1.19 (0.78, 1.8), p = 0.79 | 1.48 (0.87, 2.52), p = 0.46 | 0.87 (0.54, 1.4), p = 0.81 | 0.79 (0.41, 1.52), p = 0.65 | 0.6 (0.13, 2.63), p = 0.78 |
| Socio-demographics | Employment status: In education | Suppressed | Suppressed | Suppressed | Suppressed | Suppressed | Suppressed | Suppressed | Suppressed | Suppressed | Suppressed | |
| Socio-demographics | Employment status: Looking after home or family (unpaid care) | 0.92 (0.34, 2.48), p = 0.94 | Suppressed | Suppressed | Suppressed | 1.66 (0.43, 6.37), p = 0.7 | Suppressed | 0.79 (0.28, 2.2), p = 0.87 | Suppressed | 0.81 (0.28, 2.36), p = 0.87 | 0.84 (0.19, 3.68), p = 0.88 | 7.87 (0.12, 520.99), p = 0.78 |
| Socio-demographics | Employment status: Maternity leave | | | | | Suppressed | Suppressed | | | | | |
| Socio-demographics | Employment status: Other | Suppressed | Suppressed | Suppressed | Suppressed | Suppressed | Suppressed | Suppressed | Suppressed | | | |
| Socio-demographics | Employment status: Permanently (or long-term) sick or disabled | 2.64 (0.69, 10.13), p = 0.47 | Suppressed | Suppressed | **13.66 (2.25, 83.13), p = 0.03** | Suppressed | Suppressed | 1.41 (0.41, 4.91), p = 0.87 | Suppressed | Suppressed | Suppressed | Suppressed |
| Socio-demographics | Employment status: Retired | 1.4 (0.83, 2.36), p = 0.55 | 1.0 (0.52, 1.94), p = 0.99 | 2.88 (0.68, 12.27), p = 0.31 | 2.73 (0.9, 8.29), p = 0.2 | 0.85 (0.3, 2.48), p = 0.89 | 0.97 (0.24, 4.02), p = 0.97 | 1.05 (0.63, 1.74), p = 0.92 | 1.56 (0.81, 3.02), p = 0.54 | 0.87 (0.51, 1.48), p = 0.85 | 0.66 (0.29, 1.46), p = 0.5 | 0.51 (0.08, 3.12), p = 0.78 |
| Socio-demographics | Employment status: Self-employed | 1.53 (0.88, 2.64), p = 0.41 | 1.31 (0.62, 2.77), p = 0.76 | Suppressed | Suppressed | 0.72 (0.28, 1.84), p = 0.7 | 0.81 (0.23, 2.86), p = 0.86 | 0.86 (0.4, 1.85), p = 0.91 | 1.48 (0.57, 3.85), p = 0.73 | 1.23 (0.71, 2.14), p = 0.78 | 1.93 (1.0, 3.72), p = 0.25 | 0.74 (0.08, 7.03), p = 0.94 |
| Socio-demographics | Employment status: Semi-retired/part-time employment | Suppressed | Suppressed | Suppressed | Suppressed | Suppressed | Suppressed | Suppressed | Suppressed | | | |
| Socio-demographics | Employment status: Unemployed | Suppressed | Suppressed | | | Suppressed | Suppressed | Suppressed | Suppressed | 1.22 (0.35, 4.33), p = 0.87 | 2.79 (0.77, 10.05), p = 0.35 | Suppressed |
| Socio-demographics | Employment status: Unpaid/voluntary work | Suppressed | Suppressed | Suppressed | Suppressed | Suppressed | Suppressed | 0.94 (0.31, 2.84), p = 0.95 | Suppressed | Suppressed | Suppressed | Suppressed |

| | | | | | | | | | | | | |
|---|---|---|---|---|---|---|---|---|---|---|---|---|
| Socio-demographics | Ethnicity: Other than white | Suppressed | Suppressed | Suppressed | Suppressed | Suppressed | Suppressed | 1.76 (0.66, 4.7), p = 0.62 | 2.3 (0.8, 6.57), p = 0.42 | Suppressed | Suppressed | Suppressed |
| Socio-demographics | IMD: -1 decile (increasing deprivation) | 1.03 (0.95, 1.11), p = 0.65 | 1.04 (0.94, 1.15), p = 0.76 | 1.15 (1.01, 1.32), p = 0.11 | 1.11 (0.99, 1.24), p = 0.18 | 1.02 (0.93, 1.12), p = 0.85 | 1.06 (0.94, 1.2), p = 0.54 | 0.99 (0.93, 1.06), p = 0.92 | 1.06 (0.98, 1.15), p = 0.49 | | | |
| Socio-demographics | IMD: -1 quintile (increasing deprivation) | | | | | | | | | 1.0 (0.85, 1.17), p = 0.98 | 1.02 (0.83, 1.26), p = 0.88 | 0.87 (0.53, 1.45), p = 0.86 |
| Socio-demographics | Highest educational attainment: NVQ level 3 or lower | 0.88 (0.59, 1.32), p = 0.72 | 0.91 (0.52, 1.59), p = 0.9 | **2.76 (1.36, 5.62), p = 0.03** | **2.23 (1.26, 3.94), p = 0.03** | 1.09 (0.6, 1.98), p = 0.89 | 1.57 (0.71, 3.47), p = 0.49 | 1.03 (0.74, 1.44), p = 0.92 | 1.04 (0.67, 1.62), p = 0.92 | 1.42 (0.93, 2.18), p = 0.48 | 1.82 (0.98, 3.38), p = 0.25 | 1.02 (0.25, 4.15), p = 0.99 |
| Socio-demographics | RUC: Urban | 1.01 (0.68, 1.51), p = 0.95 | 1.28 (0.71, 2.3), p = 0.76 | 1.49 (0.66, 3.36), p = 0.5 | 0.88 (0.49, 1.61), p = 0.78 | 0.5 (0.29, 0.87), p = 0.05 | 0.78 (0.36, 1.68), p = 0.69 | 1.16 (0.8, 1.66), p = 0.79 | 1.0 (0.62, 1.62), p = 0.99 | | | |
| Socio-demographics | Sex: Male | 1.4 (0.86, 2.27), p = 0.49 | 1.14 (0.57, 2.28), p = 0.9 | 0.63 (0.19, 2.13), p = 0.62 | 0.64 (0.26, 1.58), p = 0.55 | 1.06 (0.5, 2.25), p = 0.93 | 1.37 (0.53, 3.5), p = 0.69 | 0.79 (0.47, 1.35), p = 0.77 | 0.63 (0.3, 1.35), p = 0.55 | 1.62 (1.1, 2.37), p = 0.13 | 1.64 (0.97, 2.76), p = 0.25 | 1.99 (0.56, 7.04), p = 0.78 |

### Factors associated with low antibody levels: TwinsUK figures

#### TwinsUK figures: Socio-demographic variables

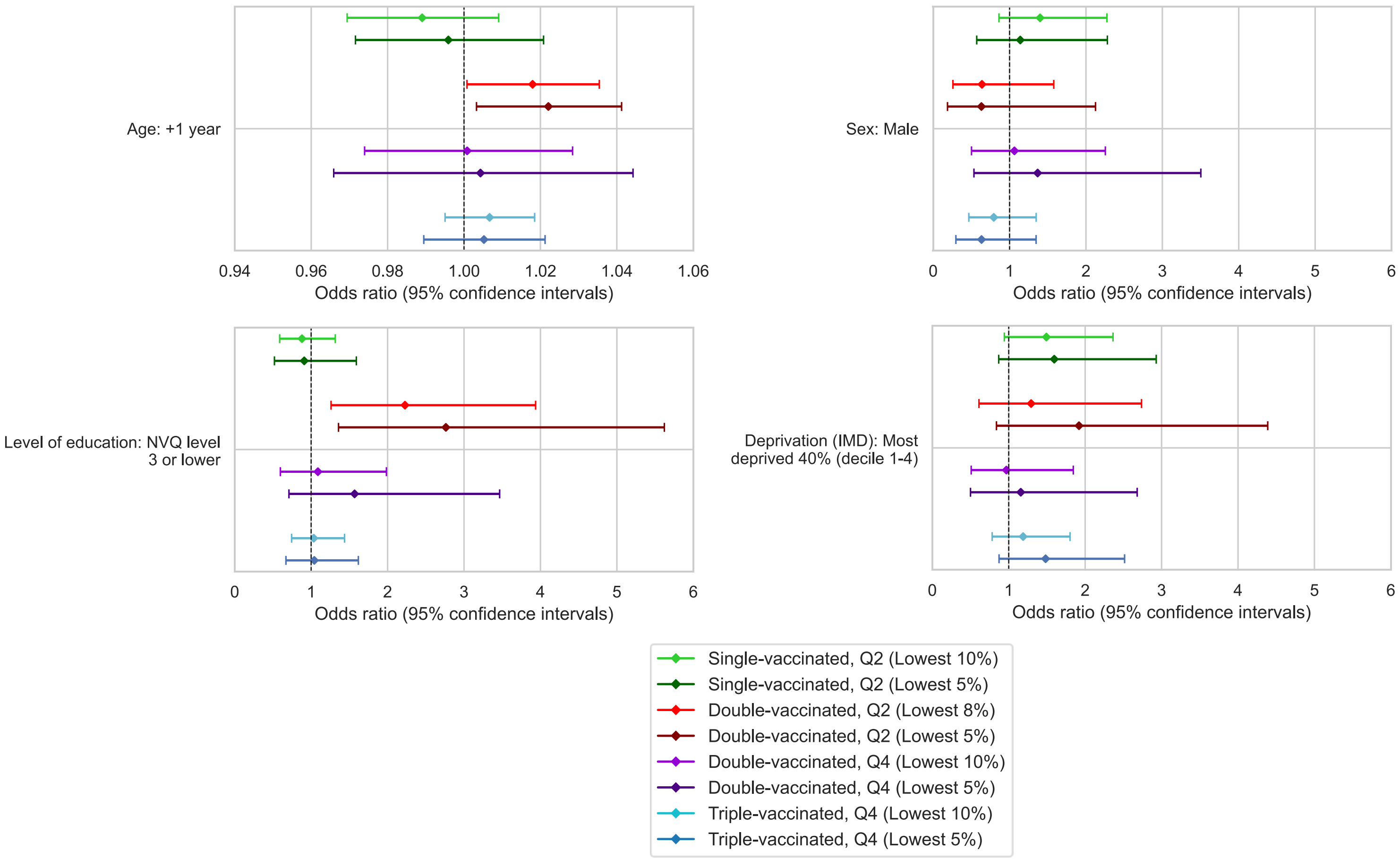

Figure S 7. Odds ratios with unadjusted 95% confidence intervals for selected socio-demographic variables, testing associations with low anti-Spike antibody levels (lowest 5%, 8% or 10%, as indicated in legend), for TwinsUK individuals tested in Q2 or Q4, after first, second and third vaccination. Each sub-plot presents results from a distinct model, including the variable of interest and adjustment variables of age, sex, name of most recent vaccine received and weeks since most recent vaccination. Note x-axis ranges on subplots vary. Odds ratio = 1 is indicated with a dashed black line.

#### TwinsUK figures: SARS-CoV-2 vaccination variables

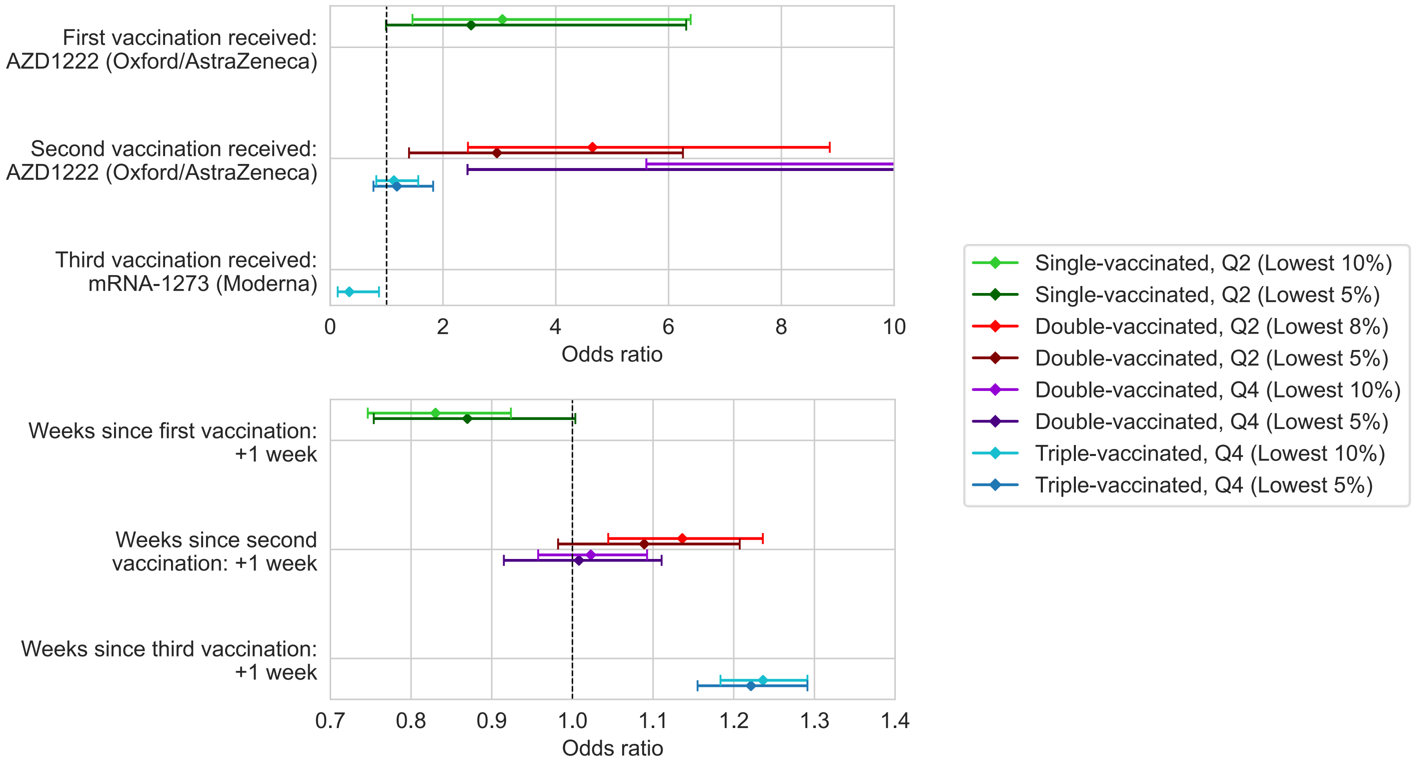

Figure S 8. Odds ratios with unadjusted 95% confidence intervals for selected SARS-CoV-2 vaccination variables, testing associations with low anti-Spike antibody levels (lowest 5%, 8% or 10%, as indicated in legend), for TwinsUK individuals tested in Q2 or Q4, after first, second and third vaccination. Each sub-plot presents results from a distinct model, including the variable of interest and adjustment variables of age, sex, name of most recent vaccine received and weeks since most recent vaccination. Note x-axis ranges on subplots vary. Odds ratio = 1 is indicated with a dashed black line.

#### TwinsUK figures: SARS-CoV-2 infection variables

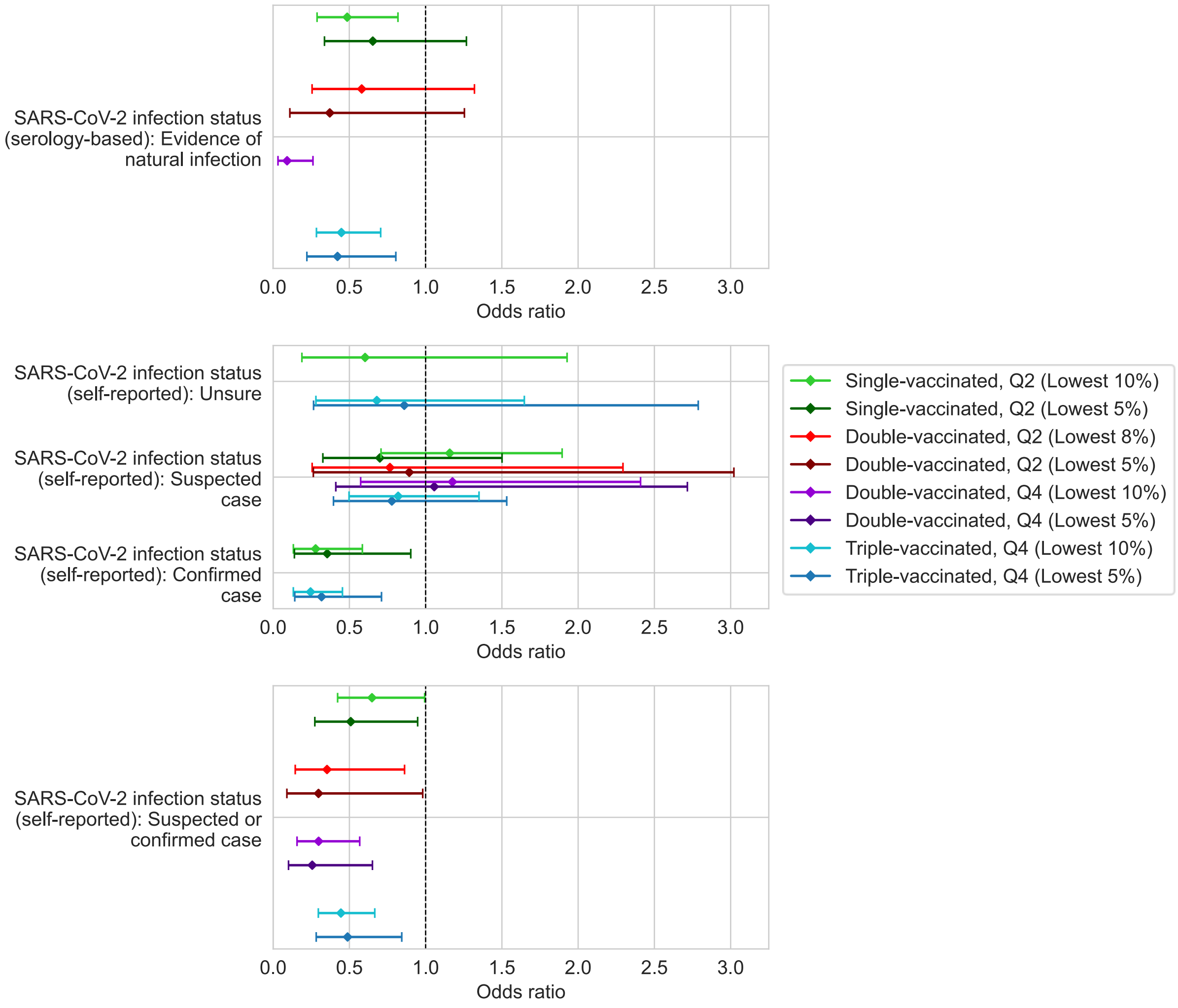

Figure S 9. Odds ratios with unadjusted 95% confidence intervals for selected SARS-CoV-2 infection status variables, testing associations with low anti-Spike antibody levels (lowest 5%, 8% or 10%, as indicated in legend), for TwinsUK individuals tested in Q2 or Q4, after first, second and third vaccination. Each sub-plot presents results from a distinct model, including the variable of interest and adjustment variables of age, sex, name of most recent vaccine received and weeks since most recent vaccination. Note x-axis ranges on subplots vary. Odds ratio = 1 is indicated with a dashed black line.

#### TwinsUK figures: COVID-19 at-risk variables

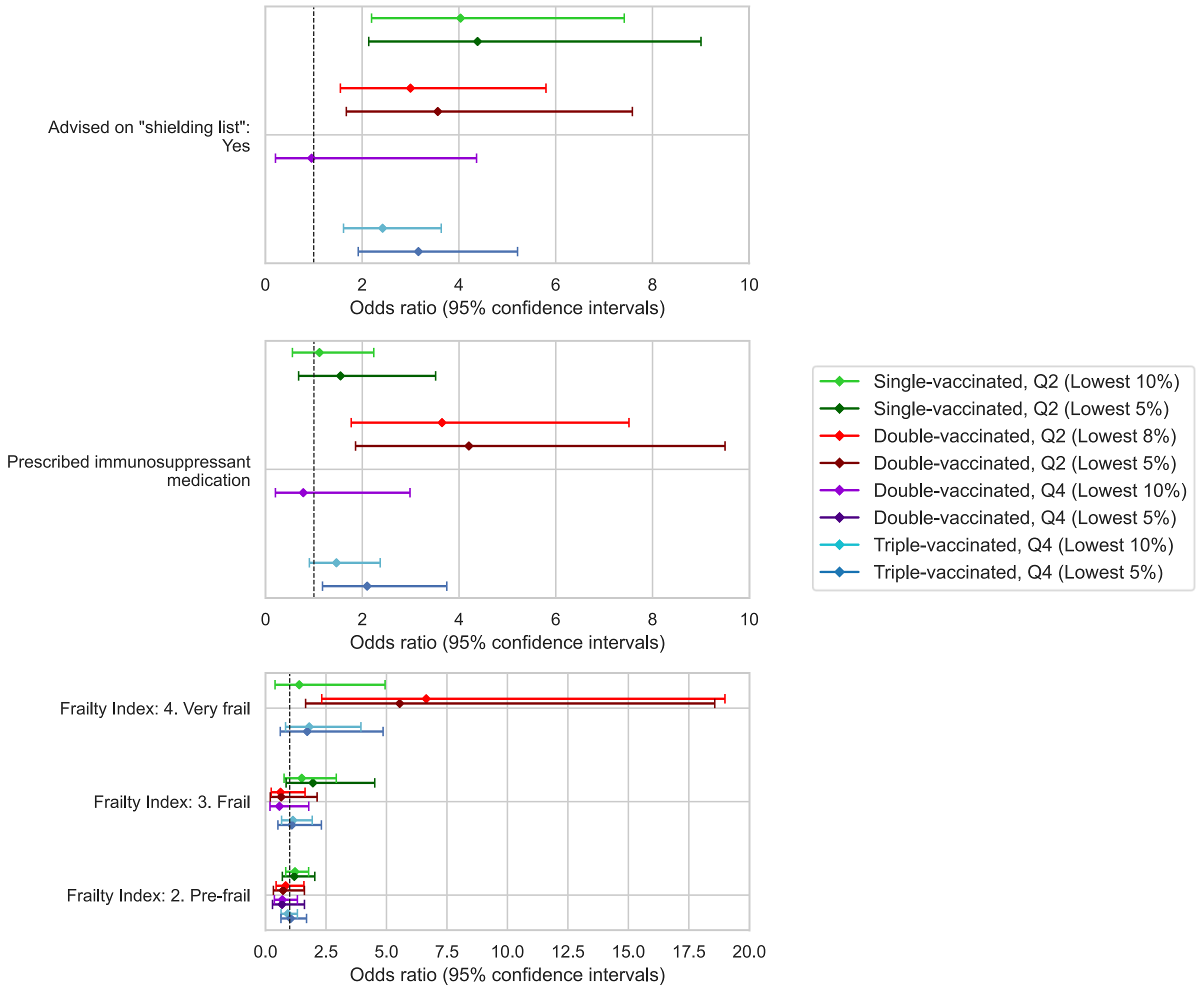

Figure S 10. Odds ratios with unadjusted 95% confidence intervals for selected variables previously identified as COVID-19 at-risk groups, testing associations with low anti-Spike antibody levels (lowest 5%, 8% or 10%, as indicated in legend), for TwinsUK individuals tested in Q2 or Q4, after first, second and third vaccination. Each sub-plot presents results from a distinct model, including the variable of interest and adjustment variables of age, sex, name of most recent vaccine received and weeks since most recent vaccination. Note x-axis ranges on subplots vary. Odds ratio = 1 is indicated with a dashed black line.

#### TwinsUK figures: General health variables

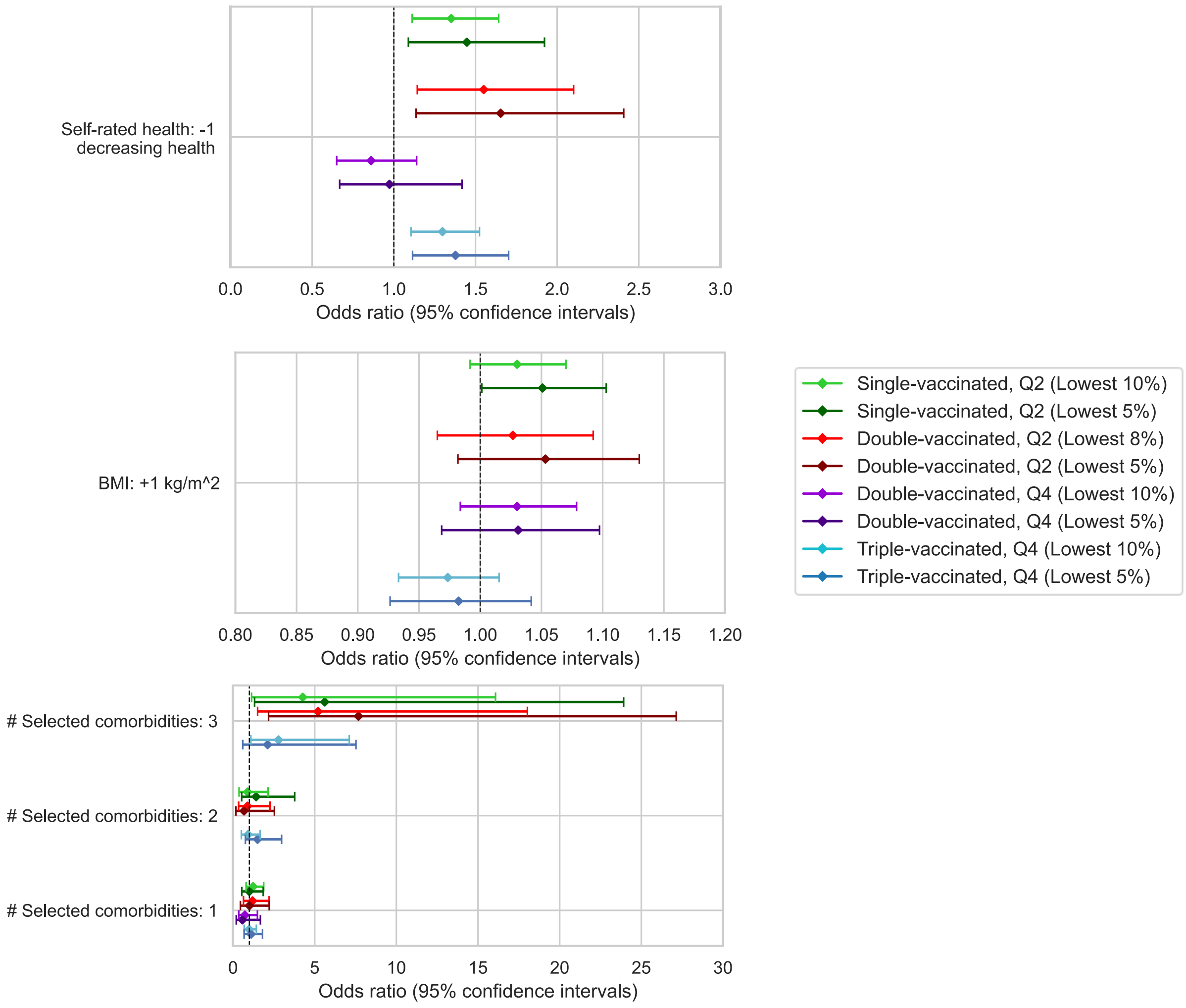

Figure S 11. Odds ratios with unadjusted % confidence intervals for selected general health variables, testing associations with low anti-Spike antibody levels (lowest 5%, 8% or 10%, as indicated in legend), for TwinsUK individuals tested in Q2 or Q4, after first, second and third vaccination. Each sub-plot presents results from a distinct model, including the variable of interest and adjustment variables of age, sex, name of most recent vaccine received and weeks since most recent vaccination. Note x-axis ranges on subplots vary. Odds ratio = 1 is indicated with a dashed black line.

#### TwinsUK figures: Mental health variables

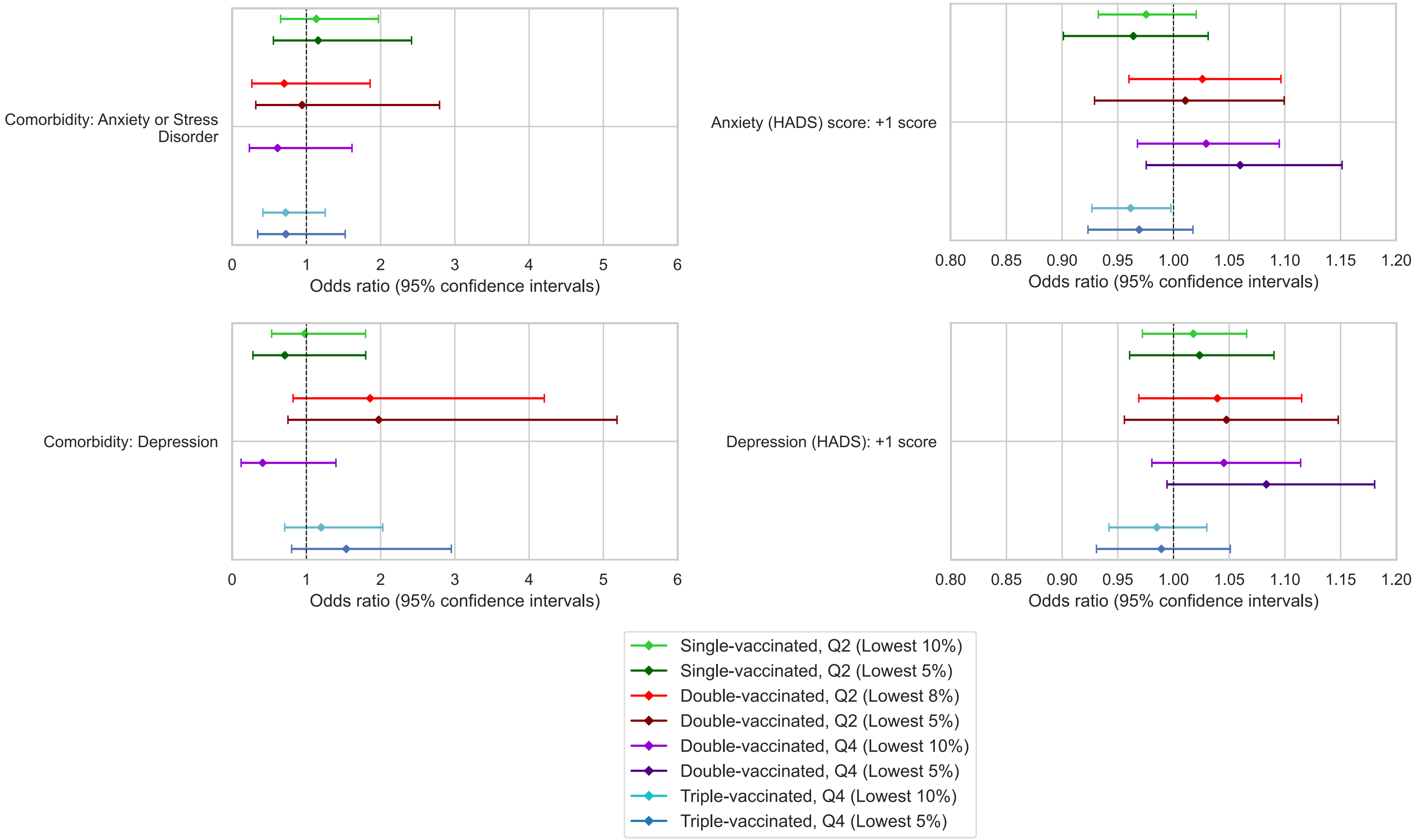

Figure S 12. Odds ratios with unadjusted 95% confidence intervals for selected mental health variables, testing associations with low anti-Spike antibody levels (lowest 5%, 8% or 10%, as indicated in legend), for TwinsUK individuals tested in Q2 or Q4, after first, second and third vaccination. Each sub-plot presents results from a distinct model, including the variable of interest and adjustment variables of age, sex, name of most recent vaccine received and weeks since most recent vaccination. Note x-axis ranges on subplots vary. Odds ratio = 1 is indicated with a dashed black line.

#### TwinsUK figures: Comorbidities

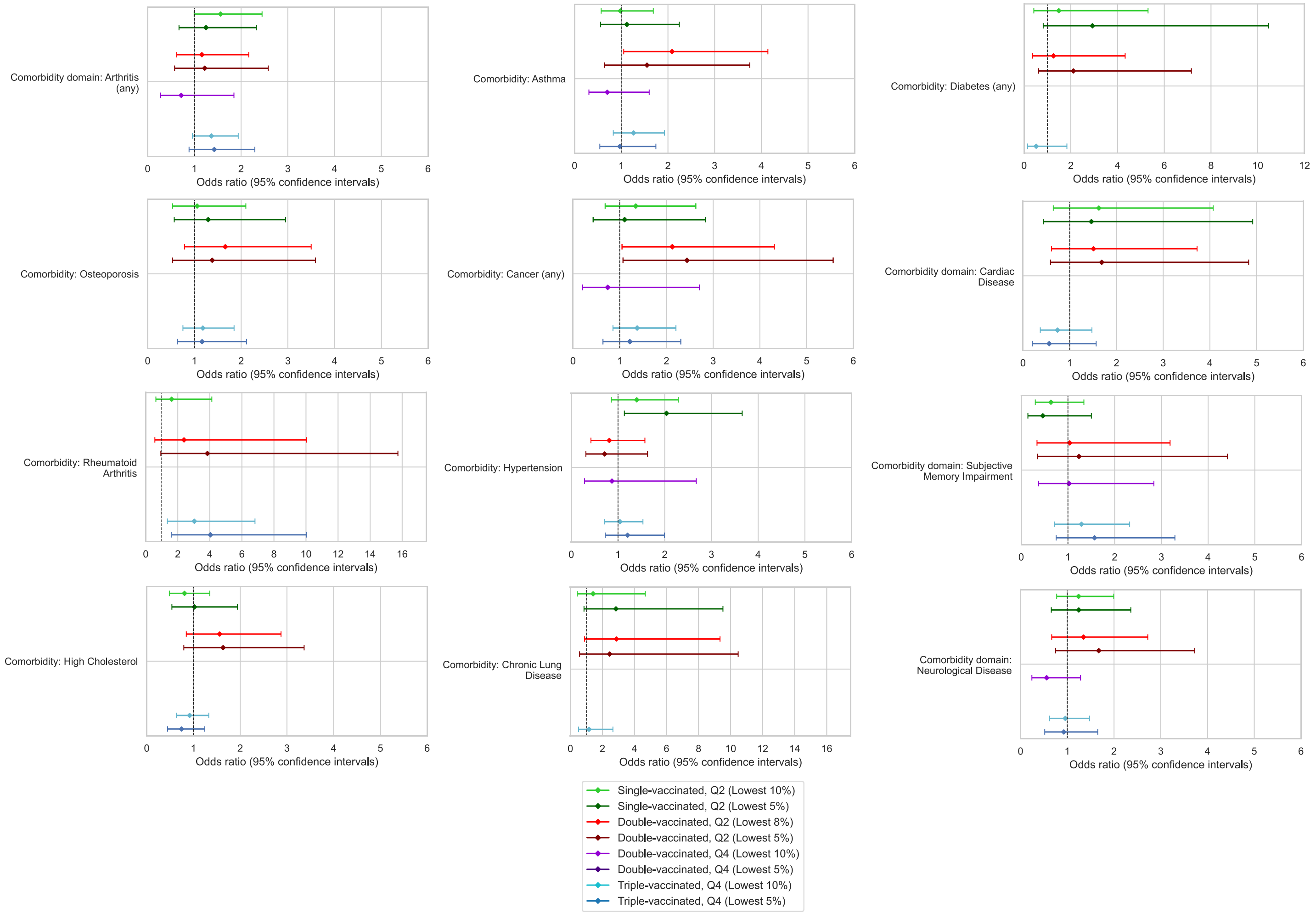

Figure S 13. Odds ratios with unadjusted 95% confidence intervals for various selected individual comorbidities and comorbidity domains, testing associations with low anti-Spike antibody levels (lowest 5%, 8% or 10%, as indicated in legend), for TwinsUK individuals tested in Q2 or Q4, after first, second and third vaccination. Each sub-plot presents results from a distinct model, including the variable of interest and adjustment variables of age, sex, name of most recent vaccine received and weeks since most recent vaccination. Note x-axis ranges on subplots vary. Odds ratio = 1 is indicated with a dashed black line.

#### Factors associated with low antibody levels: Age, sex and BMI (combined ALSPAC & TwinsUK)

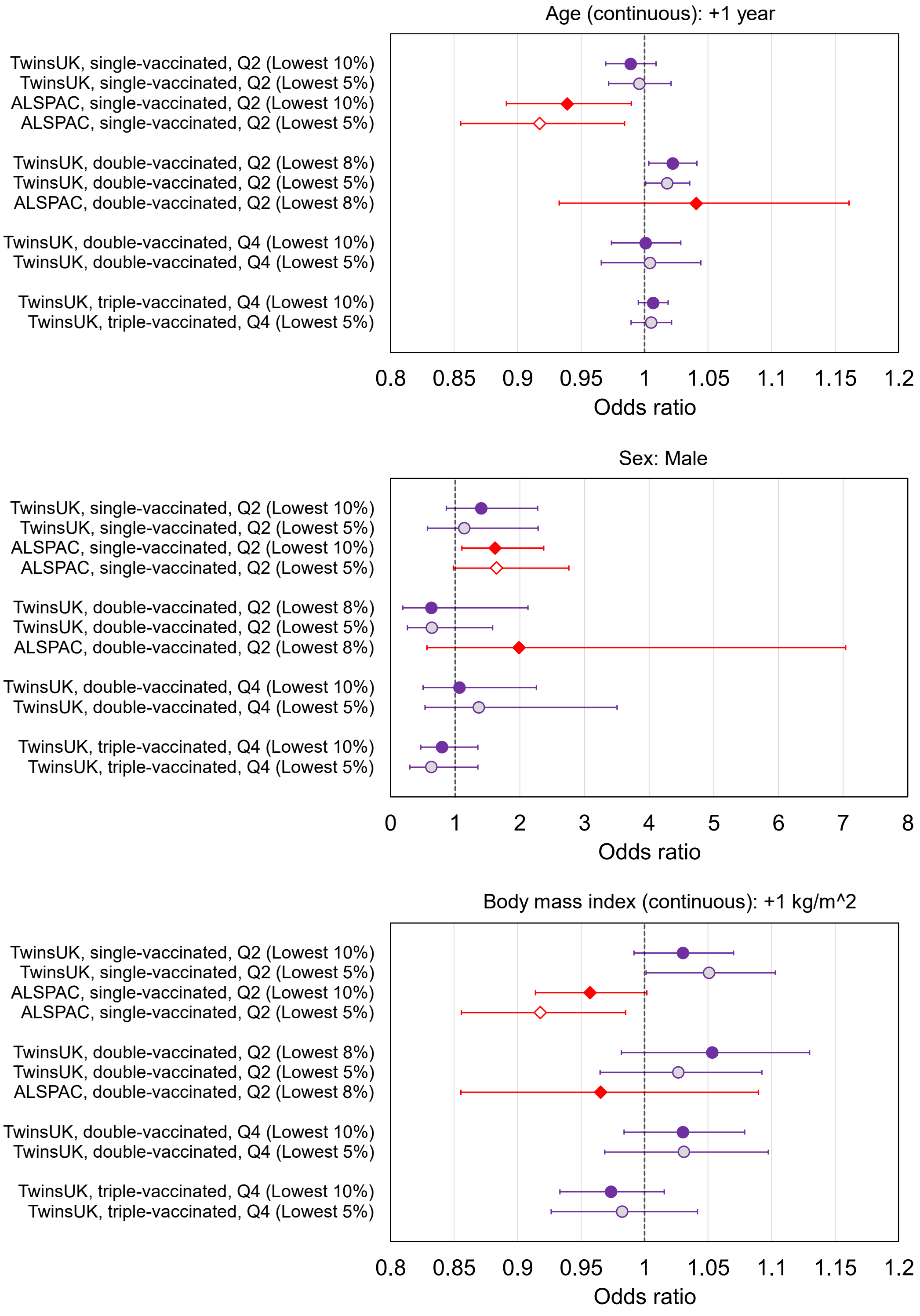

Figure S 14. Odds ratios with unadjusted 95% confidence intervals for selected variables, testing associations with low anti-Spike antibody levels (lowest 5%, 8% or 10%), for TwinsUK (purple circles) and ALSPAC (red diamonds) individuals tested in Q2 or Q4, after first, second and third vaccination. Each point estimate originates from a distinct multivariate logistic regression model, including the variable of interest and adjustment variables of age, sex, name of most recent vaccine received and weeks since most recent vaccination. Note x-axis ranges on subplots vary. Odds ratio = 1 is indicated with a dashed black line.

#### Twin-pair analysis: Antibody level differences within related pairs and between non-related pairs

Table S 10. Descriptive statistics of differences in anti-Spike antibody levels between pairs after third SARS-CoV-2 vaccination within TwinsUK. Pair-differences are calculated between all complete pairs of monozygotic (MZ) twins and/or dizygotic (DZ) twins, and all combinations of non-related pairs.

| | Intra-pair difference in anti-Spike levels (BAU/mL) after third vaccination | | | |
|---|---|---|---|---|
| | **Difference between non-related pairs** | **Difference within related pairs** | **Difference within DZ pairs** | **Difference within MZ pairs** |
| **Count of pair differences** | 1874561 | 455 | 167 | 286 |
| **Mean** | 8744 | 7037 | 8061 | 6444 |
| **Standard deviation** | 6369 | 5735 | 6112 | 5441 |
| **Minimum** | 0 | 0 | 0 | 0 |
| **25%** | 3205 | 2332 | 2770.25 | 1984 |
| **Median** | 7862 | 5678 | 6831 | 4981 |
| **75%** | 13546 | 10934 | 12481 | 9715 |
| **Maximum** | 25000 | 24625 | 23189 | 24625 |

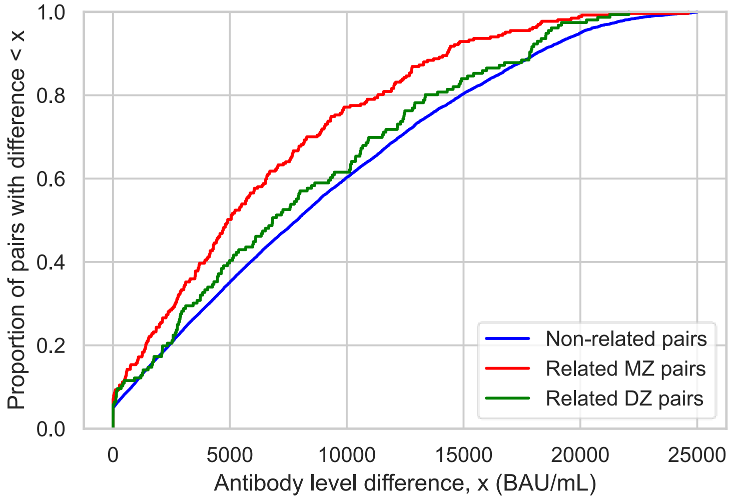

Figure S 15. Empirical cumulative distribution functions describing the difference in anti-Spike antibody levels after third SARS-CoV-2 vaccination within TwinsUK, with pair-differences calculated between all complete pairs of related monozygotic (MZ) twins, dizygotic (DZ) twins, and all combinations of non-related pairs.

#### Twin-pair analysis: Within-between regression models

Results of ‘within-between’ twin-pair generalised linear regression analysis testing association with anti-Spike antibody levels after third vaccination are also provided in supplementary spreadsheet file for easier viewing.

Table S 11. Results of generalised linear mixed effects models testing association with anti-Spike antibody levels after third SARS-CoV-2 vaccination within and between twin-pairs within TwinsUK. Coefficients with unadjusted 95% confidence intervals and unadjusted p-values are presented. Family structure is included as a random effect, allowing intercepts to vary between twin-pairs. Models are adjusted for age, sex, weeks since third vaccination, third vaccine received and serology-based infection status. Variables with (two-sided) p-values < 0.05 are highlighted in bold.

| **Exposure variable** | **Self-rated health (ordinal 1-5 scale, unit: +1, increasing health)** | | **Advised on "Shielded Patient List" (0: No, 1: Yes)** | | **Comorbidity: Rheumatoid arthritis (0: No, 1: Yes)** | | **Prescribed immunosuppressant medication (0: No, 1: Yes)** | | **Highest educational attainment (0: NVQ level 3 or lower, 1: NVQ level 4 or higher)** | | **Frailty index (log-transformed, unit: +1 standard deviation, increasing frailty)** | | **Multimorbidity (0: < 3/5 comorbidities, 1: 3/5 comorbidities)** | |
|---|---|---|---|---|---|---|---|---|---|---|---|---|---|---|
| **Zygosity** | **MZ** | **DZ** | **MZ** | **DZ** | **MZ** | **DZ** | **MZ** | **DZ** | **MZ** | **DZ** | **MZ** | **DZ** | **MZ** | **DZ** |
| **n** | 554 | 322 | 572 | 334 | 572 | 334 | 572 | 334 | 572 | 334 | 486 | 308 | 572 | 334 |
| **Exposure variable: Family mean ('between-pair')** | **2628.02 (1958.61, 3297.44), p < 0.0001** | **1112.97 (14.1, 2211.83), p = 0.05** | -1835.79 (-5199.34, 1527.77), p = 0.28 | 1186.88 (-2790.16, 5163.91), p = 0.56 | -7410.35 (-16435.28, 1614.58), p = 0.11 | **-7066.57 (-13662.52, -470.61), p = 0.04** | -1520.6 (-4845.39, 1804.2), p = 0.37 | -3400.74 (-7132.33, 330.85), p = 0.07 | **3021.69 (1416.43, 4626.94), p = 0.0002** | 1126.5 (-880.9, 3133.89), p = 0.27 | **-1492.75 (-2408.54, -576.96), p = 0.001** | -488.47 (-1761.03, 784.09), p = 0.45 | 2243.69 (-9226.11, 13713.48), p = 0.7 | -5830.71 (-14130.41, 2469.0), p = 0.17 |
| **Exposure variable: Individual difference to family mean ('within-pair')** | 643.13 (-316.09, 1602.35), p = 0.19 | -475.4 (-1648.82, 698.03), p = 0.43 | **-3684.38 (-6491.81, -876.96), p = 0.01** | -2975.43 (-6816.68, 865.83), p = 0.13 | **-5799.45 (-11176.03, -422.86), p = 0.03** | -2842.12 (-9956.15, 4271.91), p = 0.43 | -1733.37 (-4088.0, 621.26), p = 0.15 | -1494.04 (-4518.44, 1530.36), p = 0.33 | -96.53 (-2067.99, 1874.94), p = 0.92 | -1747.06 (-4260.26, 766.15), p = 0.17 | 92.9 (-927.72, 1113.53), p = 0.86 | -873.85 (-2223.79, 476.1), p = 0.2 | -6026.79 (-12830.25, 776.68), p = 0.08 | 2646.79 (-4510.21, 9803.79), p = 0.47 |
| ***Wald test of ‘between-pair’ = ‘within-pair’*** | **p =0.0008** | p = 0.05 | p = 0.41 | p = 0.14 | p = 0.76 | p = 0.39 | p = 0.92 | p = 0.44 | **p = 0.02** | p = 0.08 | **p = 0.02** | p = 0.68 | p = 0.22 | p = 0.13 |
| **Age (unit: +1 year)** | **116.95 (78.99, 154.91), p < 0.0001** | **196.14 (133.23, 259.06), p < 0.0001** | **244.21 (222.03, 266.4), p < 0.0001** | **253.43 (224.45, 282.41), p < 0.0001** | **243.07 (220.96, 265.18), p < 0.0001** | **258.87 (229.88, 287.86), p < 0.0001** | **245.37 (222.57, 268.18), p < 0.0001** | **259.64 (230.14, 289.14), p < 0.0001** | **222.48 (198.07, 246.89), p < 0.0001** | **249.03 (218.2, 279.86), p < 0.0001** | **241.09 (217.23, 264.96), p < 0.0001** | **256.93 (227.48, 286.39), p < 0.0001** | **242.76 (220.61, 264.9), p < 0.0001** | **255.68 (226.61, 284.74), p < 0.0001** |

| | | | | | | | | | | | | | | |
|---|---|---|---|---|---|---|---|---|---|---|---|---|---|---|
| **Sex: Male (reference: Female)** | -1206.07 (-3313.35, 901.22), p = 0.26 | 172.32 (-3254.85, 3599.5), p = 0.92 | -994.16 (-3241.51, 1253.19), p = 0.39 | 101.98 (-3402.5, 3606.45), p = 0.95 | -943.01 (-3180.95, 1294.92), p = 0.41 | -118.75 (-3582.72, 3345.21), p = 0.95 | -1027.05 (-3283.86, 1229.76), p = 0.37 | -120.4 (-3598.88, 3358.08), p = 0.95 | -937.44 (-3136.95, 1262.07), p = 0.4 | 51.9 (-3451.63, 3555.42), p = 0.98 | -1097.0 (-3670.17, 1476.16), p = 0.4 | -1225.65 (-5250.07, 2798.78), p = 0.55 | -971.71 (-3235.26, 1291.83), p = 0.4 | 2.67 (-3480.8, 3486.13), p = 1.0 |
| **Weeks since third vaccination (unit: +1 week)** | **-760.4 (-975.36, -545.44), p < 0.0001** | **-825.15 (-1112.87, -537.44), p < 0.0001** | **-577.95 (-793.54, -362.37), p < 0.0001** | **-831.28 (-1123.09, -539.47), p < 0.0001** | **-575.56 (-790.64, -360.48), p < 0.0001** | **-839.98 (-1127.12, -552.84), p < 0.0001** | **-593.74 (-809.02, -378.46), p < 0.0001** | **-814.6 (-1102.88, -526.31), p < 0.0001** | **-626.04 (-840.02, -412.07), p < 0.0001** | **-843.02 (-1131.61, -554.43), p < 0.0001** | **-626.13 (-865.16, -387.11), p < 0.0001** | **-825.41 (-1124.41, -526.41), p < 0.0001** | **-587.59 (-803.05, -372.13), p < 0.0001** | **-818.93 (-1108.37, -529.48), p < 0.0001** |
| **Third vaccine: mRNA-1273 (reference: BNT162b2)** | **3763.01 (1795.99, 5730.04), p = 0.0002** | 2770.26 (-225.02, 5765.53), p = 0.07 | **5016.49 (3067.23, 6965.74), p < 0.0001** | **3256.26 (286.13, 6226.39), p = 0.03** | **5235.92 (3283.09, 7188.75), p < 0.0001** | **3051.53 (98.1, 6004.97), p = 0.04** | **5150.06 (3190.12, 7110.0), p < 0.0001** | **3396.55 (439.76, 6353.35), p = 0.02** | **4778.81 (2825.59, 6732.04), p < 0.0001** | **2988.59 (6.4, 5970.77), p = 0.05** | **5117.66 (2870.42, 7364.89), p < 0.0001** | 2621.63 (-473.77, 5717.04), p = 0.1 | **5064.95 (3107.49, 7022.41), p < 0.0001** | **3332.52 (362.72, 6302.31), p = 0.03** |
| **Third vaccine: Other (reference: BNT162b2)** | -2958.22 (-7800.03, 1883.59), p = 0.23 | 4712.73 (-2337.57, 11763.03), p = 0.19 | -2437.17 (-7369.43, 2495.09), p = 0.33 | 4996.75 (-2140.9, 12134.4), p = 0.17 | -2700.36 (-7632.39, 2231.67), p = 0.28 | 4623.3 (-2496.36, 11742.96), p = 0.2 | -2366.81 (-7326.11, 2592.5), p = 0.35 | 5062.52 (-2076.14, 12201.17), p = 0.16 | -2640.2 (-7575.46, 2295.06), p = 0.29 | 4859.49 (-2270.14, 11989.11), p = 0.18 | -2384.03 (-7411.74, 2643.69), p = 0.35 | 4693.79 (-2381.22, 11768.79), p = 0.19 | -2649.17 (-7592.78, 2294.44), p = 0.29 | 4743.8 (-2399.78, 11887.37), p = 0.19 |
| **SARS-CoV-2 infection status (serology-based): Evidence of natural infection (reference: No evidence)** | **2871.98 (1569.76, 4174.2), p < 0.0001** | **4798.46 (2800.73, 6796.18), p < 0.0001** | **3592.85 (2285.58, 4900.11), p < 0.0001** | **4883.75 (2869.54, 6897.95), p < 0.0001** | **3583.58 (2275.31, 4891.86), p < 0.0001** | **4893.27 (2907.17, 6879.36), p < 0.0001** | **3593.19 (2276.94, 4909.45), p < 0.0001** | **4803.36 (2811.87, 6794.85), p < 0.0001** | **3683.59 (2375.91, 4991.26), p < 0.0001** | **4825.56 (2823.54, 6827.59), p < 0.0001** | **3585.43 (2133.75, 5037.12), p < 0.0001** | **4494.55 (2415.7, 6573.4), p < 0.0001** | **3495.63 (2182.19, 4809.08), p < 0.0001** | **4904.14 (2911.29, 6896.99), p < 0.0001** |
| **Group variance** | **0.68 (0.4, 0.95), p < 0.0001** | 0.14 (-0.06, 0.35), p = 0.17 | **0.89 (0.56, 1.23), p < 0.0001** | 0.16 (-0.05, 0.37), p = 0.13 | **0.88 (0.55, 1.2), p < 0.0001** | 0.14 (-0.06, 0.34), p = 0.17 | **0.88 (0.55, 1.2), p < 0.0001** | 0.15 (-0.06, 0.35), p = 0.16 | **0.81 (0.5, 1.12), p < 0.0001** | 0.16 (-0.05, 0.37), p = 0.13 | **0.68 (0.38, 0.98), p < 0.0001** | 0.21 (-0.02, 0.44), p = 0.07 | **0.89 (0.56, 1.22), p < 0.0001** | 0.15 (-0.05, 0.35), p = 0.15 |
